# The Impact of the UK COVID-19 Lockdown on the Screening, Diagnostics and Incidence of Breast, Colorectal, Lung and Prostate Cancer in the UK: a Population-Based Cohort Study

**DOI:** 10.1101/2023.07.21.23292937

**Authors:** Nicola L Barclay, Marta Pineda Moncusí, Annika M. Jödicke, Daniel Prieto-Alhambra, Berta Raventós, Danielle Newby, Antonella Delmestri, Wai Yi Man, Xihang Chen, Marti Català, The OPTIMA Consortium

**Author notes:** Correspondence to: **Marta Pineda Moncusi**, Centre for Statistics in Medicine, Nuffield Department of Orthopaedics, Rheumatology and Musculoskeletal Sciences (NDORMS), University of Oxford, Botnar Research Centre, Old Road, Headington, Oxford, OX3 7LD, United Kingdom.

## Abstract

**Objectives:** This study aimed to assess the impact of the COVID-19 lockdown on the screening and diagnosis of breast, colorectal, lung, and prostate cancer. The study also investigated whether the rates returned to pre-pandemic levels by December 2021.

**Design:** Cohort study.

**Setting:** Electronic health records from UK primary care Clinical Practice Research Datalink (CPRD) GOLD database.

**Participants:** The study included individuals registered with CPRD GOLD between January 2017 and December 2021, with at least 365 days of prior observation.

**Main outcome measures:** The study focused on screening, diagnostic tests, referrals and diagnoses of first-ever breast, colorectal, lung, and prostate cancer. Incidence rates (IR) were stratified by age, sex and region, and incidence rate ratios (IRR) were calculated to compare rates during and after lockdown with the reference period before lockdown. Forecasted rates were estimated using negative binomial regression models.

**Results:** Among 5,191,650 eligible participants, the initial lockdown resulted in reduced screening and diagnostic tests for all cancers, which remained dramatically reduced across the whole observation period for almost all tests investigated. For cancer incidence rates, there were significant IRR reductions in breast (0.69), colorectal (0.74), and prostate (0.71) cancers. However, the reduction in lung cancer incidence (0.92) was non-significant. Extrapolating to the entire UK population, an estimated 18,000 breast, 13,000 colorectal, 10,000 lung, and 21,000 prostate cancer diagnoses were missed from March 2020 to December 2021.

**Conclusion:** The national COVID-19 lockdown in the UK had a substantial impact on cancer screening, diagnostic tests, referrals and diagnoses. Although incidence rates started to recover after the lockdown, they remained significantly lower than pre-pandemic levels for breast and prostate cancers and associated tests. Delays in diagnosis are likely to have adverse consequences on cancer stage, treatment initiation, mortality rates, and years of life lost. Urgent strategies are needed to identify undiagnosed cases and address the long-term implications of delayed diagnoses.

**WHAT IS ALREADY KNOWN ON THIS TOPIC:**

- Breast, colorectal, lung, and prostate cancer are the most common causes of cancer death in the UK.
- The COVID-19 pandemic led to the postponement of cancer screening programs and reductions in diagnostic tests, resulting in delays in diagnosis and treatment initiation, impacting prognosis and mortality rates.
- Comprehensive data on the impact of changing social restrictions and post-lockdown periods is lacking in the UK, along with an assessment of specific screening pathways and patient experiences within the healthcare system.

**WHAT THIS STUDY ADDS:**

- The first UK national COVID-19 lockdown resulted in reductions in screening, diagnostic tests, and referrals, particularly for mammograms, colonoscopies, and visits to breast surgeons, leading to underdiagnosis of breast, colorectal, and prostate cancers. Despite some increase in rates after the lockdown, they remained significantly lower than pre-pandemic levels by December 2021, particularly for prostate cancer.
- Most affected populations were women aged 60-79 years for breast and colorectal cancer; men aged 60-79 years for lung cancer; and men aged 40-59 years for prostate cancer.
- Delays in diagnosis are likely to have consequences on cancer stage at diagnosis, treatment initiation, mortality rates, and total years of life lost. Strategies such as public awareness campaigns, targeted screening programs, and improved coordination between primary care and hospitals are needed to address the backlog and identify the potential ∼62,000 missed cancer cases in the UK.

## INTRODUCTION

Breast, colorectal, lung and prostate cancer are the four most common causes of cancer death in the United Kingdom (UK) [1]. Population screening programs (e.g. mammograms for breast cancer; faecal immunochemical tests (FIT) for colorectal cancer) aid early diagnosis, leading to better outcomes and prognosis [2]. However, due to the COVID-19 pandemic, and the first UK national lockdown (23rd March 2020), many health systems postponed cancer screening and diagnostic tests, to reduce spread of infection, and deployed staff towards critical COVID-19 patient care. ‘Stay at home’ advice, fear of contracting COVID-19, and social distancing measures introduced during the pandemic may also have altered health-seeking behaviour [3]. Combined, these changes in clinical practice and patient behaviour resulted in delays in diagnosis and treatment initiation, impacting on prognosis, mortality rates and total years of life lost [4].

Data suggest that countries responded to the COVID-19 pandemic differently. A review of studies from various countries showed significant declines in breast and lung cancer screenings and diagnostic biopsies during the pandemic [4]. In the USA, breast cancer screenings remained below expected levels even after one year [5]. However, Canada saw a return to pre-pandemic screening levels for breast, cervical, and colorectal cancer by 2021 [6]. In the UK, urgent cancer referrals initially dropped by up to 80%, with routine referrals reduced as patients delayed appointments [7, 8]. Referral rates for breast cancer mostly recovered by August 2020 and remained stable during subsequent lockdowns [9]. It remains unclear if this trend applies to other tests and cancer types. Reduced screening and referrals led to a decline in cancer rates globally [10–31]. However, there is a lack of studies estimating cancer incidence specifically in the UK during the pandemic and post-lockdown periods.

Using data from routinely recorded primary care electronic health records, the present study aims to 1) examine the frequencies and incidence rates (IR) of all consultations, cancer screening/ diagnostic tests/ referrals and breast, colorectal, lung and prostate cancer diagnoses in the general population before (from January 2017 to February 2020), during (March 2020 to June 2020) and after (July 2020 to December 2021) the first lockdown; 2) characterise newly diagnosed cancer patients in terms of frequencies of all consultations, procedures, measurements, comorbidities and medication use before, during and after lockdown; and 3) use time-series analyses to model the discrepancy between the observed and expected cancer diagnosis rates using data from 3 years prior to the pandemic to estimate how many cancer diagnoses may have been missed due to the pandemic, and whether diagnosis rates have stabilised to pre-pandemic levels. We focus on these four cancers as they are the most common and those where we have rapid diagnostics/screening tools available in the UK.

## METHODS

### Study participants

This study is a population-based cohort study using routinely collected electronic health records from UK Clinical Practice Research Datalink (CPRD) GOLD. CPRD GOLD contains anonymised patient-level information on demographics, lifestyle data, clinical diagnoses, prescriptions and preventive care contributed by general practitioners (GP) from the UK [32]. The use of CPRD data for this study was approved by the Independent Scientific Advisory Committee (22_002331). This database has previously been mapped to the Observational Medical Outcomes Partnership (OMOP) Common Data Model (CDM) [33]. People were eligible if they were registered between January 2017 and December 2021 with at least one year of prior clinical history. Additional criteria for the incident cancer diagnosis cohorts were including individuals who had a diagnosis or record of cancer, specifically for breast, colorectal, lung, or prostate cancer; excluding individuals diagnosed with the same type of cancer at any point in their clinical history and excluding those with metastases.

### Exposure

The ‘exposure’ was the date of the first UK national lockdown (23^rd^ March 2020), which was used to dissect the full study period into three distinct time-periods: pre-pandemic (January 2017 to February 2020), during lockdown (March 2020 to June 2020), and post-lockdown (July 2020 to December 2021). Additionally, we further dissected the extended post-lockdown periods distinguished by the changing social restrictions as shown in **Figure 1**.

**Figure 1.**
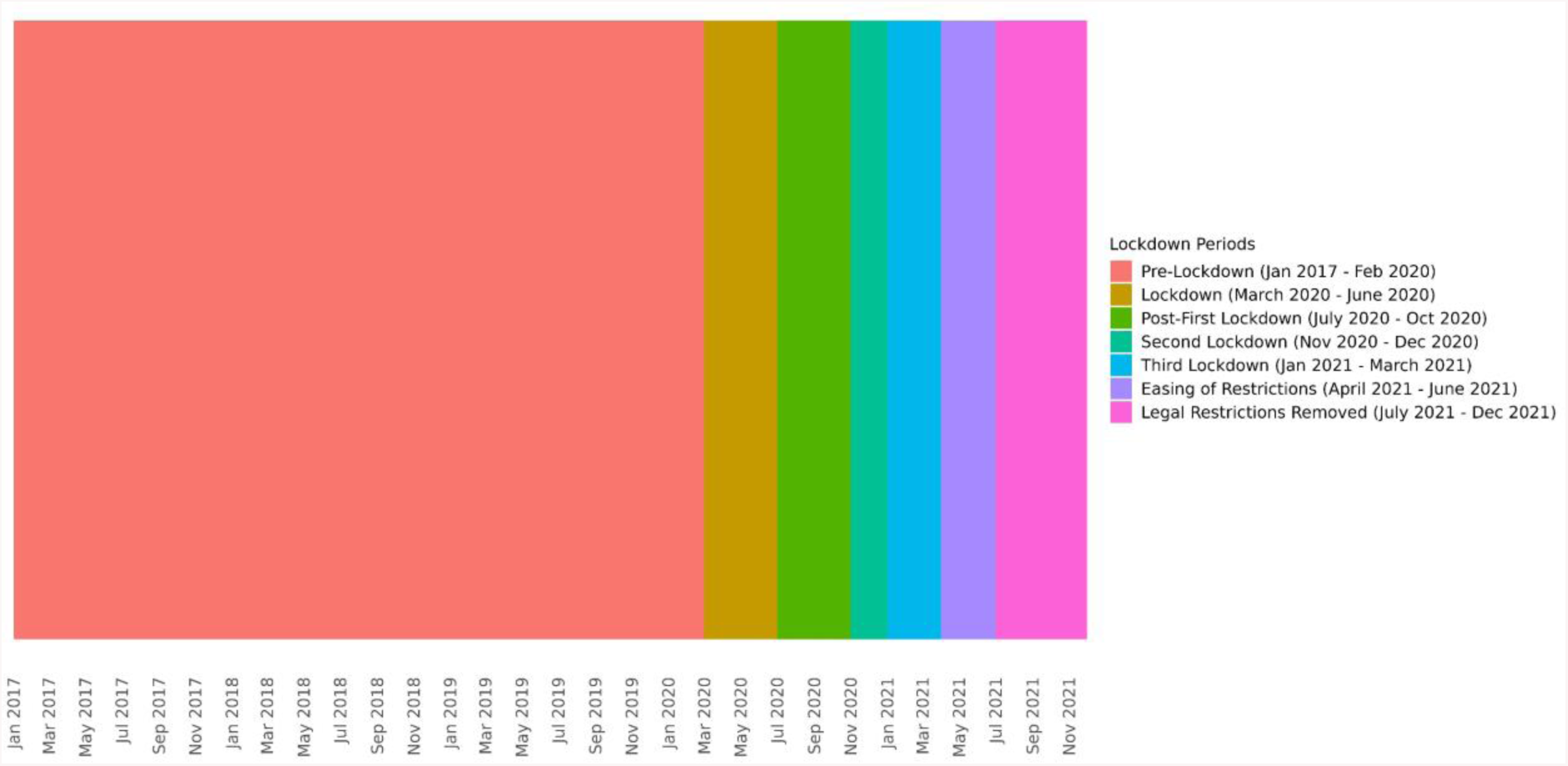
Dates of the observation period, dissected into periods distinguished by the changing social restrictions across the COVID-19 pandemic in the UK.

### Outcomes

For aim 1, frequencies of screening/ diagnostic tests and referrals relevant to each cancer were selected: they constitute the primary tools used in the cancer diagnostic pathways in the UK (see **Supplementary Table S1**). For aim 1 and 3, IR of cancer diagnoses included first-ever (incident) diagnoses of breast, colorectal, lung and prostate cancer. For aim 2, cancer patients were characterised on all comorbidities and medication usage available within CPRD GOLD across the study periods. All diagnoses, observations, measurements, procedures and medications were defined based on SNOMED/ Rx Norm/ LOINC codes (as appropriate), in the OMOP-mapped data. A list of all codes used to define each outcome can be found in our associated shiny app: https://dpa-pde-oxford.shinyapps.io/CancerCovid_CohortDiagnosticsShiny_paper1/

### Statistical analyses

#### Characterisations

Frequencies of screening, diagnostic tests, referrals, and interactions with the healthcare service, were calculated before, during and after the first lockdown in the general population and each cancer cohort. For cancer cohorts, counts were calculated in the 1-30 days; 31-180 days, and >180 days prior to index date (date of cancer diagnosis). Additionally, age at index date, sex, comorbidities, Charlson Comorbidity Index, Comorbidity Scale (CHADS2Vasc), Diabetes Complications and Severity Index (DCSI) and medication use were estimated for each cancer cohort using the FeatureExtraction R package [34]. Continuous variables were summarised as means and standard deviation or variances; and categorical variables as counts and percentages. Significant differences in these variables across time-periods were estimated using standardised mean difference (SMD). Where frequency counts were less than five, data were censored to further enhance patient/practice confidentiality.

#### Incidence Rates

Incidence rates (IR) with 95% confidence intervals (CI) were calculated for all outcomes and estimated annually, monthly, and within the pre-pandemic, lockdown, and extended post-lockdown periods across the entire study period (January 1^st^ 2017 to 1^st^ December 2021) using the IncidencePrevalence R package [35]. Patients who entered the database within this time (also referred to as the denominator population) contributed time-at-risk up to their first screening/ diagnostic test/ referral/ cancer diagnosis during the study period. Patients continued to contribute time-at-risk until the earliest of a record of screening/ diagnostic test/ referral/ cancer diagnosis, transfer out of the database, end of the study period or death. Incidence rate ratios (IRR) with 95% CI were calculated to examine differences in incidence of the lockdown and extended post-lockdown periods compared to the 3 years prior to the pandemic. IR were stratified by age (in 20-year age bands) sex, and region in the UK (England, Northern Ireland, Scotland and Wales). Sensitivity analysis focussing on prevalent cancer diagnoses (removing the requirement of the diagnosis being the first in the person’s history) was performed. Incidence rate ratios (IRR) were calculated using the IR estimates across the post-lockdown periods divided by the reference period before lockdown.

#### Time Series Analyses

Negative binomial logistic regression models were used to predict cancer IR each month since the beginning of the pandemic and to use these predictions to compare with observed IR. To validate our method, models were trained on data from January 2017 to February 2019 and used to forecast IR from March 2019 to March 2020. To account for seasonality, month was fitted as a categorical variable, and time (in number of months since the beginning of the study) was fitted as a continuous variable, as has been used previously for forecasting diagnoses over the pandemic [36, 37]. To validate our model fit, we examined whether the predicted versus observed counts and IR fell within 95% prediction intervals (PI). Results of our validation model can be visualised in **Supplementary Figures S1 and S2**). Using this approach, we trained the model using pre-pandemic data from January 2018 to February 2020 to forecast expected counts and IR from March 2020 onwards. Dates were chosen so that we had roughly equal number of months before vs. after lockdown. Number of ‘missing’ diagnoses were calculated as the difference between forecasted (observed) and expected number of incident cancer diagnoses during each time-period. The expected and observed counts were converted to IR by dividing the number of counts by the monthly observed person-month denominator population. The raw monthly cancer diagnosis counts were then extrapolated to the total population of the UK by multiplying the raw counts by a scalar representing the difference in population coverage of the CPRD database to the current UK population. All analyses were carried out using R Version 4.2.3.0

### Patient and public involvement

No patients or members of the public were involved in the design, analysis or interpretation of this study or the reported data because the study aims to examine population-level trends and patterns rather than individual experiences or perspectives.

## RESULTS

### Patient Characteristics

Overall, there were 5,191,650 people eligible to be included in the denominator population from January 2017. Total counts of patients excluded after applying the exclusion criteria for incidence estimates, are shown in Supplementary Table S2. The population structure of CPRD GOLD, in terms of age and sex was similar across the three time-periods (see Supplementary Tables S3-S4), but the proportion of practices from the different regions in the UK changed over time with fewer practices in England and greater proportion in Scotland during and after lockdown (Supplementary Figure S3). Demographics and total number of patients registered in CPRD in each of the time-periods, and diagnoses of first-ever breast, colorectal, lung and prostate cancer, are shown in Table 1. Mean age at date of diagnosis and sex distribution of cancer patients were largely the same across the three lockdown periods. Interactions with the healthcare system and routes to diagnosis were substantially reduced for patients receiving their cancer diagnosis during lockdown compared to those diagnosed pre-pandemic. Patients diagnosed after lockdown had fewer interactions with the healthcare system than pre-pandemic, though to a lesser extent than those diagnosed during lockdown (see Supplementary Tables S4-8). Across the lockdown periods there were no notable differences in comorbidities and medication prescriptions for those diagnosed with breast, colorectal, lung or prostate cancer (see Supplementary Tables S9-S12).

**Table 1.**
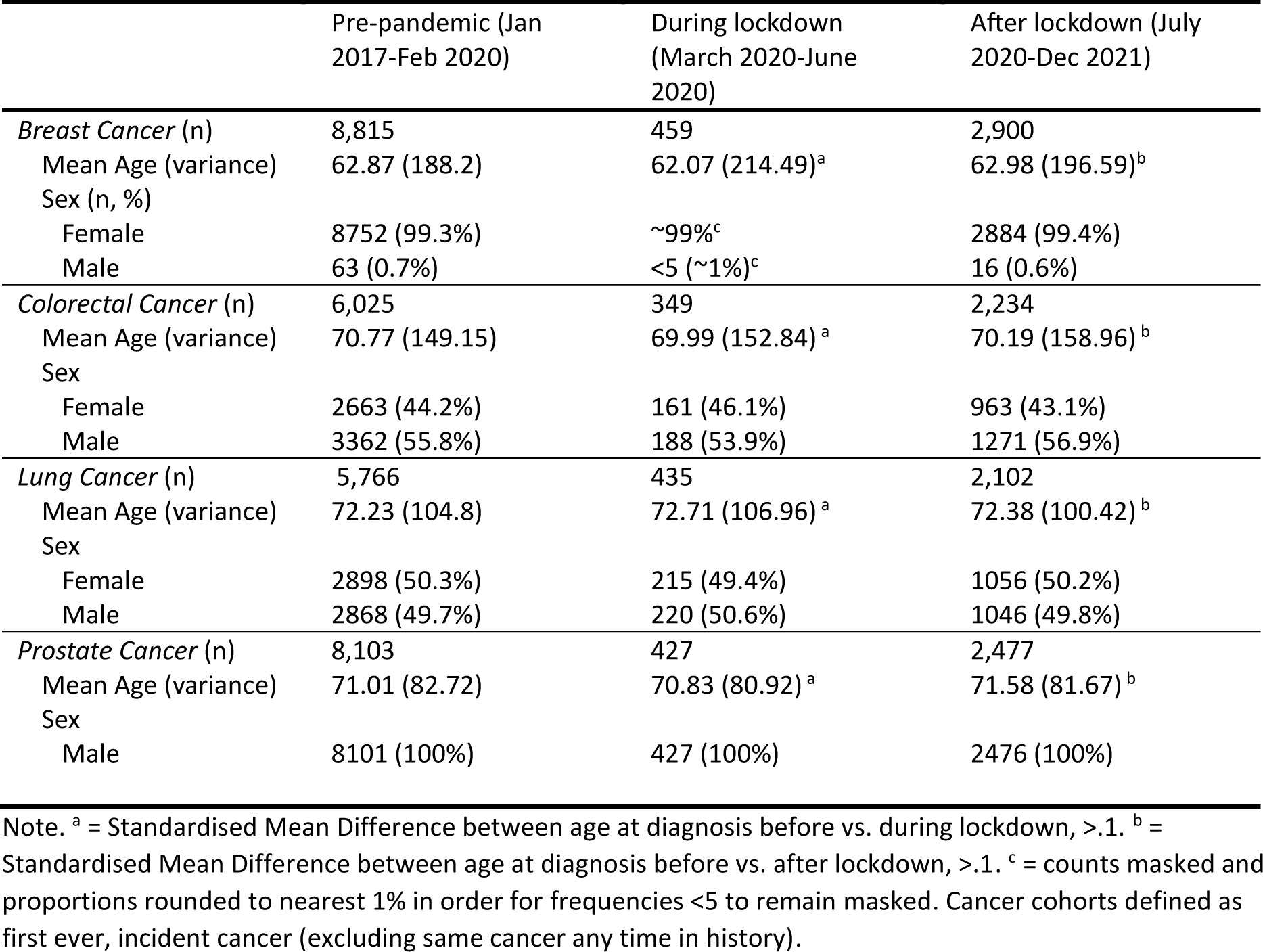
Counts, age and sex distribution in cancer cohorts in CPRD GOLD diagnosed before, during and after lockdown.

|  | Pre-pandemic (Jan 2017-Feb 2020) | During lockdown (March 2020-June 2020) | After lockdown (July 2020-Dec 2021) |
| --- | --- | --- | --- |
| *Breast Cancer (n)* | 8,815 | 459 | 2,900 |
| Mean Age (variance) | 62.87 (188.2) | 62.07 (214.49)<sup>a</sup> | 62.98 (196.59)<sup>b</sup> |
| Sex (n, %) |  |  |  |
| Female | 8752 (99.3%) | ~99%<sup>c</sup> | 2884 (99.4%) |
| Male | 63 (0.7%) | <5 (~1%)<sup>c</sup> | 16 (0.6%) |
| *Colorectal Cancer (n)* | 6,025 | 349 | 2,234 |
| Mean Age (variance) | 70.77 (149.15) | 69.99 (152.84)<sup>a</sup> | 70.19 (158.96)<sup>b</sup> |
| Sex |  |  |  |
| Female | 2663 (44.2%) | 161 (46.1%) | 963 (43.1%) |
| Male | 3362 (55.8%) | 188 (53.9%) | 1271 (56.9%) |
| *Lung Cancer (n)* | 5,766 | 435 | 2,102 |
| Mean Age (variance) | 72.23 (104.8) | 72.71 (106.96)<sup>a</sup> | 72.38 (100.42)<sup>b</sup> |
| Sex |  |  |  |
| Female | 2898 (50.3%) | 215 (49.4%) | 1056 (50.2%) |
| Male | 2868 (49.7%) | 220 (50.6%) | 1046 (49.8%) |
| *Prostate Cancer (n)* | 8,103 | 427 | 2,477 |
| Mean Age (variance) | 71.01 (82.72) | 70.83 (80.92)<sup>a</sup> | 71.58 (81.67)<sup>b</sup> |
| Sex |  |  |  |
| Male | 8101 (100%) | 427 (100%) | 2476 (100%) |

Note. <sup>a</sup> = Standardised Mean Difference between age at diagnosis before vs. during lockdown, >.1. <sup>b</sup> = Standardised Mean Difference between age at diagnosis before vs. after lockdown, >.1. <sup>c</sup> = counts masked and proportions rounded to nearest 1% in order for frequencies <5 to remain masked. Cancer cohorts defined as first ever, incident cancer (excluding same cancer any time in history).

### Incidence

#### Incidence of Screening/ diagnostic tests/ referrals across lockdown periods

**Figure 2** (and **Supplementary Table S14)** shows incidence rate ratios (IRR) of screening/ diagnostic tests/ referrals during the lockdown and extended post-lockdown periods compared to pre-pandemic rates. The number of routinely performed screening and diagnostic tests reduced during the first lockdown. Whilst rates of some screening/diagnostic tests increased across the extended post-lockdown periods (e.g. biopsy of breast IRR ranged from 0.76-1.35; and biopsy of prostate IRR ranged from 0.68-1.08), rates remained below those observed during the pre-pandemic era across nearly all extended post-lockdown periods, particularly so for colonoscopies (IRR ranged from 0.35-0.84), mammograms (IRR ranged from 0.23-0.98), and visits to breast surgeons (IRR ranged from 0.26-0.37). IR (per 100,000 person months or years as appropriate) of screening/ diagnostic tests and referrals are shown in **Supplementary Table S13, and Supplementary Figures S4-S7**.

**Figure 2.**
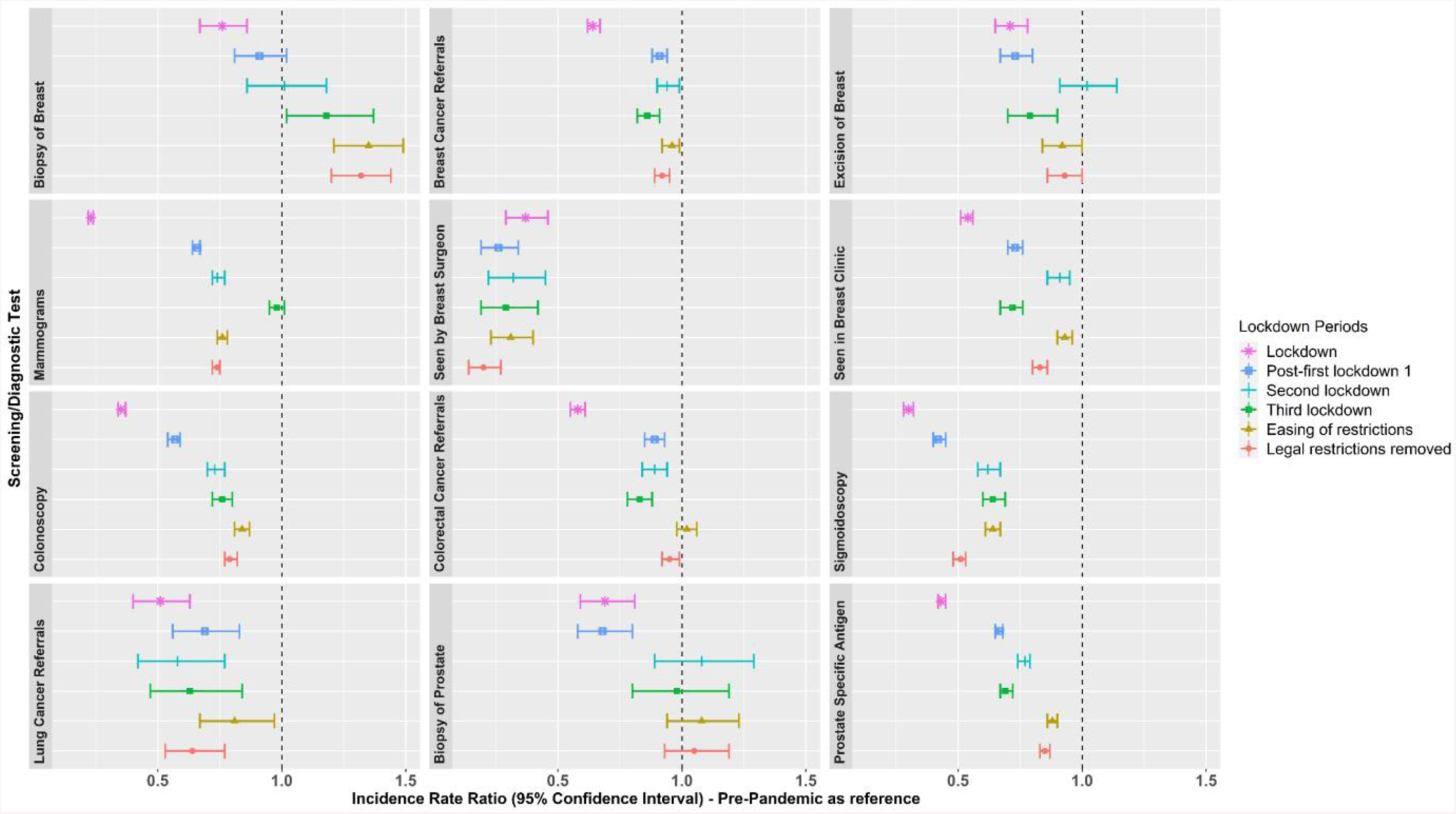
Incidence Rate Ratios of screening/ diagnostic tests and referrals in the extended post-lockdown periods compared to pre-pandemic rates. Note. Lockdown periods defined as: Lockdown (March 2020 to June 2020); post-first lockdown (July 2020 to October 2020); second lockdown (Nov 2020 to Dec 2020); third lockdown (Jan 2021 to March 2021); easing of restrictions (April 2021 to June 2021); and most legal restrictions removed (July 2021 to December 2021).

#### Incidence of breast, colorectal, lung and prostate cancer across different lockdown periods

**Figure 3** shows incidence rate ratios of the cancer diagnoses during the lockdown and extended post-lockdown periods compared to pre-pandemic rates. Diagnosis rates reduced during the initial lockdown for breast (IRR: 0.69 [95% CI: 0.63-0.74]), colorectal (IRR: 0.74 [95% CI: 0.67-0.81]); and prostate cancer (IRR: 0.71 [95% CI: 0.66-0.78]); but not for lung cancer (IRR: 0.92 [95% CI: 0.84-1.01]) (see **Supplementary Table S15** for full results). Whilst diagnosis rates started to increase across the extended post-first lockdown periods (ranging from 0.72-1.09), particularly during the second lockdown onwards for breast, colorectal and lung cancer, rates remained lower than the pre-pandemic era once legal restrictions were removed for breast (IRR: 0.93 [95% CI: 0.87-0.99]) and prostate cancer (IRR: 0.80 [95% CI: 0.74-0.86]).

**Figure 3.**
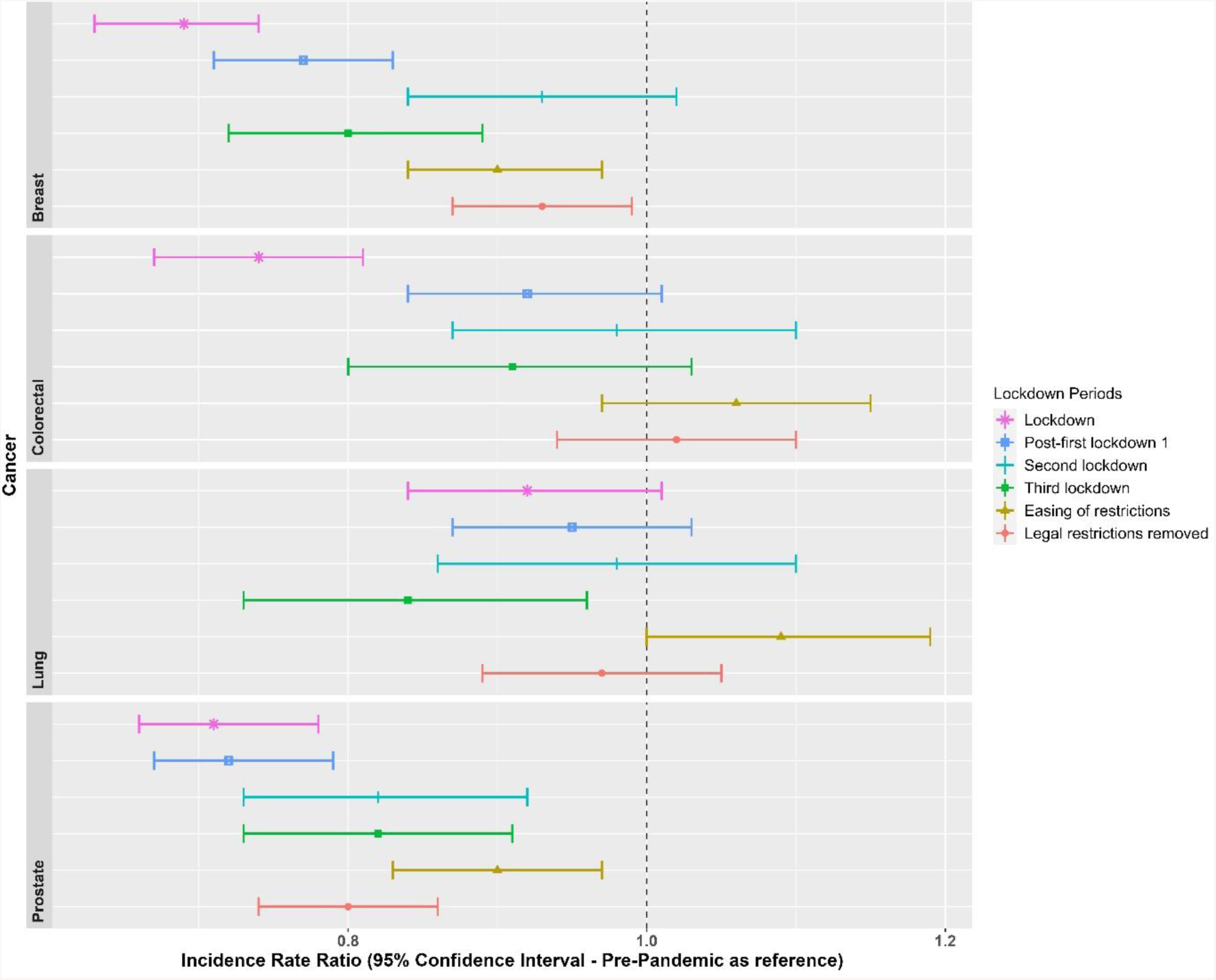
Incidence Rate Ratios for the cancer diagnoses, in each lockdown period, with the pre-pandemic era (Jan 2017 to Feb 2020) as a reference. Note. Lockdown periods defined as: Lockdown (March 2020 to June 2020); post-first lockdown (July 2020 to October 2020); second lockdown (Nov 2020 to Dec 2020); third lockdown (Jan 2021 to March 2021); easing of restrictions (April 2021 to June 2021); and most legal restrictions removed (July 2021 to December 2021).

IR and IRR of cancer diagnoses overall and stratified by age and sex are included in **Supplementary Figures S8-S10** and **Supplementary Tables S16-S23**. During the first lockdown, women aged 60-79 years were significantly underdiagnosed with breast cancer (IRR 0.65) compared to pre-pandemic levels, which improved once legal restrictions were lifted (IRR 0.9). The same age group was consistently underdiagnosed with colorectal cancer (IRR 0.66 during the first lockdown; IRR 0.6 during the third lockdown). Among men, those aged 80-150 years were most underdiagnosed with lung cancer during the third lockdown (IRR 0.66), while men aged 60-79 years consistently experienced underdiagnosis of lung cancer (IRR 0.84 during lockdown; 0.88 post-first lockdown).

Men aged 40-59 years were consistently underdiagnosed with prostate cancer (IRR ranging from 0.49 to 0.83) across different lockdown periods. IR stratified by region across three lockdown periods showed slightly smaller IR in England for breast cancer post-lockdown, colorectal, lung and prostate cancer pre-pandemic, and prostate cancer post-lockdown, compared to the other UK regions (**Supplementary Figure 11**).

##### Forecasting expected cancer diagnosis rates after lockdown

The forecasted cancer diagnosis rates after the lockdown were estimated using negative binomial regression models based on pre-pandemic data. Durbin-Watson statistics for all cancers were between 1.12-1.72, and plots of residuals show autocorrelation only for 22.5% of timepoints. **Figure 4** shows that breast cancer incidence rates were significantly below expected levels for six months after the lockdown, and prostate cancer rates remained below expectations for a year (where points fall outside of the PI). The observed diagnosis rates during the first lockdown were much lower than expected for all four cancers, ranging from 15.4% to 33.9% reductions (**Supplementary Table S24)**. Although the proportion of potential underdiagnoses decreased over time, diagnosis rates remained lower than expected in the last 2 months of follow-up for all bar breast cancer. Overall, the model estimated around 18,000 missed breast cancer diagnoses, 13,000 for colorectal cancer, 10,000 for lung cancer, and 21,000 for prostate cancer across the UK population from March 2020 to December 2021 (see **Supplementary Table S25**).

**Figure 4.**
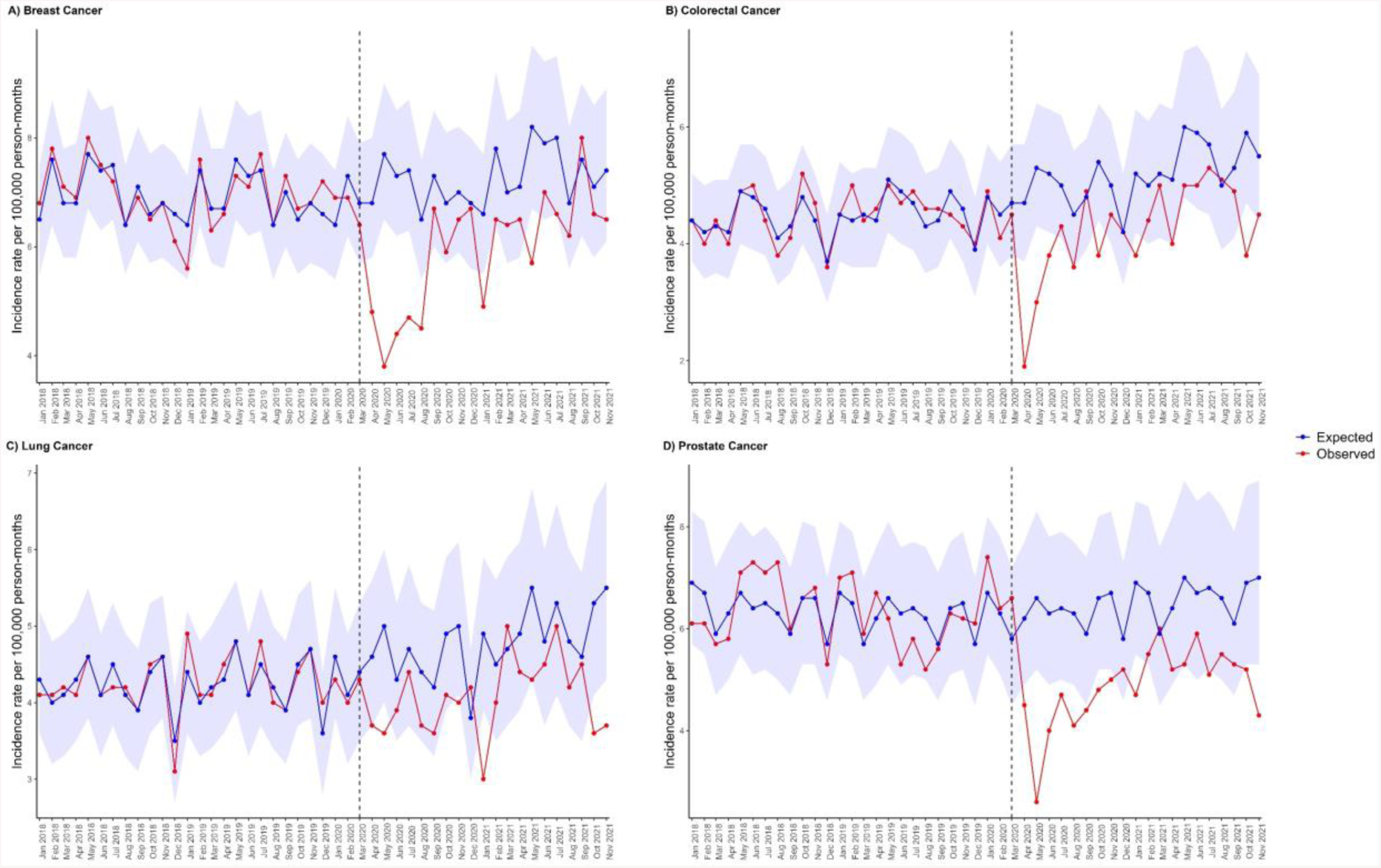
Expected and observed IR per 100,000 person months of A) breast cancer, B) colorectal cancer, C) lung cancer, and D) prostate cancer in primary care records from CPRD GOLD UK. Points represent monthly IR. Expected rates (with 95% prediction intervals represented by the shaded areas) were calculated using negative binomial regression using observed data from January 2017 to February 2020 to estimate expected counts from March 2020 to November 2021. The vertical line indicates the start of lockdown in March 2020.

When stratifying by age and sex, during the first lockdown women aged 60-79 years were most underdiagnosed for breast cancer (37.4%) and colorectal cancer (45.8%); whereas men aged 60-79 years were most underdiagnosed for lung cancer (31%); and men aged 40-59 years were most underdiagnosed for prostate cancer (35.3%).

Across the total observation period from March 2020 to December 2021, women aged 20 to 39 years had the greatest proportion of underdiagnoses of breast cancer (39.1%) (**Supplementary Table S30**). The greatest absolute number of potential missed breast cancer cases was for women aged 60 to 79 years (n=498, or n=10,751 extrapolated to the whole UK population). Women aged 60 to 79 years had the greatest proportion of underdiagnosed colorectal cancer (29.3%), with 260 estimated missed colorectal cancer cases (or n= 5,613 extrapolated to the whole UK population). For lung cancer, men aged 60 to 79 years had the greatest proportion of potential underdiagnoses (26.5%), reflecting potentially 310 missed lung cancer cases (or n= 6,693 extrapolated to the whole UK population). For prostate cancer, men aged 40 to 59 years suffered the greatest proportion of potential underdiagnoses (26.8%), reflecting potentially 104 missed prostate cancer cases (or n= 2,245 extrapolated to the whole UK population).

## DISCUSSION

### Statement of principal findings

The findings of this study revealed a reduction in number of routinely performed screening, diagnostic tests, and referrals during the period from March 2020 to December 2021 compared to data from January 2017 to February 2020. Particularly during the first lockdown, there was a substantial decrease in mammograms, sigmoidoscopies, colonoscopies, and visits to breast surgeons by 77%, 70%, 65%, and 63% respectively, compared to pre-pandemic rates. Similar findings were reported in other countries, such as Slovenia [10] and Argentina [38].

Although some rates of screening, diagnostic tests, and referrals increased in the post-first lockdown period, they remained below pre-pandemic levels. For instance, mammograms, colonoscopies, and visits to breast surgeons were still reduced by 26%, 21%, and 80% respectively between July 2021 and December 2021. These findings contradict studies from Catalonia, Spain, and Canada, where mammograms and colonoscopies returned to expected levels by December 2021 [17, 30]. Similarly, data from Canada shows that breast cancer screening returned to pre-pandemic levels by December 2020; and faecal occult blood tests for colorectal cancer by September 2020 [6]. In the UK, the data from CPRD GOLD did not show a recovery to pre-pandemic levels for screening and diagnostic tests. Possible explanations include the fact that the UK was the only European country to have additional lockdowns after the first, and that the NHS has experienced staff shortages and strikes over recent years impacting on its capacity to catch up.

Lockdown had varying effects on screening and referral procedures compared to diagnostic procedures in CPRD GOLD. Diagnostic procedures were not deprioritized after lockdown, except for visits to breast surgeons, indicating efforts to reduce the backlog. Screening, on the other hand, was more susceptible to postponement or lower prioritization, as it is used for asymptomatic individuals, as shown in the data.

### Research in Context

There are multiple explanations for the persistent reductions in screening, diagnostic tests, and referrals during extended lockdown periods. Variations in screening reductions may be related to the prevalence of COVID-19 restrictions/infections across countries [4]. Reports indicate that the UK’s response to the pandemic was inadequate, resulting in a significant impact on the country and the need for subsequent lockdowns. The UK faced high infection rates, hospitalizations, and a substantial death toll [39]. Healthcare resources were diverted from standard care, affecting cancer diagnostic pathways until December 2021. Although rates showed some increase from March 2020 to December 2021, they were inconsistent. Data from a systematic review predicted a clearance of the screening backlog (specifically mammograms) in the USA within 12-24 weeks [4, 40], whereas our data suggest that even after 52-73 weeks, the queue was not cleared in the UK. That said, changes in screening methods (such as a switch from direct appointments to open invitations for routine mammograms) may have affected number of patients screened at least for breast cancer across this time period [41, 42].

Reduced screening and diagnostic tests lead to decreased cancer detection and diagnosis. Breast, colorectal, and prostate cancers were significantly underdiagnosed during lockdown and remained below expected levels until June 2021 for breast cancer and until December 2021 for prostate cancer. The expected effect of these reductions in rates is that diagnoses will be delayed, and prognosis worsened by these backlogs in diagnosis and treatment. These findings contradict a study from Catalonia, Spain, where breast cancer diagnoses recovered to pre-pandemic levels within this time-frame [17, 30]. Belgium’s cancer registry data also showed recovery by June 2020. Although incidence rates for colorectal and lung cancer returned to pre-pandemic levels, these rates likely represent missed diagnoses during lockdown, requiring substantial catch-up to compensate for the shortfall.

Our model predicts that prostate cancer had the highest number of missed cases, with an estimated 21,525 (25%) of expected cases missed from March 2020 to December 2021. Similar reductions in prostate cancer diagnosis were observed in other studies [17]. Lung cancer was the least affected, with 14.2% of expected cases missed, which aligns with other reports [17, 30]. It is possible this is because we have limited screening tools for lung cancer, leading to comparatively smaller diagnosis rates compared to other cancers. Though the increased use of chest radiography during COVID-19 infections may have inadvertently led to the identification of potential lung cancer symptoms and subsequent diagnosis [17]. Stratification analyses revealed consistent underdiagnosis in specific age groups: women aged 60 to 79 for breast and colorectal cancer, men aged 60 to 79 for lung cancer, and men aged 40 to 59 years for prostate cancer. It is already known that we see a steep rise in risk for these cancers from these ages onwards [43, 44]. These findings emphasize the urgency of prioritizing screening and diagnostics in these populations to detect the missed cases.

### Strengths of the Study

This study benefits from the strengths of CPRD GOLD, known for its extensive UK population coverage and comprehensive healthcare records [32], facilitating thorough phenotyping of screening, diagnostics, and cancer cases. The longitudinal nature of the database enabled an extended observation period beyond the typical one-year post-lockdown timeframe. Unlike most studies, our analysis covers screening and diagnostic rates up to December 2021. Further research should explore additional data to assess if the UK has fully recovered from the rate shortfalls.

### Weaknesses of the Study

Although this study has many strengths it does have some limitations. First, as these data are derived from primary care and not linked to cancer registry data there were many screening and diagnostic tests of relevance to this study that were not captured in the database. This is common of studies using primary care data, as many diagnostic tests and procedures occur in hospital settings. Furthermore, cancer diagnoses may have shifted to hospital settings during the pandemic, and there may be a time-lag in recording cancer diagnoses in primary care records. Thus, it is likely that the estimated shortfall in screening/diagnostic tests, and cancer diagnosis rates in the present study, are underestimated. Second, the composition of patients and practices in the database have changed over time. Indeed, with the advent of the CPRD AURUM database, some practices were transferred out of GOLD and into AURUM, thus accounting for the reduced source population counts across time-points. Reassuringly, the IR of the cancers in the three broad time-periods across regions were largely similar, except for slightly smaller IR in England across some time-points, likely reflecting the change in population composition. Thirdly, the generalizability of findings is predominantly limited to Scotland and Wales, with less representation from England and Northern Ireland. Finally, as real-world evidence, causal inference is challenging, and other factors could have influenced the reduction in cancer diagnoses during lockdown, such as pre-existing trends in screening/diagnostic tests and cancer diagnoses, seasonality patterns, or COVID-19-related deaths. Whilst we did observe a pre-existing downward trend for visits to breast surgeons, there were no other observed pre-existing trends in our data, and our modelling statistically accounted for seasonal variability. However, data from Catalonia suggest a small proportion of missed diagnoses were attributed to COVID-19 deaths [45].

### Implications for Clinicians and Policymakers

Delays in diagnosis are likely to impact on cancer stage at time of diagnosis, treatment initiation, mortality rates and total years of life lost. To effectively tackle the existing backlog and potential long-term consequences on cancer survival, it may be necessary to implement strategies to identify those potential ∼62,000 cancer cases missed. These could include raising public awareness through targeted campaigns aimed at particular age groups that have been most affected, encouraging participation in screening programs, and enhancing the coordination between primary care facilities and hospitals. Increases in screening and diagnostic testing may need to be increased in the months following December 2021 to account for the observed shortfall in the UK. These measures are vital for effective public health intervention and reducing the impact of delayed diagnoses on cancer outcomes.

## Data Availability

Patient level data cannot be shared without approval from data custodians owing to local information governance and data protection regulations. Aggregated data, analytical code, and detailed definitions of algorithms for identifying the events are available in GitHub repositories (https://github.com/oxford-pharmacoepi/CancerCovid_CohortDiagnostics; https://github.com/oxford-pharmacoepi/CancerCovid_Characterisations; https://github.com/oxford-pharmacoepi/CancerCovid_IncidencePrevalence; https://github.com/oxford-pharmacoepi/CancerCovid_NegativeBinomialReg)

https://github.com/oxford-pharmacoepi/CancerCovid_Characterisations;

https://github.com/oxford-pharmacoepi/CancerCovid_IncidencePrevalence

https://github.com/oxford-pharmacoepi/CancerCovid_NegativeBinomialReg

## COMPETING INTERESTS

All authors have completed the ICMJE disclosure form at http://www.icmje.org/disclosure-of-interest/ and declare the following interests: DPA’s research group has received research grants from the European Medicines Agency; the Innovative Medicines Initiative; Amgen, Chiesi, and UCB Biopharma; and consultancy or speaker fees from Astellas, Amgen, and UCB Biopharma.

## ROLE OF THE FUNDING SOURCE

This research was partially funded by the European Health Data and Evidence Network (EHDEN) (grant number 806968), the Optimal treatment for patients with solid tumours in Europe through Artificial Intelligence (OPTIMA) initiative, and the Oxford NIHR Biomedical Research Centre. OPTIMA is funded through the IMI2 Joint Undertaking and is listed under grant agreement No. 101034347.

IMI2 receives support from the European Union’s Horizon 2020 research and innovation programme and the European Federation of Pharmaceutical Industries and Associations (EFPIA). IMI supports collaborative research projects and builds networks of industrial and academic experts in order to boost pharmaceutical innovation in Europe. The views communicated within are those of OPTIMA. Neither the IMI nor the European Union, EFPIA, or any Associated Partners are responsible for any use that may be made of the information contained herein. The study funders had no role in the conceptualisation, design, data collection, analysis, decision to publish, or preparation of the manuscript. DPA receives funding from the UK National Institute for Health Research (NIHR) in the form of a senior research fellowship and the Oxford NIHR Biomedical Research Centre.

## CONTRIBUTOR STATEMENT

MPM, MCS, AJ, DN and DPA conceived the study. MPM, MCS, DPA and NB were involved in developing the study design. NB, MCS, MPM and BR wrote and executed the statistical code. AD and WYM mapped the CPRD GOLD data to the OMOP CDM. NB wrote the manuscript. All authors interpreted the results. All authors were involved in the editing of drafts of the manuscript. All authors approved the final version and had final responsibility for the decision to submit for publication. DPA is the guarantor. The corresponding author attests that all listed authors meet authorship criteria and that no others meeting the criteria have been omitted.

## ETHICS APPROVAL

The protocol for this research was approved by the independent scientific advisory committee for Medicine and Healthcare products Regulatory Agency database research (protocol number 22_002331).

## TRANSPARENCY STATEMENT

The lead author (and the manuscript’s guarantor) affirms that the manuscript is an honest, accurate, and transparent account of the study being reported; that no important aspects of the study have been omitted; and that any discrepancies from the study as originally planned (and, if relevant, registered) have been explained.

## Notes

### Competing Interest Statement

DPAs research group has received research grants from the European Medicines Agency; the Innovative Medicines Initiative, Amgen, Chiesi, and UCB Biopharma, and consultancy or speaker fees from Astellas, Amgen, and UCB Biopharma.

### Summary of Updates

Masking of counts <5 and rounding of masked proportions updated throughout.

## REFERENCES

1. Cancer Research UK. Twenty most common causes of cancer death [online]. 2022. [https://www.cancerresearchuk.org/health-professional/cancer-statistics/mortality/common-cancers-compared#heading-Zero] (accessed 12th Oct 2022).

2. Loud JT, Murphy J. Cancer screening and early detection in the 21st century. Seminars in Oncology Nursing. 2017;33:121–8.

3. Abraham SA, Agyare DF, Yeboa NK, et al. The influence of COVID-19 pandemic on the health seeking behaviors of adults living with chronic conditions: a view through the health belief model. J Prim Care Community Health. 2023; 14. doi: 10.1177/21501319231159459.

4. Alkatout I, Biebl M, Momenimovahed Z, et al. Has COVID-19 affected cancer screening programs? A systematic review. Front Oncol. 2021;11:675038.

5. Milgrom ZZ, Milgrom DP, Han Y, et al. Breast Cancer Screening, Diagnosis, and Surgery during the Pre- and Peri-pandemic: Experience of Patients in a Statewide Health Information Exchange. Ann Surg Oncol. 2023;30:2883–94.

6. Decker KM, Feely A, Bucher O, et al. Evaluating the impact of the COVID-19 pandemic on cancer screening in a central Canadian province. Prev Med. 2022;155:106961.

7. Bodkin H. Cancer referrals down by the 80 per cent in some areas as coronavirus fears keep patients from hospitals. The Telegraph. 2020. 15th April 2020.

8. Maringe C, Spicer J, Morris M, et al. The impact of the COVID-19 pandemic on cancer deaths due to delays in diagnosis in England, UK: a national, population-based, modelling study. Lancet Oncol. 2020;21:1023–34.

9. Gathani T, Dodwell D, Horgan K. The impact of the first 2 years of the COVID-19 pandemic on breast cancer diagnoses: a population-based study in England. Br J Cancer. 2022:1–3.

10. Zadnik V, Mihor A, Tomsic S, et al. Impact of COVID-19 on cancer diagnosis and management in Slovenia - preliminary results. Radiol Oncol. 2020;54:329–34.

11. Ward ZJ, Walbaum M, Walbaum B, et al. Estimating the impact of the COVID-19 pandemic on diagnosis and survival of five cancers in Chile from 2020 to 2030: a simulation-based analysis. Lancet Oncol. 2021;22:1427–37.

12. Vrdoljak E, Balja MP, Marušić Z, et al. COVID-19 Pandemic Effects on Breast Cancer Diagnosis in Croatia: A Population- and Registry-Based Study. Oncologist. 2021;26:e1156–e60.

13. Van Wyk AC, De Jager LJ, Razack R, et al. The initial impact of the COVID-19 pandemic on the diagnosis of new cancers at a large pathology laboratory in the public health sector, Western Cape Province, South Africa. S Afr Med J. 2021;111:570–4.

14. Uyl-de Groot CA, Schuurman MS, Huijgens PC, et al. Fewer cancer diagnoses during the COVID-19 epidemic according to diagnosis, age and region. TSG-Tijdschrift voor gezondheidswetenschappen. 2021;99:1–8.

15. Quarello P, Ferrari A, Mascarin M, et al. Diagnostic Delay in Adolescents with Cancer During COVID-19 Pandemic: A New Price for Our Patients to Pay. J Adolesc Young Adult Oncol. 2022;11:316–9.

16. Peacock HM, Tambuyzer T, Verdoodt F, et al. Decline and incomplete recovery in cancer diagnoses during the COVID-19 pandemic in Belgium: a year-long, population-level analysis. ESMO Open. 2021;6:100197.

17. Mora N, Guiriguet C, Cantenys R, et al. Cancer diagnosis in primary care after second pandemic year in Catalonia: a time-series analysis of primary care electronic health records covering about 5 million people. Fam Pract. 2023;40:183–7.

18. Longcroft-Wheaton G, Tolfree N, Gangi A, et al. Data from a large Western centre exploring the impact of COVID-19 pandemic on endoscopy services and cancer diagnosis. Frontline Gastroenterol. 2021;12:193–9.

19. Liu C, Piao H, Zhang T, et al. Delayed Diagnosis and Treatment of Cancer Patients During the COVID-19 Pandemic in Henan, China: An Interrupted Time Series Analysis. Front Public Health. 2022;10:881718.

20. Linck PA, Garnier C, Depetiteville MP, et al. Impact of the COVID-19 lockdown in France on the diagnosis and staging of breast cancers in a tertiary cancer centre. Eur Radiol. 2022;32:1644–51.

21. Knoll K, Reiser E, Leitner K, et al. The impact of COVID-19 pandemic on the rate of newly diagnosed gynecological and breast cancers: a tertiary center perspective. Arch Gynecol Obstet. 2022;305:945–53.

22. Kim HY, Kim MG, Kang MR, et al. No evidence of delay in colorectal cancer diagnosis during the COVID-19 pandemic in Gwangju and Jeonnam, unclosed areas in Korea. Epidemiol Health. 2022:e2022092.

23. Kempf E, Lamé G, Layese R, et al. New cancer cases at the time of SARS-Cov2 pandemic and related public health policies: A persistent and concerning decrease long after the end of the national lockdown. Eur J Cancer. 2021;150:260–7.

24. Kaltofen T, Hagemann F, Harbeck N, et al. Changes in gynecologic and breast cancer diagnoses during the first wave of the COVID-19 pandemic: analysis from a tertiary academic gyneco-oncological center in Germany. Arch Gynecol Obstet. 2022;305:713–8.

25. Jacob L, Kalder M, Kostev K. Decrease in the number of patients diagnosed with cancer during the COVID-19 pandemic in Germany. J Cancer Res Clin Oncol. 2022;148:3117–23.

26. Ferrara G, De Vincentiis L, Ambrosini-Spaltro A, et al. Cancer Diagnostic Delay in Northern and Central Italy During the 2020 Lockdown Due to the Coronavirus Disease 2019 Pandemic. Am J Clin Pathol. 2021;155:64–8.

27. Erdmann F, Wellbrock M, Trübenbach C, et al. Impact of the COVID-19 pandemic on incidence, time of diagnosis and delivery of healthcare among paediatric oncology patients in Germany in 2020: Evidence from the German Childhood Cancer Registry and a qualitative survey. Lancet Reg Health Eur. 2021;9:100188.

28. Dinmohamed AG, Visser O, Verhoeven RHA, et al. Fewer cancer diagnoses during the COVID-19 epidemic in the Netherlands. Lancet Oncol. 2020;21:750–1.

29. De Vincentiis L, Carr RA, Mariani MP, et al. Cancer diagnostic rates during the 2020 ‘lockdown’, due to COVID-19 pandemic, compared with the 2018-2019: an audit study from cellular pathology. J Clin Pathol. 2021;74:187–9.

30. Coma E, Guiriguet C, Mora N, et al. Impact of the COVID-19 pandemic and related control measures on cancer diagnosis in Catalonia: a time-series analysis of primary care electronic health records covering about five million people. BMJ Open. 2021;11:e047567.

31. Cantini L, Mentrasti G, Russo GL, et al. Evaluation of COVID-19 impact on DELAYing diagnostic-therapeutic pathways of lung cancer patients in Italy (COVID-DELAY study): fewer cases and higher stages from a real-world scenario. ESMO Open. 2022;7:100406.

32. Herrett E, Gallagher AM, Bhaskaran K, et al. Data resource profile: clinical practice research datalink (CPRD). Int J Epidemiol. 2015;44:827–36.

33. Voss EA, Makadia R, Matcho A, et al. Feasibility and utility of applications of the common data model to multiple, disparate observational health databases. J Am Med Inform Assoc. 2015;22:553–64.

34. Schuemie M, Suchard, M., Ryan, P., Reps, J., Sena, A. Feature Extraction. R Package Version 3.2.0 [online]. 2021. [https://ohdsi.github.io/FeatureExtraction/] (accessed 23rd June 2023).

35. Burn E, Raventos, B., Catala, M., Du, M., Guo, Y., Black, A>, Inberg, G., Lopez, K. IncidencePrevalence: Estimate Incidence and Prevalence using the OMOP Common Data Model. R Package Verion 0.4.0 [online]. 2023. [https://cran.r-project.org/web/packages/IncidencePrevalence/index.html] (accessed 23rd June 2023).

36. Raventós B, Pistillo A, Reyes C, et al. Impact of the COVID-19 pandemic on diagnoses of common mental health disorders in adults in Catalonia, Spain: a population-based cohort study. BMJ open. 2022;12:e057866.

37. Williams R, Jenkins DA, Ashcroft DM, et al. Diagnosis of physical and mental health conditions in primary care during the COVID-19 pandemic: a retrospective cohort study. The Lancet Public Health. 2020;5:e543–e50.

38. Bozovich GE, Alves De Lima A, Fosco M, et al. [Collateral damage of COVID-19 pandemic in private healthcare centres of Argentina]. Medicina (B Aires). 2020;80 Suppl 3:37–41.

39. World Health Organization (WHO). Coronavirus disease (COVID-19) pandemic [online]. 2020. [https://www.who.int/emergencies/diseases/novel-coronavirus-2019] (accessed 23rd June 2023).

40. Song H, Bergman A, Chen AT, et al. Disruptions in preventive care: Mammograms during the COVID-19 pandemic. Health services research. 2021;56:95–101.

41. Rossi PG, Giordano L. Mammography screening: please don’t be vague, tell me when I should come! The Lancet Oncology. 2017;18:848–9.

42. NHS. Breast Cancer Screening Recovery Services [online]. 2020. [https://selondonccg.nhs.uk/wp-content/uploads/2020/11/Breast-screening-new-tel-no.pdf] (accessed 26th June 2023).

43. Cancer Research UK. Prostate cancer incidence statistics [online]. 2021. [https://www.cancerresearchuk.org/health-professional/cancer-statistics/statistics-by-cancer-type/prostate-cancer/incidence#heading-One] (accessed 26th June 2023).

44. Cancer Research UK. Breast cancer incidence (invasive) statistics [online]. 2021. [https://www.cancerresearchuk.org/health-professional/cancer-statistics/statistics-by-cancer-type/breast-cancer/incidence-invasive#heading-One] (accessed 26th June 2023).

45. Pifarre IAH, Vidal-Alaball J, Gil J, et al. Missing Diagnoses during the COVID-19 Pandemic: A Year in Review. Int J Environ Res Public Health. 2021;18:5335.

