## Supplementary material for "The Impact of the UK COVID-19 Lockdown on the Screening, Diagnostics and Incidence of Breast, Colorectal, Lung and Prostate Cancer in the UK: a Population-Based Cohort Study"

Supplementary Tables and Figures

**Table S1. Screening and Diagnostic Tests Examined in Characterisations and Incidence Analyses**

| **Screening/ Diagnostic Test** | **Incidence Cohort** |
|---|---|
| *Breast* | |
| Referral to breast clinic | Breast cancer referrals |
| Referral to mammography clinic | Breast cancer referrals |
| Fast track referral for suspected breast cancer | Breast cancer referrals |
| Referral to breast surgeon | Breast cancer referrals |
| Screening mammograms | Mammograms |
| Seen in breast clinic | Seen in breast clinic |
| Diagnostic mammograms | Mammograms |
| Diagnostic mammograms with ultrasound | Mammograms |
| Biopsy of breast | Biopsy of Breast |
| Stereotactically guided needle biopsy of breast | Biopsy of Breast |
| Percutaneous needle biopsy of breast | Biopsy of Breast |
| Fine needle aspiration of breast | Biopsy of Breast |
| Wire guided local excision of breast | Biopsy of Breast |
| Excision of mammary duct | Biopsy of Breast |
| Wide local excision of breast | Biopsy of Breast |
| Excision of breast tissue | Biopsy of Breast |
| Seen by breast surgeon | Seen by breast surgeon |
| *Colorectal* | |
| Colorectal cancer screening programme | Colorectal cancer screening programme |
| Fast-track referral for suspected colorectal cancer | Colorectal cancer referrals |
| Quantitative faecal immunochemical test | / |
| Ultrasonography of intestine | / |
| Colonoscopy | Colonoscopy |
| Sigmoidoscopy | Sigmoidoscopy |
| Ultrasound of gastrointestinal tract | / |
| Ultrasonography of abdomen | / |
| Ultrasonography of rectum | / |
| *Lung* | |
| Fast track referral for suspected lung cancer | Lung cancer referrals |
| Bronchoscopy | Bronchoscopy |
| Endobronchial ultrasonography guided transbronchial needle aspiration | Diagnostic procedures of chest |
| Mediastinoscopy | Diagnostic procedures of chest |
| CT and biopsy of chest | Diagnostic procedures of chest |
| Ultrasound and biopsy of chest | Diagnostic procedures of chest |
| Diagnostic radiology procedures of chest | Diagnostic procedures of chest |
| *Prostate* | |
| Prostate specific antigen (PSA) test | Prostate specific antigen (PSA) test |
| PSA monitoring | / |
| Biopsy of prostate | Biopsy of prostate |

Note. / = Tests/measurements/procedures that did not have sufficient counts in the CPRD GOLD database were not included in the incidence analyses.

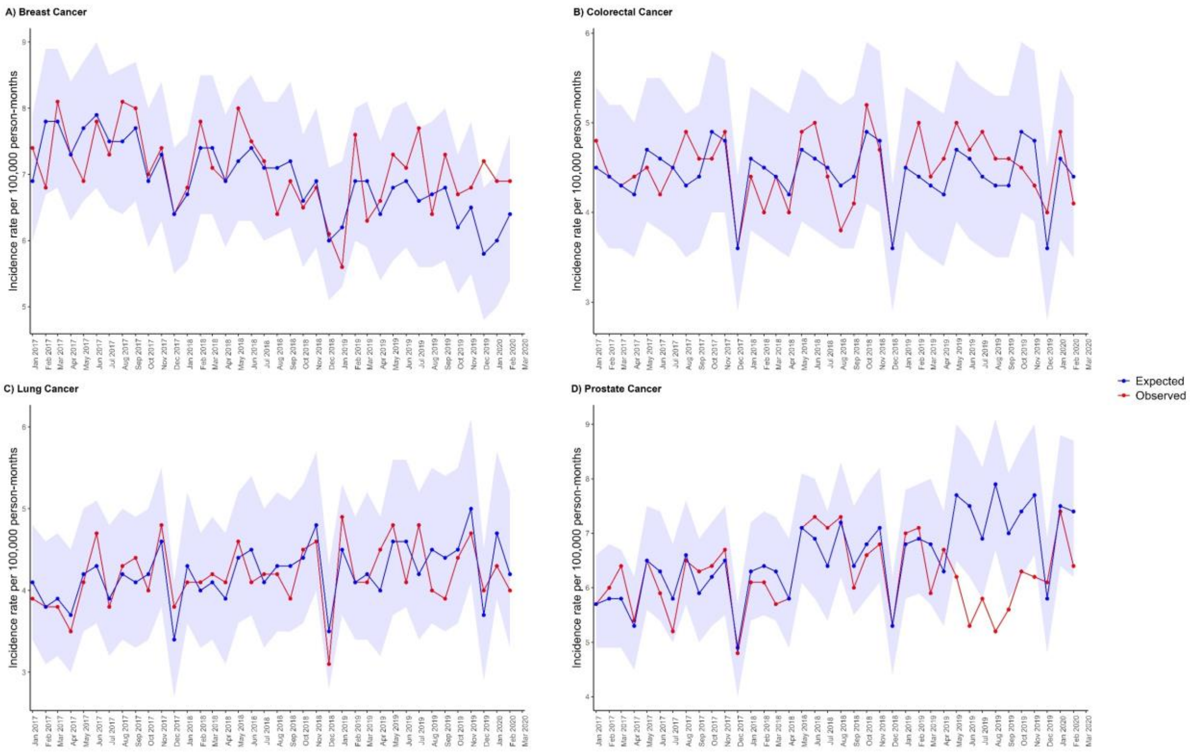


**Figure S1.** Expected and observed incidence rates per 100,000 person months of A) breast cancer, B) colorectal cancer, C) lung cancer, and D) prostate cancer in primary care records from CPRD UK, from the validation model. Note. Points represent monthly incidence rates. Expected rates (with 95% prediction intervals) were calculated using negative binomial regression using observed data from January 2017 to February 2020 to estimate expected counts during the same period to validate the methodology. The shaded areas represent the 95% prediction intervals.

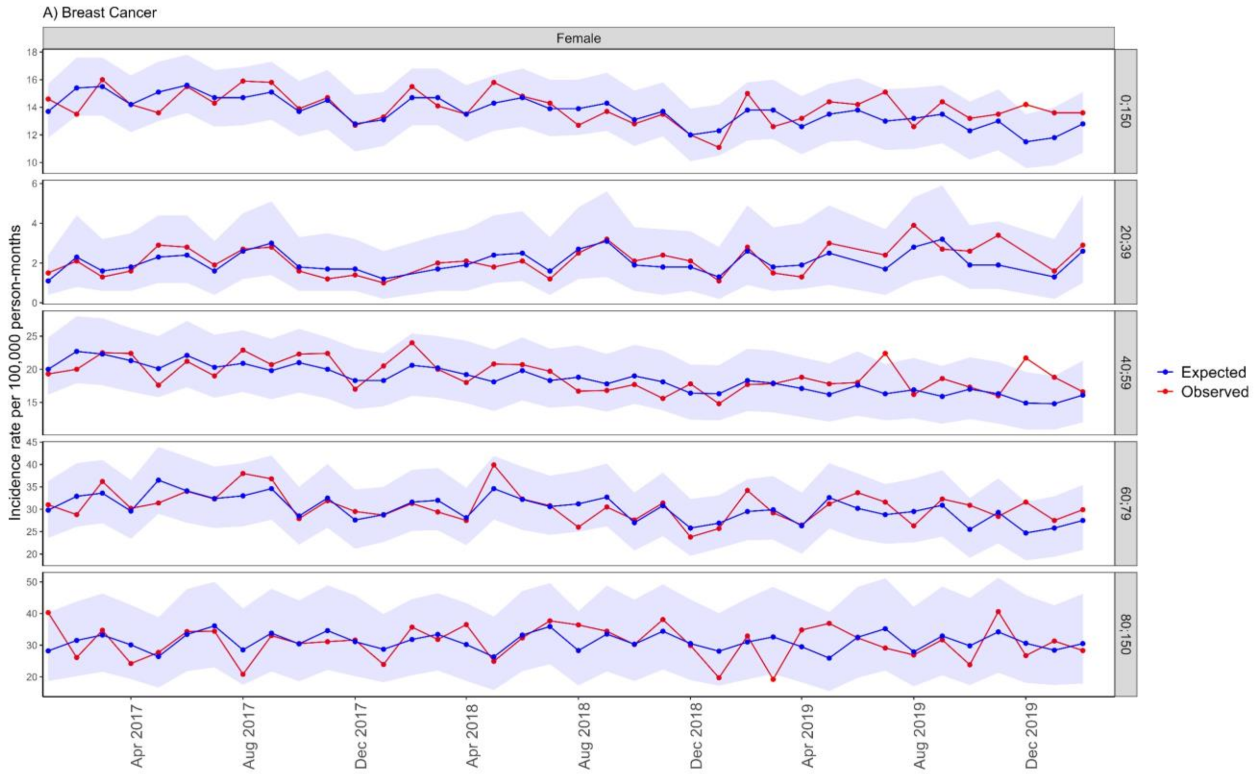

A) Breast Cancer
Female
Incidence rate per 100,000 person-months
0;150
20;39
40;59
60;79
80;150
Expected
Observed
Apr 2017
Aug 2017
Dec 2017
Apr 2018
Aug 2018
Dec 2018
Apr 2019
Aug 2019
Dec 2019


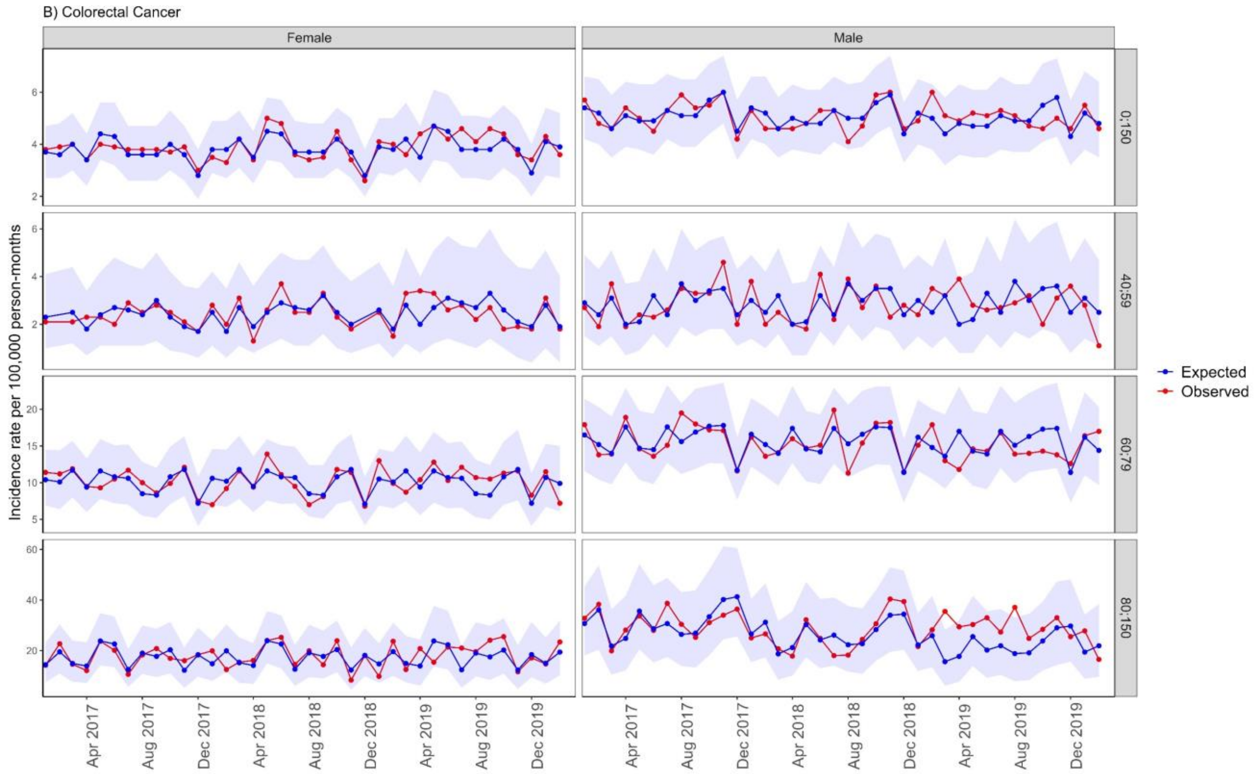

B) Colorectal Cancer
Female
Male
Incidence rate per 100,000 person-months
0;150
40;59
60;79
80;150
Expected
Observed
Apr 2017
Aug 2017
Dec 2017
Apr 2018
Aug 2018
Dec 2018
Apr 2019
Aug 2019
Dec 2019

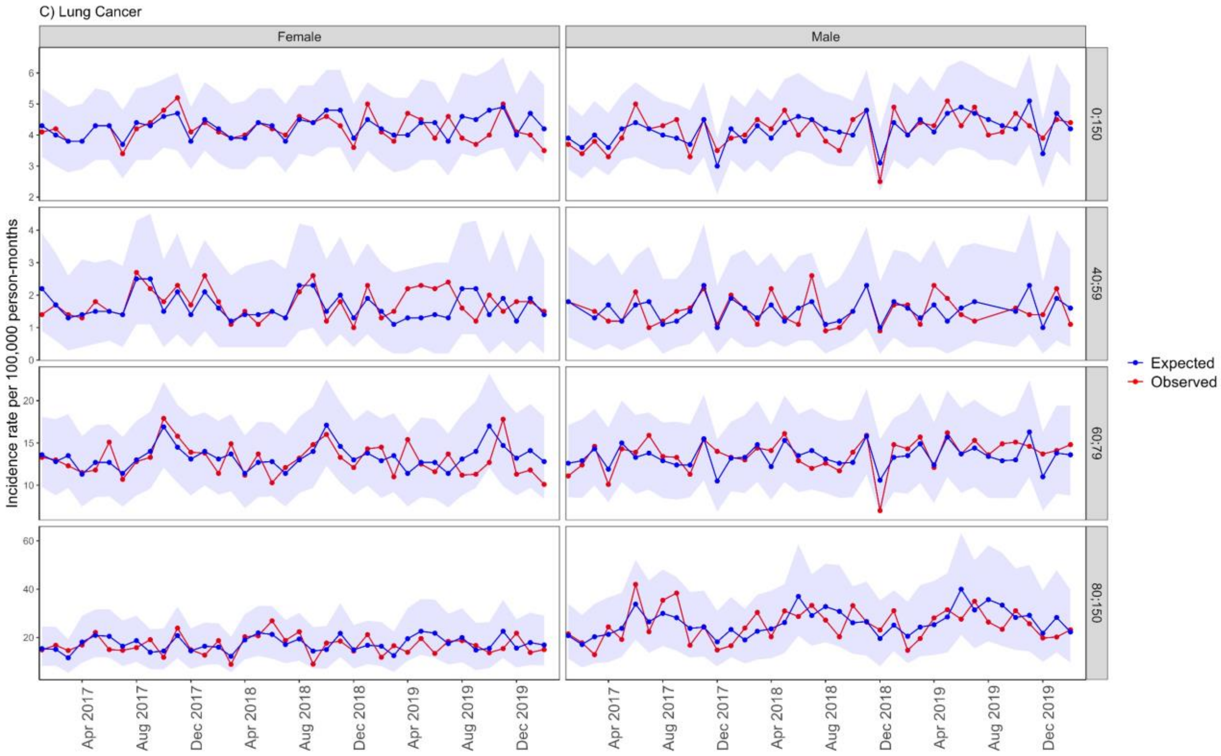


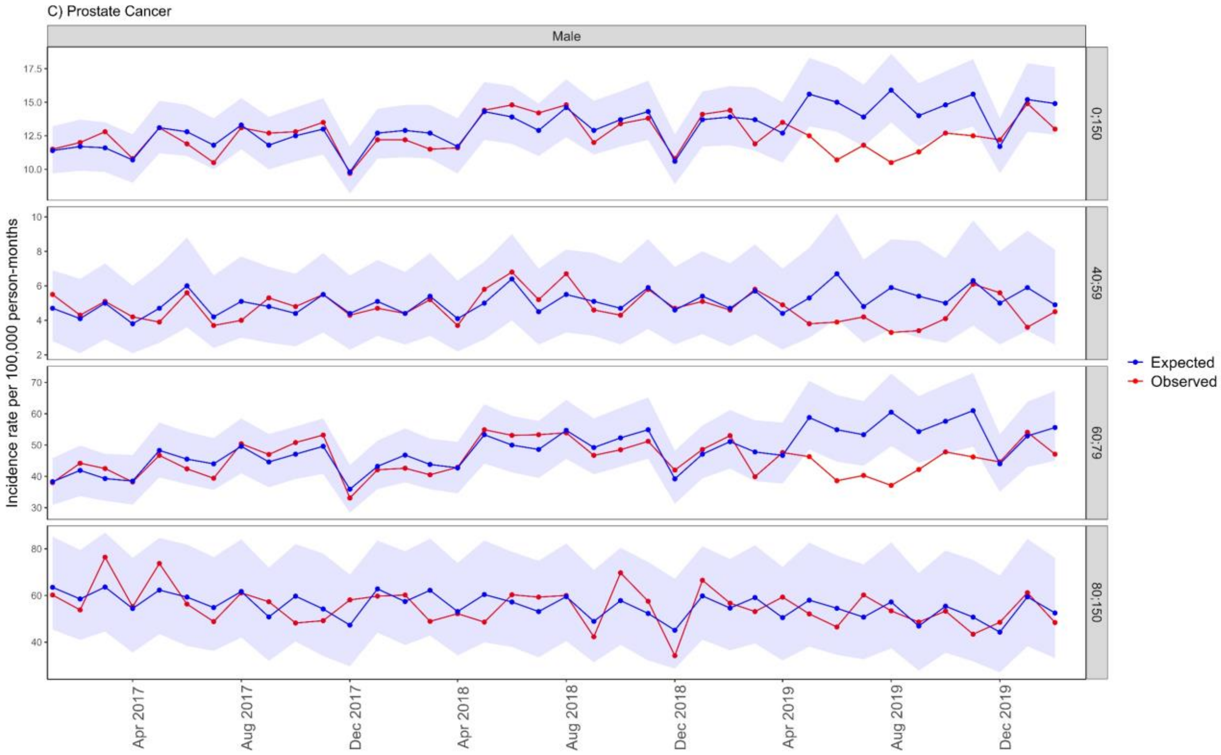


**Figure S2.** Expected and observed incidence rates per 100,000 person months of A) breast cancer, B) colorectal cancer, C) lung cancer, and D) prostate cancer in primary care records from CPRD UK, after lockdown, stratified by age and sex, from the validation model. Note. Points represent monthly incidence rates. Expected rates (with 95% prediction intervals) were calculated using negative binomial regression using observed data from January 2017 to February 2020 to estimate expected counts during the same period to validate the methodology. The shaded areas represent the 95% prediction intervals.

**Table S2.** Source population starting counts, final counts after applying excision criteria, and reasons for exclusion

| Denominator count (n) | Reason for exclusion | Excluded (n) |
|---|---|---|
| 16,958,587 | Starting population | NA |
| 16,958,587 | Missing year of birth | 0 |
| 16,958,587 | Missing sex | 0 |
| 16,958,587 | Cannot satisfy age criteria during the study period based on year of birth | 0 |
| 5,694,486 | No observation time available during study period (January 2017 to December 2021) | 11,264,101 |
| 5,694,486 | Doesn't satisfy age criteria during the study period | 0 |
| 5,191,650 | Prior history requirement not fulfilled during study period | 502,836 |
| 5,191,268 | Excluded due to prior breast cancer diagnosis (do not pass outcome washout during study period) | 382 |
| 5,191,291 | Excluded due to prior colorectal cancer diagnosis (do not pass outcome washout during study period) | 359 |
| 5,191,131 | Excluded due to prior lung cancer diagnosis (do not pass outcome washout during study period) | 519 |
| 5,191,367 | Excluded due to prior prostate cancer diagnosis (do not pass outcome washout during study period) | 283 |

**Table S3.** Proportion of source population in different age groups for those enrolled in CPRD GOLD before, during and after lockdown

| Age Group | Proportion Pre-pandemic (Jan 2017-Feb 2020) | Proportion During Lockdown (March 2020-June 2020) | Proportion After lockdown (July 2020-Dec 2021) |
|---|---|---|---|
| 0 to 9 years | 24.45% | 24.91% | 24.46% |
| 10 to 19 years | 9.38% | 9.09% | 8.99% |
| 20 to 29 years | 15.09% | 14.49% | 15.71% |
| 30 to 39 years | 17.24% | 17.46% | 17.57% |
| 40 to 49 years | 13.16% | 13.79% | 13.38% |
| 50 to 59 years | 10.04% | 10.54% | 10.25% |
| 60 to 69 years | 6.41% | 6.38% | 6.2% |
| 70 to 79 years | 2.85% | 2.44% | 2.43% |
| 80 to 89 years | 1.07% | 0.73% | 0.79% |
| 90 to 99 years | 0.3% | 0.16% | 0.2% |
| 100+ years | 0.01% | 0.01% | 0.01% |

Note. Counts before applying exclusion criteria

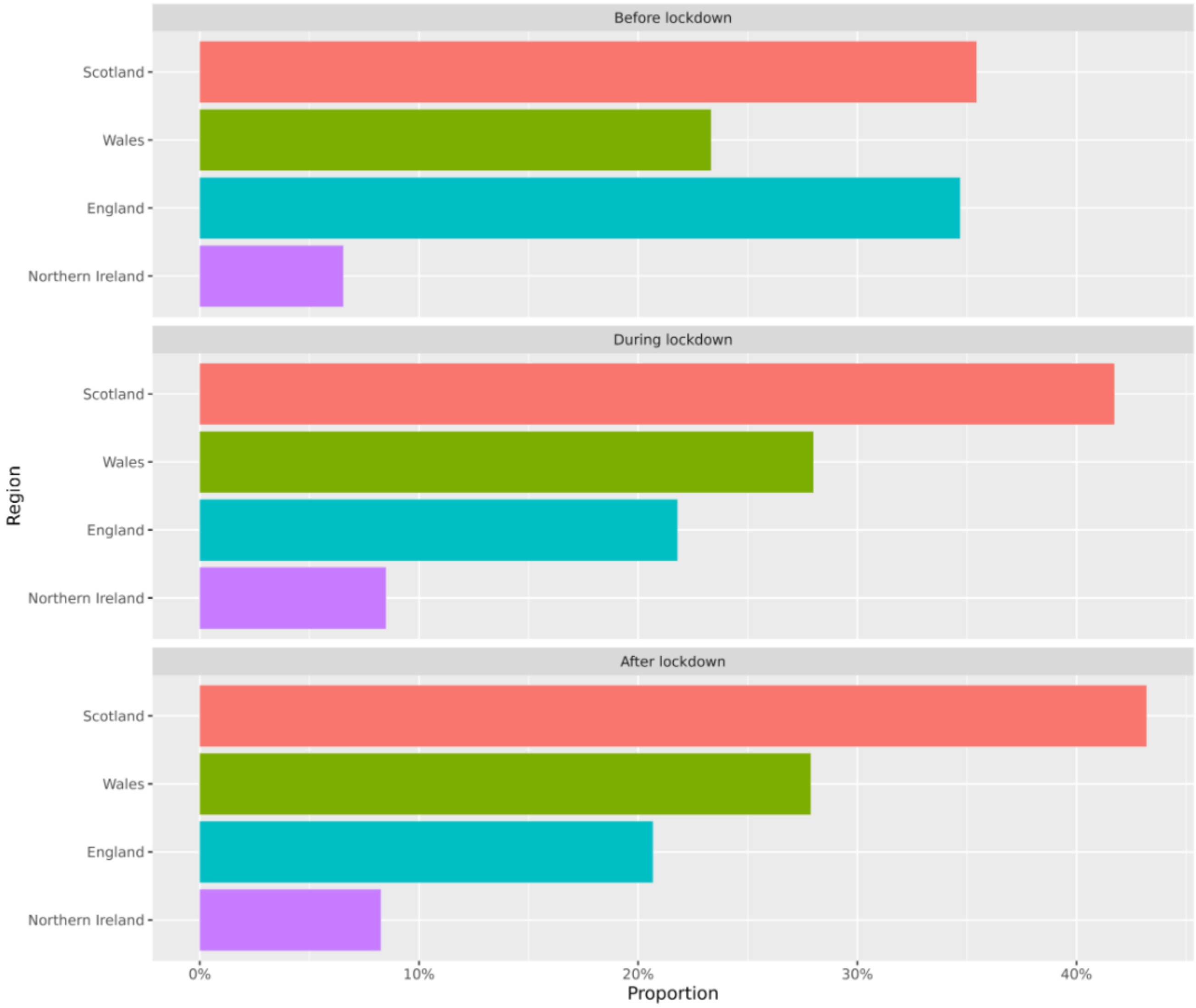


**Figure S3**. Proportion of source population observed in each region of the UK in the CPRD GOLD, for those enrolled before, during and after lockdown

**Table S4.** Counts of healthcare visits, screening and diagnostic procedures, before, during and after lockdown (including descendant concepts) in the denominator population

| Screening/Diagnostic Test | N Pre-pandemic (Jan 2017-Feb 2020) | N During Lockdown (March 2020-June 2020) | N After lockdown (July 2020-Dec 2021) |
|---|---|---|---|
| Visits in the healthcare system | 184,471,700 | 12,108,287 | 74,106,599 |
| *Breast* | | | |
| Referral to Breast clinic | 31,079 | 1571 | 10,535 |
| Referral to mammography clinic | 1634 | 64 | 515 |
| Fasttrack referral for suspected breast cancer | 13,203 | 707 | 4277 |
| Referral to breast surgeon | 21,443 | 1014 | 7489 |
| Seen in breast clinic | 106,697 | 4114 | 28,463 |
| Seen by breast surgeon | 4427 | 116 | 332 |
| Diagnostic mammograms | 1139 | 14 | 124 |
| Screening mammography | 2295 | 78 | 160 |
| Diagnostic mammogram and ultrasound | 264,009 | 2561 | 46,480 |
| Biopsy of breast | 4906 | 293 | 1987 |
| Stereotactically guided core needle biopsy of breast | 1483 | 70 | 539 |
| Percutaneous needle biopsy of breast | 1555 | 71 | 556 |
| Fine needle aspiration of breast | 586 | 43 | 201 |
| Wire guided local excision of breast lump | 218 | 7 | 52 |
| Excision of mammary duct | 223 | 6 | 45 |
| Wide local excision of breast lesion | 3247 | 215 | 1038 |
| Excision of lesion of breast | 4888 | 275 | 1484 |
| Excision of breast tissue | 9652 | 511 | 2810 |
| *Colorectal* | | | |
| Fast track referral for suspected colorectal cancer | 7977 | 472 | 2954 |
| Bowel cancer screening programme | 10,172 | 275 | 3554 |
| Quantitative faecal immunochemical tests | 12 | 0 | 23 |
| Screening colonoscopy | 0 | 0 | 0 |
| Colonoscopies | 340 | <5 | 81 |

**Table S4.** Counts of healthcare visits, screening and diagnostic procedures, before, during and after lockdown (including descendant concepts) in the denominator population

| Screening/Diagnostic Test | N Pre-pandemic (Jan 2017-Feb 2020) | N During Lockdown (March 2020-June 2020) | N After lockdown (July 2020-Dec 2021) |
|---|---|---|---|
| Ultrasonography of intestine | 0 | 0 | 0 |
| Endoscopic ultrasound of upper gastrointestinal tract | 124 | <5 | 15 |
| Ultrasound of gastrointestinal tract | 124 | <5 | 15 |
| Ultrasonography of abdomen | 18,303 | 532 | 4989 |
| Ultrasonography of rectum | 0 | 0 | 0 |
| Sigmoidoscopy | 234 | <5 | 57 |
| *Lung* | | | |
| Fast track referral for suspected lung cancer | 2105 | 79 | 547 |
| Bronchoscopy | 133 | 5 | 22 |
| Endobronchial ultrasonography guided transbronchial needle aspiration | 0 | 0 | 0 |
| Mediastinoscopy | 106 | 5 | 23 |
| Mediastinoscopy - inspection only | 33 | <5 | 12 |
| CT and biopsy of chest | 0 | 0 | 0 |
| US scan and biopsy of chest | 0 | 0 | 0 |
| Diagnostic Radiology (Diagnostic Imaging) Procedures of the Chest | 0 | 0 | 0 |
| MRI of chest | 394 | 19 | 110 |
| *Prostate* | | | |
| PSA monitoring | 2920 | 139 | 825 |
| Prostate specific antigen measurement | 521,564 | 21,683 | 177,358 |
| PSA (prostate-specific antigen) level measurement | 1055 | 24 | 219 |
| Biopsy of prostate | 3269 | 155 | 1052 |
| Open biopsy of prostate | 114 | 9 | 46 |

Note. Counts <5 masked to maintain patient confidentiality.

**Table S5.** Counts of healthcare visits, screening/ diagnostic tests and referrals, occurring in the month, ≤6 months, and >6 months prior to diagnosis date, for breast cancer patients diagnosed before, during and after lockdown

| Screening/diagnostic test | Time window prior to diagnosis date | N Pre-pandemic (8,815) | N During Lockdown (459) | N After Lockdown (2,900) |
|---|---|---|---|---|
| Biopsy of breast | 1 to 30 days | 550 | 45 | 268 |
| | 31 to 180 days | 87 | 5 | 37 |
| | 180+ days | 335 | 14 | 113 |
| Diagnostic mammogram & ultrasound | 1 to 30 days | 921 | 35 | 288 |
| | 31 to 180 days | 397 | 14 | 79 |
| | 180+ days | 12,161 | 663 | 3,876 |
| Diagnostic mammograms | 1 to 30 days | 13 | <5 | <5 |
| | 31 to 180 days | 5 | 0 | <5 |
| | 180+ days | 54 | <5 | 13 |
| Excision of breast tissue | 1 to 30 days | 206 | 10 | 62 |
| | 31 to 180 days | 74 | 5 | 25 |
| | 180+ days | 333 | 18 | 101 |
| Excision of lesion of breast | 1 to 30 days | 153 | 6 | 41 |
| | 31 to 180 days | 48 | <5 | 16 |
| | 180+ days | 227 | 11 | 57 |
| Excision of mammary duct | 1 to 30 days | <5 | 0 | <5 |
| | 31 to 180 days | <5 | 0 | 0 |
| | 180+ days | 23 | <5 | 13 |
| Fasttrack referral for suspected breast cancer | 1 to 30 days | 349 | 30 | 94 |
| | 31 to 180 days | 182 | 10 | 74 |
| | 180+ days | 87 | 8 | 22 |
| Fine needle aspiration of breast | 1 to 30 days | 25 | <5 | 13 |
| | 31 to 180 days | 5 | 0 | <5 |
| | 180+ days | 100 | <5 | 20 |
| Percutaneous needle biopsy of breast | 1 to 30 days | 255 | 18 | 108 |
| | 31 to 180 days | 36 | <5 | 10 |

**Table S5.** Counts of healthcare visits, screening/ diagnostic tests and referrals, occurring in the month, ≤6 months, and >6 months prior to diagnosis date, for breast cancer patients diagnosed before, during and after lockdown

| Screening/diagnostic test | Time window prior to diagnosis date | N Pre-pandemic (8,815) | N During Lockdown (459) | N After Lockdown (2,900) |
|---|---|---|---|---|
| | 180+ days | 58 | <5 | 21 |
| Referral to breast clinic | 1 to 30 days | 490 | 36 | 161 |
| | 31 to 180 days | 419 | 25 | 185 |
| | 180+ days | 559 | 28 | 228 |
| Referral to breast surgeon | 1 to 30 days | 360 | 33 | 103 |
| | 31 to 180 days | 295 | 13 | 120 |
| | 180+ days | 290 | 26 | 110 |
| Referral to mammography clinic | 1 to 30 days | 27 | <5 | 11 |
| | 31 to 180 days | 29 | <5 | 11 |
| | 180+ days | 56 | <5 | 8 |
| Screening mammography | 1 to 30 days | 12 | 0 | 0 |
| | 31 to 180 days | 9 | 0 | 0 |
| | 180+ days | 118 | <5 | 9 |
| Seen by breast surgeon | 1 to 30 days | 125 | 9 | 18 |
| | 31 to 180 days | 37 | 5 | <5 |
| | 180+ days | 59 | <5 | 21 |
| Seen in breast clinic | 1 to 30 days | 2,437 | 129 | 663 |
| | 31 to 180 days | 596 | 36 | 196 |
| | 180+ days | 2,056 | 115 | 705 |
| Stereotactically guided core needle biopsy | 1 to 30 days | 252 | 17 | 108 |
| | 31 to 180 days | 36 | <5 | 10 |
| | 180+ days | 56 | 0 | 19 |
| Visits within healthcare system | 1 to 30 days | 32,146 | 1,798 | 11,123 |
| | 31 to 180 days | 87,799 | 4,389 | 30,649 |
| | 180+ days | 2,340,878 | 126,691 | 827,739 |
| Wide local excision of breast lesion | 1 to 30 days | 111 | <5 | 31 |

**Table S5.** Counts of healthcare visits, screening/ diagnostic tests and referrals, occurring in the month, ≤6 months, and >6 months prior to diagnosis date, for breast cancer patients diagnosed before, during and after lockdown

| **Screening/diagnostic test** | **Time window prior to diagnosis date** | **N Pre-pandemic (8,815)** | **N During Lockdown (459)** | **N After Lockdown (2,900)** |
|---|---|---|---|---|
| | 31 to 180 days | 34 | <5 | 8 |
| | 180+ days | 31 | 0 | <5 |
| Wire guided local excision of breast lump | 1 to 30 days | 10 | <5 | <5 |
| | 31 to 180 days | <5 | 0 | <5 |
| | 180+ days | <5 | 0 | <5 |

Note. Pre-pandemic = Jan 2017-Feb 2020; During Lockdown = March 2020-June 2020; After lockdown = July 2020-Dec 2021. Counts <5 masked to maintain patient confidentiality.

**Table S6.** Counts of healthcare visits, screening and diagnostic procedures, occurring in the month, 6 months, and 6 months+, for colorectal cancer patients diagnosed before, during and after lockdown (including descendant concepts)

| **Screening/diagnostic test** | **Time window prior to index date** | **N Pre-pandemic (6,025)** | **N During Lockdown (349)** | **N After Lockdown (2,234)** |
|---|---|---|---|---|
| Bowel cancer screening programme | 1 to 30 days | 14 | 0 | <5 |
| | 31 to 180 days | 38 | <5 | 7 |
| | 180+ days | 360 | 19 | 104 |
| Colonoscopies | 1 to 30 days | <5 | 0 | 0 |
| | 31 to 180 days | <5 | 0 | <5 |
| | 180+ days | 8 | 0 | <5 |
| Visits within healthcare system | 1 to 30 days | 28,556 | 1,722 | 11,472 |
| | 31 to 180 days | 85,288 | 4,532 | 32,678 |
| | 180+ days | 1,751,840 | 105,839 | 705,013 |

Note. Pre-pandemic = Jan 2017-Feb 2020; During Lockdown = March 2020-June 2020; After lockdown = July 2020-Dec 2021. There were no results for the following screening/diagnostic tests across these time-periods: Endoscopic ultrasound of upper gastrointestinal tract; Fast track referral for suspected colorectal cancer; Quantitative faecal immunochemical tests; Screening colonoscopies; Sigmoidoscopy; Ultrasonography of abdomen; Ultrasonography of intestine; Ultrasonography of rectum; Ultrasound of gastrointestinal tract. Counts <5 masked to maintain patient confidentiality.

**Table S7.** Counts of healthcare visits, screening and diagnostic procedures, occurring in the month, 6 months, and 6 months+, for lung cancer patients diagnosed before, during and after lockdown (including descendant concepts)

| **Screening/diagnostic test** | **Time window prior to index date** | **N Pre-pandemic (5,766)** | **N During Lockdown (435)** | **N After Lockdown (2,102)** |
|---|---|---|---|---|
| Bronchoscopy | 1 to 30 days | 0 | <5 | <5 |
| | 31 to 180 days | <5 | 0 | 0 |
| | 180+ days | <5 | 0 | <5 |
| Mediastinoscopy | 1 to 30 days | <5 | 0 | 0 |
| | 31 to 180 days | <5 | 0 | 0 |
| | 180+ days | <5 | 0 | <5 |
| Visits within healthcare system | 1 to 30 days | 35,816 | 2,811 | 13,483 |
| | 31 to 180 days | 98,896 | 7,452 | 35,804 |
| | 180+ days | 2,056,404 | 164,494 | 825,358 |

Note. Pre-pandemic = Jan 2017-Feb 2020; During Lockdown = March 2020-June 2020; After lockdown = July 2020-Dec 2021. There were no results for the following screening/diagnostic tests across these time-periods: CT and biopsy of chest; Diagnostic Radiology (Diagnostic Imaging) Procedures of the Chest; Endobronchial ultrasonography guided transbronchial needle aspiration; Fast track referral for lung cancer; MRI of chest; US scan and biopsy of chest. Counts < masked to maintain patient confidentiality.

**Table S8.** Counts of healthcare visits, screening and diagnostic procedures, occurring in the month, 6 months, and 6 months+, for prostate cancer patients diagnosed before, during and after lockdown (including descendant concepts)

| Screening/diagnostic test | Time window prior to index date | N Pre-pandemic (8,103) | N during lockdown (427) | N after lockdown (4,277) |
|---|---|---|---|---|
| Visits within healthcare system | 1 to 30 days | 32,686 | 1,734 | 10,956 |
| | 31 to 180 days | 118,353 | 6,358 | 38,489 |
| | 180+ days | 2,125,748 | 120,351 | 716,009 |

Note. Pre-pandemic = Jan 2017-Feb 2020; During Lockdown = March 2020-June 2020; After lockdown = July 2020-Dec 2021. There were no results for the following screening/diagnostic tests across these time-periods: Biopsy of prostate; Open biopsy of prostate; PSA monitoring; Prostate specific antigen measurement – level; Prostate specific antigen measurement.

**Table S9.** Characterisation of breast cancer patients diagnosed before, during and after lockdown

| | Pre-pandemic | During Lockdown | After Lockdown | Standardised Mean Differences | |
|---|---|---|---|---|---|
| **Characteristic** | **Count (% of n=8,815)** | **Count (% of n=459)** | **Count (% of n=2,900)** | **Before vs. during lockdown** | **Before vs. after lockdown** |
| **Medical history: General** | | | | | |
| Acute respiratory disease | 374 (4.2) | 15 (3.3) | 46 (1.6) | -0.05 | **-0.16** |
| Attention deficit hyperactivity disorder | <5 (~0) | 0 (0) | 0 (0) | -0.02 | -0.02 |
| Chronic liver disease | 12 (0.1) | 0 (0) | 5 (0.2) | -0.05 | 0.01 |
| Chronic obstructive lung disease | 112 (1.3) | <5 (~0) | 26 (0.9) | **-0.12** | -0.04 |
| Crohn's disease | <5 (~0) | 0 (0) | <5 (~0) | -0.03 | 0.00 |
| Dementia | 29 (0.3) | <5 (~0) | 13 (0.4) | 0.02 | 0.02 |
| Depressive disorder | 102 (1.2) | 5 (1.1) | 19 (0.7) | -0.01 | -0.05 |
| Diabetes mellitus | 89 (1) | <5 (~0) | 33 (1.1) | -0.10 | 0.01 |
| Gastroesophageal reflux disease | 27 (0.3) | <5 (~0) | 11 (0.4) | 0.02 | 0.01 |
| Gastrointestinal hemorrhage | 60 (0.7) | <5 (~1) | 11 (0.4) | 0.02 | -0.04 |
| Hyperlipidemia | 43 (0.5) | <5 (~0) | <5 (0.1) | -0.05 | -0.06 |
| Hypertensive disorder | 141 (1.6) | 5 (1.1) | 44 (1.5) | -0.04 | -0.01 |
| Lesion of liver | 29 (0.3) | 0 (0) | 12 (0.4) | -0.08 | 0.01 |
| Obesity | 18 (0.2) | <5 (~0) | <5 (~0) | 0.00 | -0.02 |
| Osteoarthritis | 195 (2.2) | 6 (1.3) | 39 (1.3) | -0.07 | -0.07 |
| Pneumonia | 39 (0.4) | <5 (~0) | 13 (0.4) | -0.04 | 0.00 |
| Psoriasis | 47 (0.5) | <5 (~1) | 9 (0.3) | 0.02 | -0.03 |
| Renal impairment | 100 (1.1) | 5 (1.1) | 31 (1.1) | 0.00 | -0.01 |

**Table S9.** Characterisation of breast cancer patients diagnosed before, during and after lockdown

| | Pre-pandemic | During Lockdown | After Lockdown | Standardised Mean Differences | |
|---|---|---|---|---|---|
| **Characteristic** | **Count (% of n=8,815)** | **Count (% of n=459)** | **Count (% of n=2,900)** | **Before vs. during lockdown** | **Before vs. after lockdown** |
| Rheumatoid arthritis | 18 (0.2) | <5 (~0) | 7 (0.2) | 0.00 | 0.01 |
| Schizophrenia | <5 (~0) | 0 (0) | <5 (~0) | -0.02 | 0.02 |
| Ulcerative colitis | 7 (0.1) | 0 (0) | <5 (~0) | -0.04 | -0.02 |
| Urinary tract infectious disease | 212 (2.4) | 7 (1.5) | 53 (1.8) | -0.06 | -0.04 |
| Visual system disorder | 415 (4.7) | 24 (5.2) | 88 (3) | 0.02 | -0.09 |
| **Medical history: Cardiovascular disease** | | | | | |
| Atrial fibrillation | 53 (0.6) | <5 (~1) | 14 (0.5) | 0.03 | -0.02 |
| Cerebrovascular disease | 33 (0.4) | 0 (0) | 8 (0.3) | -0.09 | -0.02 |
| Coronary arteriosclerosis | <5 (~0) | 0 (0) | <5 (~0) | -0.03 | -0.01 |
| Heart disease | 153 (1.7) | 10 (2.2) | 45 (1.6) | 0.03 | -0.01 |
| Heart failure | 21 (0.2) | <5 (~1) | 7 (0.2) | 0.06 | 0.00 |
| Ischemic heart disease | 31 (0.4) | <5 (~1) | 14 (0.5) | 0.01 | 0.02 |
| Peripheral vascular disease | 12 (0.1) | 0 (0) | <5 (~0) | -0.05 | 0.00 |
| Pulmonary embolism | 18 (0.2) | 0 (0) | 11 (0.4) | -0.06 | 0.03 |
| Venous thrombosis | 57 (0.6) | 0 (0) | 14 (0.5) | **-0.11** | -0.02 |
| **Medical history: Neoplasms** | | | | | |
| Hematologic neoplasm | 40 (0.5) | 0 (0) | 21 (0.7) | **-0.10** | 0.04 |
| Malignant lymphoma | 8 (0.1) | <5 (~0) | <5 (~0) | 0.03 | 0.01 |
| Malignant neoplasm of anorectum | 11 (0.1) | 0 (0) | <5 (~0) | -0.05 | -0.02 |

**Table S9.** Characterisation of breast cancer patients diagnosed before, during and after lockdown

| | **Pre-pandemic** | **During Lockdown** | **After Lockdown** | **Standardised Mean Differences** | |
|---|---|---|---|---|---|
| **Characteristic** | **Count (% of n=8,815)** | **Count (% of n=459)** | **Count (% of n=2,900)** | **Before vs. during lockdown** | **Before vs. after lockdown** |
| Malignant neoplastic disease | 8815 (100) | 459 (100) | 2900 (100) | / | / |
| Malignant tumor of breast | 8815 (100) | 459 (100) | 2900 (100) | / | / |
| Malignant tumor of colon | 12 (0.1) | 0 (0) | <5 (~0) | -0.05 | -0.02 |
| Malignant tumor of lung | 29 (0.3) | <5 (~1) | 9 (0.3) | 0.05 | 0.00 |
| Malignant tumor of urinary bladder | <5 (~0) | 0 (0) | <5 (~0) | -0.03 | 0.00 |
| **Medication use** | | | | | |
| Agents acting on the renin-angiotensin system | 1967 (22.3) | 94 (20.5) | 650 (22.4) | -0.04 | 0.00 |
| Antibacterials for systemic use | 3783 (42.9) | 212 (46.2) | 1129 (38.9) | 0.07 | -0.08 |
| Antidepressants | 2388 (27.1) | 119 (25.9) | 807 (27.8) | -0.03 | 0.02 |
| Antiepileptics | 659 (7.5) | 42 (9.2) | 207 (7.1) | 0.06 | -0.01 |
| Antiinflammatory and antirheumatic products | 2662 (30.2) | 128 (27.9) | 774 (26.7) | -0.05 | -0.08 |
| Antineoplastic agents | 139 (1.6) | 5 (1.1) | 39 (1.3) | -0.04 | -0.02 |
| Antipsoriatics | 427 (4.8) | 26 (5.7) | 137 (4.7) | 0.04 | -0.01 |
| Antithrombotic agents | 1329 (15.1) | 63 (13.7) | 446 (15.4) | -0.04 | 0.01 |
| Beta blocking agents | 1330 (15.1) | 54 (11.8) | 427 (14.7) | **-0.10** | -0.01 |
| Calcium channel blockers | 1311 (14.9) | 61 (13.3) | 476 (16.4) | -0.05 | 0.04 |

**Table S9.** Characterisation of breast cancer patients diagnosed before, during and after lockdown

| | Pre-pandemic | During Lockdown | After Lockdown | Standardised Mean Differences | |
|---|---|---|---|---|---|
| **Characteristic** | **Count (% of n=8,815)** | **Count (% of n=459)** | **Count (% of n=2,900)** | **Before vs. during lockdown** | **Before vs. after lockdown** |
| Diuretics | 1417 (16.1) | 66 (14.4) | 423 (14.6) | -0.05 | -0.04 |
| Drugs for acid related disorders | 3433 (38.9) | 162 (35.3) | 1138 (39.2) | -0.08 | 0.01 |
| Drugs for obstructive airway diseases | 2498 (28.3) | 128 (27.9) | 766 (26.4) | -0.01 | -0.04 |
| Drugs used in diabetes | 643 (7.3) | 31 (6.8) | 210 (7.2) | -0.02 | 0.00 |
| Immunosuppressants | 126 (1.4) | <5 (~1) | 28 (1) | -0.08 | -0.04 |
| Lipid modifying agents | 2191 (24.9) | 95 (20.7) | 676 (23.3) | **-0.10** | -0.04 |
| Opioids | 2443 (27.7) | 120 (26.1) | 758 (26.1) | -0.04 | -0.04 |
| Psycholeptics | 1524 (17.3) | 88 (19.2) | 451 (15.6) | 0.05 | -0.05 |
| Psychostimulants | <5 (~0) | 0 (0) | <5 (~0) | -0.02 | 0.04 |
| **Comorbidity Scales** | **Mean (SD)** | **Mean (SD)** | **Mean (SD)** | **Std.Diff** | **Std.Diff** |
| Charlson comorbidity index | 2.8 (1.4) | 2.7 (1.3) | 2.8 (1.4) | -0.05 | -0.02 |
| CHADS2Vasc | 2.1 (1.3) | 2.1 (1.3) | 2.1 (1.3) | -0.02 | 0.01 |
| DCSI | 0.7 (1.5) | 0.6 (1.5) | 0.7 (1.5) | -0.03 | -0.01 |

Note. CHADS2Vasc: Comorbidity scale measuring congestive heart failure, hypertension, age, diabetes, stroke, vascular disease, age and sex. DCSI: Diabetes Complications and Severity Index. SD = Standard Deviation. Counts <5 masked and proportions rounded to nearest 1% in order for frequencies <5 to remain masked. Bold indicates significant standardised mean differences between lockdown periods (>.10).

**Table S10.** Characterisaton of colorectal cancer patients diagnosed before, during and after lockdown

| | Pre-pandemic | During Lockdown | After Lockdown | Standardised Mean Differences | |
|---|---|---|---|---|---|
| **Characteristic** | **Count (% of n=6,025)** | **Count (% of n=349)** | **Count (% of n=2,234)** | **Before vs. during lockdown** | **Before vs. after lockdown** |
| **Medical history: General** | | | | | |
| Acute respiratory disease | 232 (3.9) | 10 (2.9) | 46 (2.1) | -0.05 | **-0.11** |
| Chronic liver disease | 16 (0.3) | <5 (~0) | 7 (0.3) | 0.00 | 0.01 |
| Chronic obstructive lung disease | 128 (2.1) | 10 (2.9) | 33 (1.5) | 0.05 | -0.05 |
| Crohn's disease | <5 (~0) | <5 (~0) | 5 (~0) | 0.05 | 0.04 |
| Dementia | 44 (0.7) | <5 (~1) | 15 (0.7) | 0.01 | -0.01 |
| Depressive disorder | 60 (1) | 6 (1.7) | 10 (0.4) | 0.06 | -0.06 |
| Diabetes mellitus | 110 (1.8) | 8 (2.3) | 36 (1.6) | 0.03 | -0.02 |
| Gastroesophageal reflux disease | 35 (0.6) | <5 (~0) | 5 (0.2) | -0.04 | -0.06 |
| Gastrointestinal hemorrhage | 610 (10.1) | 23 (6.6) | 163 (7.3) | **-0.13** | **-0.10** |
| Hyperlipidemia | 17 (0.3) | <5 (~0) | 9 (0.4) | 0.00 | 0.02 |
| Hypertensive disorder | 83 (1.4) | 5 (1.4) | 27 (1.2) | 0.00 | -0.01 |
| Lesion of liver | 193 (3.2) | 19 (5.4) | 102 (4.6) | **0.11** | 0.07 |
| Obesity | 17 (0.3) | 0 (0) | 8 (0.4) | -0.08 | 0.01 |
| Osteoarthritis | 141 (2.3) | 12 (3.4) | 26 (1.2) | 0.07 | -0.09 |
| Pneumonia | 51 (0.8) | <5 (~1) | 21 (0.9) | 0.00 | 0.01 |
| Psoriasis | 20 (0.3) | <5 (~0) | 7 (0.3) | -0.01 | 0.00 |
| Renal impairment | 167 (2.8) | 9 (2.6) | 36 (1.6) | -0.01 | -0.08 |
| Rheumatoid arthritis | 10 (0.2) | <5 (0.3) | <5 (0.2) | 0.03 | 0.00 |
| Schizophrenia | <5 (~0) | 0 (0) | 0 (0) | -0.03 | -0.03 |
| Ulcerative colitis | 9 (0.1) | <5 (~1) | <5 (~0) | 0.10 | 0.01 |
| Urinary tract infectious disease | 136 (2.3) | 9 (2.6) | 37 (1.7) | 0.02 | -0.04 |
| Visual system disorder | 375 (6.2) | 16 (4.6) | 76 (3.4) | -0.07 | **-0.13** |

**Table S10.** Characterisaton of colorectal cancer patients diagnosed before, during and after lockdown

| | Pre-pandemic | During Lockdown | After Lockdown | Standardised Mean Differences | |
|---|---|---|---|---|---|
| **Characteristic** | **Count (% of n=6,025)** | **Count (% of n=349)** | **Count (% of n=2,234)** | **Before vs. during lockdown** | **Before vs. after lockdown** |
| **Medical history: Cardiovascular disease** | | | | | |
| Atrial fibrillation | 142 (2.4) | 10 (2.9) | 60 (2.7) | 0.03 | 0.02 |
| Cerebrovascular disease | 61 (1) | 8 (2.3) | 23 (1) | **0.10** | 0.00 |
| Coronary arteriosclerosis | 15 (0.2) | 0 (0) | <5 (~0) | -0.07 | -0.04 |
| Heart disease | 333 (5.5) | 25 (7.2) | 133 (6) | 0.07 | 0.02 |
| Heart failure | 43 (0.7) | <5 (~1) | 18 (0.8) | 0.05 | 0.01 |
| Ischemic heart disease | 92 (1.5) | <5 (~1) | 33 (1.5) | -0.03 | 0.00 |
| Peripheral vascular disease | 19 (0.3) | <5 (~1) | 5 (0.2) | 0.04 | -0.02 |
| Pulmonary embolism | 62 (1) | <5 (~1) | 25 (1.1) | 0.01 | 0.01 |
| Venous thrombosis | 66 (1.1) | <5 (~1) | 26 (1.2) | 0.00 | 0.01 |
| **Medical history: Neoplasms** | | | | | |
| Hematologic neoplasm | 13 (0.2) | <5 (~1) | 5 (0.2) | 0.09 | 0.00 |
| Malignant lymphoma | 7 (0.1) | <5 (~0) | <5 (~0) | 0.04 | -0.01 |
| Malignant neoplasm of anorectum | 1767 (29.3) | 94 (26.9) | 642 (28.7) | -0.05 | -0.01 |
| Malignant neoplastic disease | 6025 (100) | 349 (100) | 2234 (100) | / | / |
| Malignant tumor of breast | 17 (0.3) | 0 (0) | 6 (0.3) | -0.08 | 0.00 |
| Malignant tumor of colon | 3866 (64.2) | 223 (63.9) | 1439 (64.4) | -0.01 | 0.01 |
| Malignant tumor of lung | 49 (0.8) | 5 (1.4) | 17 (0.8) | 0.06 | -0.01 |
| Malignant tumor of urinary bladder | 8 (0.1) | <5 (~1) | <5 (~0) | 0.07 | -0.01 |
| **Medication use** | | | | | |
| Agents acting on the renin-angiotensin system | 2134 (35.4) | 118 (33.8) | 707 (31.6) | -0.03 | -0.08 |
| Antibacterials for systemic use | 2686 (44.6) | 146 (41.8) | 876 (39.2) | -0.06 | -0.11 |

**Table S10.** Characterisaton of colorectal cancer patients diagnosed before, during and after lockdown

| | Pre-pandemic | During Lockdown | After Lockdown | Standardised Mean Differences | |
|---|---|---|---|---|---|
| **Characteristic** | **Count (% of n=6,025)** | **Count (% of n=349)** | **Count (% of n=2,234)** | **Before vs. during lockdown** | **Before vs. after lockdown** |
| Antidepressants | 1219 (20.2) | 88 (25.2) | 490 (21.9) | **0.12** | 0.04 |
| Antiepileptics | 438 (7.3) | 32 (9.2) | 166 (7.4) | 0.07 | 0.01 |
| Antiinflammatory and antirheumatic products | 2041 (33.9) | 96 (27.5) | 634 (28.4) | **-0.14** | **-0.12** |
| Antineoplastic agents | 133 (2.2) | 6 (1.7) | 47 (2.1) | -0.04 | -0.01 |
| Antipsoriatics | 1243 (20.6) | 74 (21.2) | 503 (22.5) | 0.01 | 0.05 |
| Antithrombotic agents | 1807 (30) | 105 (30.1) | 681 (30.5) | 0.00 | 0.01 |
| Beta blocking agents | 1393 (23.1) | 74 (21.2) | 489 (21.9) | -0.05 | -0.03 |
| Calcium channel blockers | 1480 (24.6) | 85 (24.4) | 525 (23.5) | 0.00 | -0.02 |
| Diuretics | 1398 (23.2) | 83 (23.8) | 444 (19.9) | 0.01 | -0.08 |
| Drugs for acid related disorders | 2912 (48.3) | 159 (45.6) | 1076 (48.2) | -0.06 | 0.00 |
| Drugs for obstructive airway diseases | 1775 (29.5) | 103 (29.5) | 638 (28.6) | 0.00 | -0.02 |
| Drugs used in diabetes | 782 (13) | 50 (14.3) | 326 (14.6) | 0.04 | 0.05 |
| Immunosuppressants | 95 (1.6) | 7 (2) | 39 (1.7) | 0.03 | 0.01 |
| Lipid modifying agents | 2438 (40.5) | 154 (44.1) | 930 (41.6) | 0.07 | 0.02 |
| Opioids | 1865 (31) | 123 (35.2) | 653 (29.2) | 0.09 | -0.04 |
| Psycholeptics | 987 (16.4) | 57 (16.3) | 331 (14.8) | 0.00 | -0.04 |
| Psychostimulants, agents used for adhd and nootropics | <5 (~0) | 0 (0) | <5 (~0) | -0.03 | 0.01 |
| **Comorbidity Scales** | **Mean (SD)** | **Mean (SD)** | **Mean (SD)** | **Std.Diff** | **Std.Diff** |
| Charlson comorbidity index Mean | 3.4 (1.9) | 3.7 (2.3) | 3.5 (2.1) | 0.09 | 0.04 |
| CHADS2Vasc | 2.3 (1.5) | 2.3 (1.5) | 2.3 (1.5) | -0.01 | -0.01 |
| DCSI | 1.3 (1.6) | 1.3 (1.7) | 1.3 (1.6) | 0.02 | 0.01 |

Note. Pre-pandemic = Jan 2017-Feb 2020; During Lockdown = March 2020-June 2020; After lockdown = July 2020-Dec 2021; CHADS2Vasc: Comborbidity scale measuring congestive heart failure, hypertension, age, diabetes, stroke, vascular disease, age and sex. DCSI: Diabetes Complications and Severity Index. SD = Standard Deviation. Counts <5 masked and proportions rounded to nearest 1% in order for frequencies <5 to remain masked. Bold indicates significant standardised mean differences between lockdown periods (>.10).

**Table S11.** Characterization of lung patients diagnosed before, during and after lockdown

| | **Pre-pandemic** | **During Lockdown** | **After Lockdown** | **Standardised Mean Differences** | |
|---|---|---|---|---|---|
| **Characteristic** | **Count (% of n = 5,766)** | **Count (% of n=435)** | **Count (% of n = 2,102)** | **Before vs. during lockdown** | **Before vs. after lockdown** |
| **Medical history: General** | | | | | |
| Acute respiratory disease | 679 (11.8) | 42 (9.7) | 134 (6.4) | -0.07 | **-0.19** |
| Chronic liver disease | 12 (0.2) | <5 (~0) | <5 (~0) | 0.00 | -0.02 |
| Chronic obstructive lung disease | 727 (12.6) | 58 (13.3) | 199 (9.5) | 0.02 | **-0.10** |
| Crohn's disease | 7 (0.1) | 0 (0) | <5 (~0) | -0.05 | -0.03 |
| Dementia | 62 (1.1) | <5 (~0) | 11 (0.5) | **-0.11** | -0.06 |
| Depressive disorder | 82 (1.4) | 7 (1.6) | 19 (0.9) | 0.02 | -0.05 |
| Diabetes mellitus | 129 (2.2) | 7 (1.6) | 46 (2.2) | -0.05 | 0.00 |
| Gastroesophageal reflux disease | 34 (0.6) | <5 (~0) | 14 (0.7) | -0.02 | 0.01 |
| Gastrointestinal hemorrhage | 68 (1.2) | 5 (1.1) | 15 (0.7) | 0.00 | -0.05 |
| Hyperlipidemia | 22 (0.4) | 0 (0) | <5 (~0) | -0.09 | 0.03 |
| Hypertensive disorder | 89 (1.5) | 8 (1.8) | 15 (0.7) | 0.02 | 0.05 |
| Lesion of liver | 144 (2.5) | 12 (2.8) | 27 (1.3) | 0.02 | -0.02 |
| Obesity | 10 (0.2) | 0 (0) | 47 (2.2) | -0.06 | -0.02 |
| Osteoarthritis | 156 (2.7) | 7 (1.6) | <5 (~0) | -0.08 | -0.02 |

**Table S11.** Characterization of lung patients diagnosed before, during and after lockdown

| Characteristic | Pre-pandemic<br>Count (% of n = 5,766) | During Lockdown<br>Count (% of n=435) | After Lockdown<br>Count (% of n = 2,102) | Standardised Mean Differences<br>Before vs. during lockdown | <br>Before vs. after lockdown |
|---|---|---|---|---|---|
| Pneumonia | 219 (3.8) | 24 (5.5) | 52 (2.5) | 0.08 | -0.01 |
| Psoriasis | 30 (0.5) | <5 (~1) | 78 (3.7) | 0.02 | 0.00 |
| Renal impairment | 152 (2.6) | 13 (3) | 14 (0.7) | 0.02 | 0.02 |
| Rheumatoid arthritis | 22 (0.4) | 0 (0) | 59 (2.8) | -0.09 | 0.01 |
| Schizophrenia | <5 (~0) | 0 (0) | 7 (0.3) | -0.03 | -0.01 |
| Ulcerative colitis | <5 (~0) | 0 (0) | 0 (0) | -0.04 | -0.03 |
| Urinary tract infectious disease | 145 (2.5) | 7 (1.6) | <5 (~0) | -0.06 | 0.01 |
| Viral hepatitis C | 0 (0) | <5 (~0) | 38 (1.8) | 0.07 | -0.05 |
| Visual system disorder | 420 (7.3) | 29 (6.7) | 105 (5) | -0.02 | **-0.10** |
| **Medical history: Cardiovascular disease** | | | | | |
| Atrial fibrillation | 181 (3.1) | 18 (4.1) | 61 (2.9) | 0.05 | -0.01 |
| Cerebrovascular disease | 104 (1.8) | 10 (2.3) | 44 (2.1) | 0.03 | 0.02 |
| Coronary arteriosclerosis | 11 (0.2) | <5 (~1) | <5 (~0) | 0.05 | 0.00 |
| Heart disease | 421 (7.3) | 36 (8.3) | 179 (8.5) | 0.04 | 0.05 |
| Heart failure | 76 (1.3) | 6 (1.4) | 37 (1.8) | 0.01 | 0.04 |
| Ischemic heart disease | 109 (1.9) | 8 (1.8) | 49 (2.3) | 0.00 | 0.03 |
| Peripheral vascular disease | 59 (1) | 6 (1.4) | 19 (0.9) | 0.03 | -0.01 |
| Pulmonary embolism | 120 (2.1) | 13 (3) | 46 (2.2) | 0.06 | 0.01 |

**Table S11.** Characterization of lung patients diagnosed before, during and after lockdown

| | Pre-pandemic | During Lockdown | After Lockdown | Standardised Mean Differences | |
|---|---|---|---|---|---|
| **Characteristic** | **Count (% of n = 5,766)** | **Count (% of n=435)** | **Count (% of n = 2,102)** | **Before vs. during lockdown** | **Before vs. after lockdown** |
| Venous thrombosis | 79 (1.4) | <5 (~1) | 20 (1) | -0.07 | -0.04 |
| **Medical history: Neoplasms** | | | | | |
| Hematologic neoplasm | 43 (0.7) | 5 (1.1) | 13 (0.6) | 0.04 | -0.02 |
| Malignant lymphoma | 10 (0.2) | <5 (~0) | 9 (0.4) | 0.01 | 0.05 |
| Malignant neoplasm of anorectum | 14 (0.2) | <5 (~1) | 10 (0.5) | 0.09 | 0.04 |
| Malignant neoplastic disease | 5766 (100) | 435 (100) | 2102 (100) | / | / |
| Malignant tumor of breast | 33 (0.6) | <5 (~0) | 17 (0.8) | -0.02 | 0.03 |
| Malignant tumor of colon | 29 (0.5) | <5 (~0) | 11 (0.5) | -0.01 | 0.00 |
| Malignant tumor of lung | 5766 (100) | 435 (100) | 2102 (100) | / | / |
| Malignant tumor of urinary bladder | 16 (0.3) | 5 (1.1) | 9 (0.4) | 0.10 | 0.03 |
| **Medication use** | | | | | |
| Agents acting on the renin-angiotensin system | 2075 (36) | 146 (33.6) | 731 (34.8) | -0.05 | -0.03 |
| Antibacterials for systemic use | 3955 (68.6) | 292 (67.1) | 1321 (62.8) | -0.03 | **-0.12** |
| Antidepressants | 1819 (31.5) | 151 (34.7) | 702 (33.4) | 0.07 | 0.04 |
| Antiepileptics | 776 (13.5) | 46 (10.6) | 275 (13.1) | -0.09 | -0.01 |
| Antiinflammatory and antirheumatic products | 2775 (48.1) | 200 (46) | 905 (43.1) | -0.04 | **-0.10** |

**Table S11.** Characterization of lung patients diagnosed before, during and after lockdown

| | Pre-pandemic | During Lockdown | After Lockdown | Standardised Mean Differences | |
|---|---|---|---|---|---|
| **Characteristic** | **Count (% of n = 5,766)** | **Count (% of n=435)** | **Count (% of n = 2,102)** | **Before vs. during lockdown** | **Before vs. after lockdown** |
| Antineoplastic agents | 160 (2.8) | 15 (3.4) | 67 (3.2) | 0.04 | 0.02 |
| Antipsoriatics | 490 (8.5) | 38 (8.7) | 232 (11) | 0.01 | 0.09 |
| Antithrombotic agents | 2434 (42.2) | 175 (40.2) | 880 (41.9) | -0.04 | -0.01 |
| Beta blocking agents | 1402 (24.3) | 109 (25.1) | 524 (24.9) | 0.02 | 0.01 |
| Calcium channel blockers | 1564 (27.1) | 111 (25.5) | 574 (27.3) | -0.04 | 0.00 |
| Diuretics | 1507 (26.1) | 104 (23.9) | 502 (23.9) | -0.05 | -0.05 |
| Drugs for acid related disorders | 3416 (59.2) | 260 (59.8) | 1305 (62.1) | 0.01 | 0.06 |
| Drugs for obstructive airway diseases | 3132 (54.3) | 231 (53.1) | 1118 (53.2) | -0.02 | -0.02 |
| Drugs used in diabetes | 774 (13.4) | 47 (10.8) | 301 (14.3) | -0.08 | 0.03 |
| Immunosuppressants | 138 (2.4) | 11 (2.5) | 60 (2.9) | 0.01 | 0.03 |
| Lipid modifying agents | 2909 (50.5) | 216 (49.7) | 1101 (52.4) | -0.02 | 0.04 |
| Opioids | 3014 (52.3) | 241 (55.4) | 1059 (50.4) | 0.06 | -0.04 |
| Psycholeptics | 1441 (25) | 112 (25.7) | 521 (24.8) | 0.02 | 0.00 |
| **Comorbidity Scales** | **Mean (SD)** | **Mean (SD)** | **Mean (SD)** | **Std.Diff** | **Std.Diff** |
| Charlson comorbidity index | 4.0 (2.2) | 4.0 (2.2) | 4.1 (2.3) | 0.00 | 0.05 |
| CHADS2Vasc | 2.6 (1.5) | 2.7 (1.5) | 2.7 (1.5) | 0.03 | 0.02 |
| DCSI | 1.6 (1.7) | 1.6 (1.7) | 1.6 (1.6) | 0.03 | 0.02 |

Note. Pre-pandemic = Jan 2017-Feb 2020; During Lockdown = March 2020-June 2020; After lockdown = July 2020-Dec 2021; CHADS2Vasc: Comborbidity scale measuring congestive heart failure, hypertension, age, diabetes, stroke, vascular disease, age and sex. DCSI: Diabetes Complications and Severity Index. SD = Standard Deviation. Counts <5 masked and proportions rounded to nearest 1% in order for frequencies <5 to remain masked. Bold indicates significant standardised mean differences between lockdown periods (>.10).

**Table S12.** Characterisation of prostate cancer patients diagnosed before, during and after lockdown

| | Pre-pandemic | During Lockdown | After Lockdown | Standardised Mean Differences | |
|---|---|---|---|---|---|
| **Characteristic** | **Count (% of n=8,101)** | **Count (% of n=427)** | **Count of % n=2,476)** | **Before vs. during lockdown** | **Before vs. after lockdown** |
| **Medical history: General** | | | | | |
| Acute respiratory disease | 274 (3.4) | 14 (3.3) | 46 (1.9) | -0.01 | **-0.10** |
| Attention deficit hyperactivity disorder | <5 (~0) | 0 (0) | 0 (0) | -0.02 | -0.02 |
| Chronic liver disease | <5 (~0) | <5 (~1) | 5 (0.2) | 0.08 | 0.04 |
| Chronic obstructive lung disease | 176 (2.2) | 5 (1.2) | 26 (1) | -0.08 | -0.09 |
| Crohn's disease | 5 (0.1) | 0 (0) | <5 (~0) | -0.04 | -0.01 |
| Dementia | 35 (0.4) | 0 (0) | 10 (0.4) | -0.09 | 0.00 |
| Depressive disorder | 69 (0.9) | <5 (~1) | 10 (0.4) | -0.05 | -0.06 |
| Diabetes mellitus | 169 (2.1) | 16 (3.7) | 33 (1.3) | **0.10** | -0.06 |
| Gastroesophageal reflux disease | 37 (0.5) | 0 (0) | 13 (0.5) | **-0.10** | 0.01 |
| Gastrointestinal hemorrhage | 81 (1) | <5 (~1) | 16 (0.6) | -0.03 | -0.04 |
| Human immunodeficiency virus infection | <5 (~0) | 0 (0) | 0 (0) | -0.02 | -0.02 |
| Hyperlipidemia | 55 (0.7) | <5 (~0) | 8 (0.3) | -0.07 | -0.05 |
| Hypertensive disorder | 223 (2.8) | 10 (2.3) | 46 (1.9) | -0.03 | -0.06 |
| Lesion of liver | 15 (0.2) | 5 (1.2) | 16 (0.6) | **0.12** | 0.07 |
| Obesity | 12 (0.1) | 0 (0) | 6 (0.2) | -0.05 | 0.02 |
| Osteoarthritis | 234 (2.9) | 11 (2.6) | 54 (2.2) | -0.02 | -0.05 |
| Pneumonia | 51 (0.6) | <5 (~1) | 13 (0.5) | -0.02 | -0.01 |
| Psoriasis | 39 (0.5) | 0 (0) | <5 (~0) | **-0.10** | -0.07 |

| **Table S12.** Characterisation of prostate cancer patients diagnosed before, during and after lockdown | | | | | |
|---|---|---|---|---|---|
| | **Pre-pandemic** | **During Lockdown** | **After Lockdown** | **Standardised Mean Differences** | |
| **Characteristic** | **Count (% of n=8,101)** | **Count (% of n=427)** | **Count of % n=2,476)** | **Before vs. during lockdown** | **Before vs. after lockdown** |
| Renal impairment | 258 (3.2) | 9 (2.1) | 63 (2.5) | -0.07 | -0.04 |
| Rheumatoid arthritis | 8 (0.1) | <5 (~0) | <5 (~0) | 0.03 | -0.02 |
| Ulcerative colitis | <5 (~0) | 0 (0) | <5 (~0) | -0.02 | 0.01 |
| Urinary tract infectious disease | 227 (2.8) | 12 (2.8) | 58 (2.3) | 0.00 | -0.03 |
| Visual system disorder | 482 (5.9) | 28 (6.6) | 104 (4.2) | 0.03 | -0.08 |
| **Medical history: Cardiovascular disease** | | | | | |
| Atrial fibrillation | 161 (2) | 10 (2.3) | 35 (1.4) | 0.02 | -0.04 |
| Cerebrovascular disease | 76 (0.9) | 5 (1.2) | 18 (0.7) | 0.02 | -0.02 |
| Coronary arteriosclerosis | 13 (0.2) | <5 (~0) | <5 (~0) | 0.02 | -0.01 |
| Heart disease | 351 (4.3) | 18 (4.2) | 92 (3.7) | -0.01 | -0.03 |
| Heart failure | 66 (0.8) | <5 (~1) | 12 (0.5) | 0.01 | -0.04 |
| Ischemic heart disease | 78 (1) | <5 (~1) | 25 (1) | -0.03 | 0.00 |
| Peripheral vascular disease | 29 (0.4) | <5 (~1) | 5 (0.2) | 0.02 | -0.03 |
| Pulmonary embolism | 31 (0.4) | <5 (~1) | 17 (0.7) | 0.01 | 0.04 |
| Venous thrombosis | 72 (0.9) | 8 (1.9) | 23 (0.9) | 0.08 | 0.00 |
| **Medical history: Neoplasms** | | | | | |
| Hematologic neoplasm | 30 (0.4) | <5 (~1) | 10 (0.4) | 0.07 | 0.01 |
| Malignant lymphoma | 5 (0.1) | 0 (0) | <5 (~0) | -0.04 | 0.01 |
| Malignant neoplasm of anorectum | 15 (0.2) | <5 (~1) | 6 (0.2) | 0.05 | 0.01 |

**Table S12.** Characterisation of prostate cancer patients diagnosed before, during and after lockdown

| | Pre-pandemic | During Lockdown | After Lockdown | Standardised Mean Differences | |
|---|---|---|---|---|---|
| **Characteristic** | **Count (% of n=8,101)** | **Count (% of n=427)** | **Count of % n=2,476)** | **Before vs. during lockdown** | **Before vs. after lockdown** |
| Malignant neoplastic disease | 8103 (100) | 427 (100) | 2477 (100) | / | / |
| Malignant tumor of breast | 0 (0) | 0 (0) | 0 (0) | -0.03 | -0.03 |
| Malignant tumor of colon | 10 (0.1) | 0 (0) | 7 (0.3) | -0.05 | 0.04 |
| Malignant tumor of lung | 17 (0.2) | <5 (~1) | 7 (0.3) | 0.04 | 0.01 |
| Malignant tumor of urinary bladder | 47 (0.6) | 5 (1.2) | 18 (0.7) | 0.06 | 0.02 |
| **Medication use** | | | | | |
| Agents acting on the renin-angiotensin system | 3122 (38.5) | 161 (37.7) | 891 (36) | -0.02 | -0.05 |
| Antibacterials for systemic use | 3682 (45.4) | 195 (45.7) | 1064 (43) | 0.00 | -0.05 |
| Antidepressants | 1162 (14.3) | 54 (12.6) | 400 (16.1) | -0.05 | 0.05 |
| Antiepileptics | 475 (5.9) | 31 (7.3) | 161 (6.5) | 0.06 | 0.03 |
| Antiinflammatory and antirheumatic products | 3068 (37.9) | 141 (33) | 884 (35.7) | **-0.10** | -0.05 |
| Antineoplastic agents | 201 (2.5) | 15 (3.5) | 73 (2.9) | 0.06 | 0.03 |
| Antipsoriatics | 402 (5) | 19 (4.4) | 121 (4.9) | -0.02 | 0.00 |
| Antithrombotic agents | 2622 (32.4) | 128 (30) | 829 (33.5) | -0.05 | 0.02 |
| Beta blocking agents | 1806 (22.3) | 106 (24.8) | 532 (21.5) | 0.06 | -0.02 |
| Calcium channel blockers | 2304 (28.4) | 118 (27.6) | 673 (27.2) | -0.02 | -0.03 |
| Diuretics | 1425 (17.6) | 66 (15.5) | 394 (15.9) | -0.06 | -0.04 |

**Table S12.** Characterisation of prostate cancer patients diagnosed before, during and after lockdown

| | Pre-pandemic | During Lockdown | After Lockdown | Standardised Mean Differences | |
|---|---|---|---|---|---|
| **Characteristic** | **Count (% of n=8,101)** | **Count (% of n=427)** | **Count of % n=2,476)** | **Before vs. during lockdown** | **Before vs. after lockdown** |
| Drugs for acid related disorders | 3317 (40.9) | 170 (39.8) | 1060 (42.8) | -0.02 | 0.04 |
| Drugs for obstructive airway diseases | 2352 (29) | 106 (24.8) | 700 (28.3) | -0.09 | -0.02 |
| Drugs used in diabetes | 891 (11) | 58 (13.6) | 273 (11) | 0.08 | 0.00 |
| Immunosuppressants | 113 (1.4) | 9 (2.1) | 31 (1.3) | 0.05 | -0.01 |
| Lipid modifying agents | 3726 (46) | 201 (47.1) | 1152 (46.5) | 0.02 | 0.01 |
| Opioids | 2111 (26.1) | 125 (29.3) | 667 (26.9) | 0.07 | 0.02 |
| Psycholeptics | 830 (10.2) | 46 (10.8) | 279 (11.3) | 0.02 | 0.03 |
| Psychostimulants, agents used for adhd and nootropics | <5 (~0) | 0 (0) | 0 (0) | -0.02 | -0.02 |
| **Comorbidity Scales** | **Mean (SD)** | **Mean (SD)** | **Mean (SD)** | **Std.Diff** | **Std.Diff** |
| Charlson comorbidity index | 3.2 (1.7) | 3.2 (1.7) | 3.2 (1.7) | 0.03 | 0.03 |
| CHADS2Vasc | 1.9 (1.3) | 1.8 (1.3) | 1.9 (1.3) | -0.01 | 0.02 |
| DCSI | 1.2 (1.5) | 1.2 (1.4) | 1.1 (1.5) | 0.02 | -0.02 |

Note. Pre-pandemic = Jan 2017-Feb 2020; During Lockdown = March 2020-June 2020; After lockdown = July 2020-Dec 2021; CHADS2Vasc: Comborbidity scale measuring congestive heart failure, hypertension, age, diabetes, stroke, vascular disease, age and sex. DCSI: Diabetes Complications and Severity Index. SD = Standard Deviation. Counts <5 masked and proportions rounded to nearest 1% in order for frequencies <5 to remain masked. Bold indicates significant standardised mean differences between lockdown periods (>.10).

**Table S13.** Incidence rates (with 95% confidence intervals) of screening and diagnostic tests across extended lockdown periods

| Screening/Diagnostic Test | Pre-pandemic (Jan 2017-Feb 2020) | Lockdown (March 2020-June 2020) | Post-lockdown (July 2020-Dec 2021) | Post-first lockdown 1 (July 2020-Oct 2020) | Second lockdown (Nov 2020-Dec 2020) | Third lockdown (Jan 2021-Feb 2021) | Easing of restrictions (March 2021-June 2021 | Legal restrictions removed (July 2021-Dec 2021) |
|---|---|---|---|---|---|---|---|---|
| Biopsy of Breast | 2.5 (2.4 to 2.6) | 1.9 (1.7 to 2.1) | 2.9 (2.8 to 3.1) | 2.3 (2 to 2.6) | 2.5 (2.2 to 3) | 3 (2.5 to 3.4) | 3.4 (3.1 to 3.7) | 3.3 (3 to 3.6) |
| Biopsy Of Prostate | 1.7 (1.7 to 1.8) | 1.2 (1 to 1.4) | 1.7 (1.5 to 1.8) | 1.2 (1 to 1.4) | 1.9 (1.5 to 2.2) | 1.7 (1.4 to 2) | 1.9 (1.6 to 2.1) | 1.8 (1.6 to 2) |
| Bowel Cancer Screening Prog | 0.2 (0.1 to 0.2) | NR | 0.2 (0.2 to 0.3) | NR | NR | 0.2 (0.1 to 0.3) | 0.2 (0.1 to 0.4) | 0.3 (0.2 to 0.5) |
| Breast Cancer Referrals | 27.3 (27 to 27.6) | 17.5 (16.8 to 18.3) | 25.1 (24.7 to 25.6) | 24.8 (24 to 25.7) | 25.7 (24.4 to 26.9) | 23.6 (22.4 to 24.8) | 26.1 (25.2 to 27) | 25 (24.2 to 25.8) |
| Bronchoscopy | 0.2 (0.1 to 0.2) | NR | NR | NR | NR | NR | NR | NR |
| Colonoscopy | 35.7 (35.4 to 36) | 12.6 (12 to 13.2) | 26.3 (25.9 to 26.7) | 20.2 (19.4 to 21) | 26.1 (24.8 to 27.4) | 27.2 (25.9 to 28.5) | 30.1 (29.1 to 31) | 28.3 (27.5 to 29.1) |
| Colorectal Cancer Referrals | 19.9 (19.7 to 20.1) | 11.5 (11 to 12.1) | 18.5 (18.2 to 18.9) | 17.7 (17 to 18.5) | 17.7 (16.7 to 18.8) | 16.5 (15.5 to 17.6) | 20.4 (19.6 to 21.2) | 19 (18.3 to 19.7) |
| Diagnostic Procedures Of Chest | 0.2 (0.1 to 0.2) | NR | 0.2 (0.1 to 0.4) | NR | NR | NR | NR | 0.2 (0.1 to 0.4) |
| Excision Of Breast | 4.9 (4.7 to 5) | 3.5 (3.2 to 3.8) | 4.2 (4.1 to 4.4) | 3.6 (3.2 to 3.9) | 5 (4.4 to 5.6) | 3.9 (3.4 to 4.4) | 4.5 (4.1 to 4.9) | 4.5 (4.2 to 4.9) |
| Lung Cancer Referrals | 1.2 (1.1 to 1.2) | 0.6 (0.5 to 0.7) | 0.8 (0.7 to 0.9) | 0.8 (0.7 to 1) | 0.7 (0.5 to 0.9) | 0.8 (0.6 to 1) | 1 (0.8 to 1.2) | 0.8 (0.6 to 0.9) |
| Mammograms | 79.1 (78.6 to 79.6) | 18 (17.3 to 18.8) | 59.4 (58.7 to 60) | 51.9 (50.6 to 53.1) | 58.8 (57 to 60.7) | 77.3 (75.2 to 79.6) | 60 (58.6 to 61.3) | 58.4 (57.2 to 59.6) |
| Prostate Specific Antigen Test | 67.6 (67.2 to 68) | 29.2 (28.3 to 30.1) | 53 (52.3 to 53.6) | 45.1 (43.9 to 46.2) | 51.7 (50 to 53.5) | 47 (45.3 to 48.7) | 59.2 (57.8 to 60.6) | 57.6 (56.4 to 58.8) |
| Seen Breast Surgeon | 1.5 (1.5 to 1.6) | 0.6 (0.4 to 0.7) | 0.4 (0.4 to 0.5) | 0.4 (0.3 to 0.5) | 0.5 (0.3 to 0.7) | 0.4 (0.3 to 0.6) | 0.5 (0.4 to 0.6) | 0.3 (0.2 to 0.4) |
| Seen in Breast Clinic | 26.5 (26.2 to 26.8) | 14.3 (13.6 to 14.9) | 21.8 (21.4 to 22.2) | 19.3 (18.5 to 20) | 24 (22.8 to 25.3) | 19 (17.9 to 20.1) | 24.6 (23.8 to 25.5) | 21.9 (21.2 to 22.6) |

Note. NR = counts <5 censored and so not reported.

**Table S14.** Incidence rate ratios (with 95% confidence intervals) of screening and diagnostic tests compared to pre-pandemic period

| Screening/Diagnostic Test | Lockdown (March 2020-June 2020) | Post-lockdown1 (July 2020-Oct 2020) | Second lockdown (Nov 2020-Dec 2020) | Third lockdown (Jan 2021-Feb 2021) | Easing of restrictions (March 2021-June 2021) | Legal restrictions removed (July 2021-Dec 2021) |
|---|---|---|---|---|---|---|
| Biopsy of Breast | 0.76 (0.67 to 0.86) | 0.91 (0.81 to 1.02) | 1.01 (0.86 to 1.18) | 1.18 (1.02 to 1.37) | 1.35 (1.21 to 1.49) | 1.32 (1.2 to 1.44) |
| Biopsy of Prostate | 0.69 (0.59 to 0.81) | 0.68 (0.58 to 0.8) | 1.08 (0.89 to 1.29) | 0.98 (0.8 to 1.19) | 1.08 (0.94 to 1.23) | 1.05 (0.93 to 1.19) |
| Breast Cancer Referrals | 0.64 (0.62 to 0.67) | 0.91 (0.88 to 0.94) | 0.94 (0.9 to 0.99) | 0.86 (0.82 to 0.91) | 0.96 (0.92 to 0.99) | 0.92 (0.89 to 0.95) |
| Colonoscopy | 0.35 (0.34 to 0.37) | 0.57 (0.54 to 0.59) | 0.73 (0.7 to 0.77) | 0.76 (0.72 to 0.8) | 0.84 (0.81 to 0.87) | 0.79 (0.77 to 0.82) |
| Colorectal Cancer Referrals | 0.58 (0.55 to 0.61) | 0.89 (0.85 to 0.93) | 0.89 (0.84 to 0.94) | 0.83 (0.78 to 0.88) | 1.02 (0.98 to 1.06) | 0.95 (0.92 to 0.99) |
| Excision of Breast | 0.71 (0.65 to 0.78) | 0.73 (0.67 to 0.8) | 1.02 (0.91 to 1.14) | 0.79 (0.7 to 0.9) | 0.92 (0.84 to 1) | 0.93 (0.86 to 1) |
| Lung Cancer Referrals | 0.51 (0.4 to 0.63) | 0.69 (0.56 to 0.83) | 0.58 (0.42 to 0.77) | 0.63 (0.47 to 0.84) | 0.81 (0.67 to 0.97) | 0.64 (0.53 to 0.77) |
| Mammograms | 0.23 (0.22 to 0.24) | 0.66 (0.64 to 0.67) | 0.74 (0.72 to 0.77) | 0.98 (0.95 to 1.01) | 0.76 (0.74 to 0.78) | 0.74 (0.72 to 0.75) |
| PSA | 0.43 (0.42 to 0.45) | 0.67 (0.65 to 0.68) | 0.77 (0.74 to 0.79) | 0.69 (0.67 to 0.72) | 0.88 (0.86 to 0.9) | 0.85 (0.83 to 0.87) |
| Seen in Breast Clinic | 0.54 (0.51 to 0.56) | 0.73 (0.7 to 0.76) | 0.91 (0.86 to 0.95) | 0.72 (0.67 to 0.76) | 0.93 (0.9 to 0.96) | 0.83 (0.8 to 0.86) |
| Seen by Breast Surgeon | 0.37 (0.29 to 0.46) | 0.26 (0.19 to 0.34) | 0.32 (0.22 to 0.45) | 0.29 (0.19 to 0.42) | 0.31 (0.23 to 0.4) | 0.2 (0.14 to 0.27) |
| Sigmoidoscopy | 0.3 (0.28 to 0.32) | 0.42 (0.4 to 0.45) | 0.62 (0.58 to 0.67) | 0.64 (0.6 to 0.69) | 0.64 (0.61 to 0.67) | 0.51 (0.48 to 0.53) |

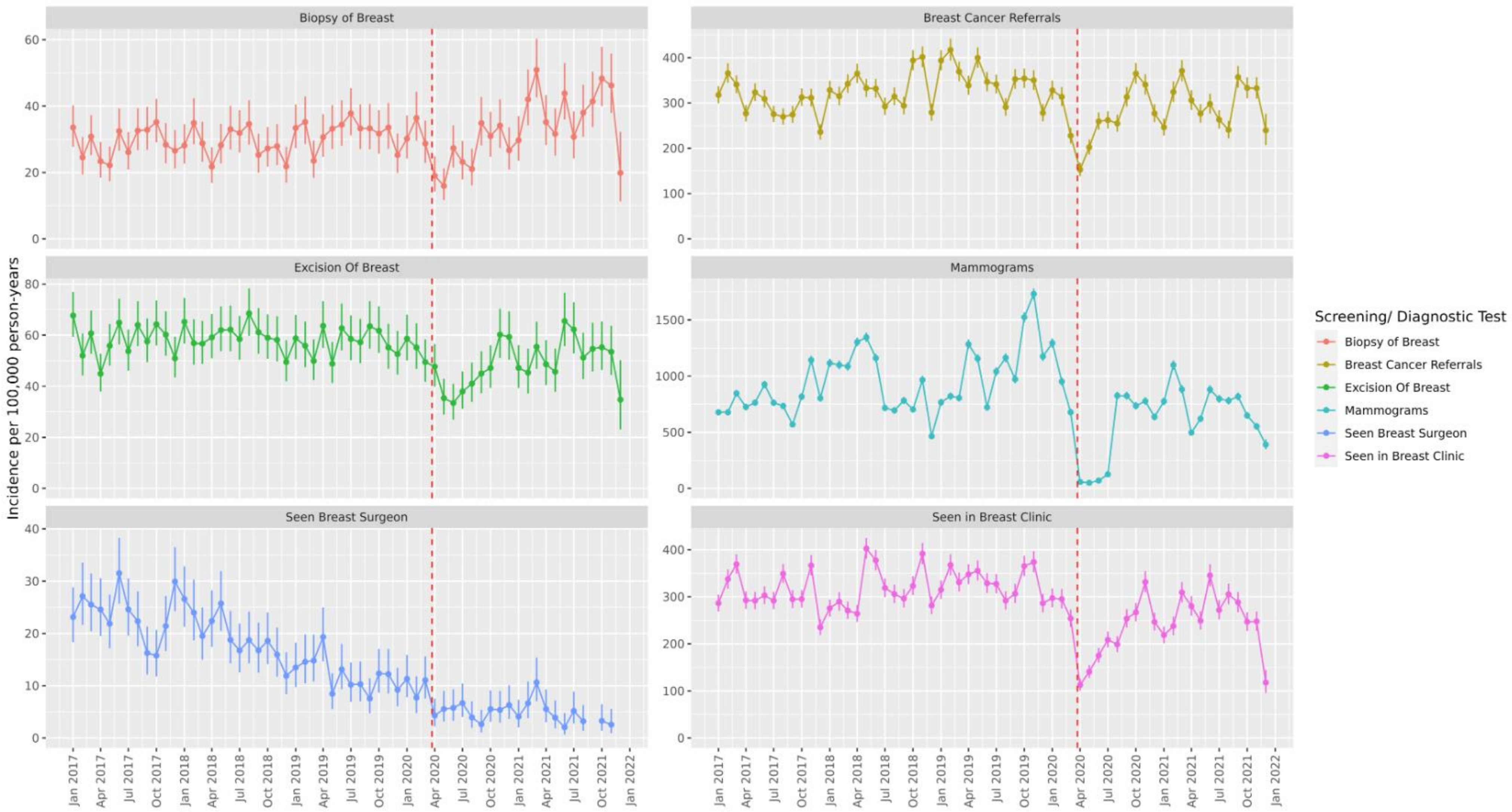


**Figure S4.** Incidence Rates (with 95% confidence intervals) per 100,000 Person-Years of Breast Cancer Screening/ Diagnostic Tests/ Referrals in Months Before and After COVID-19 Lockdown (red dashed line)

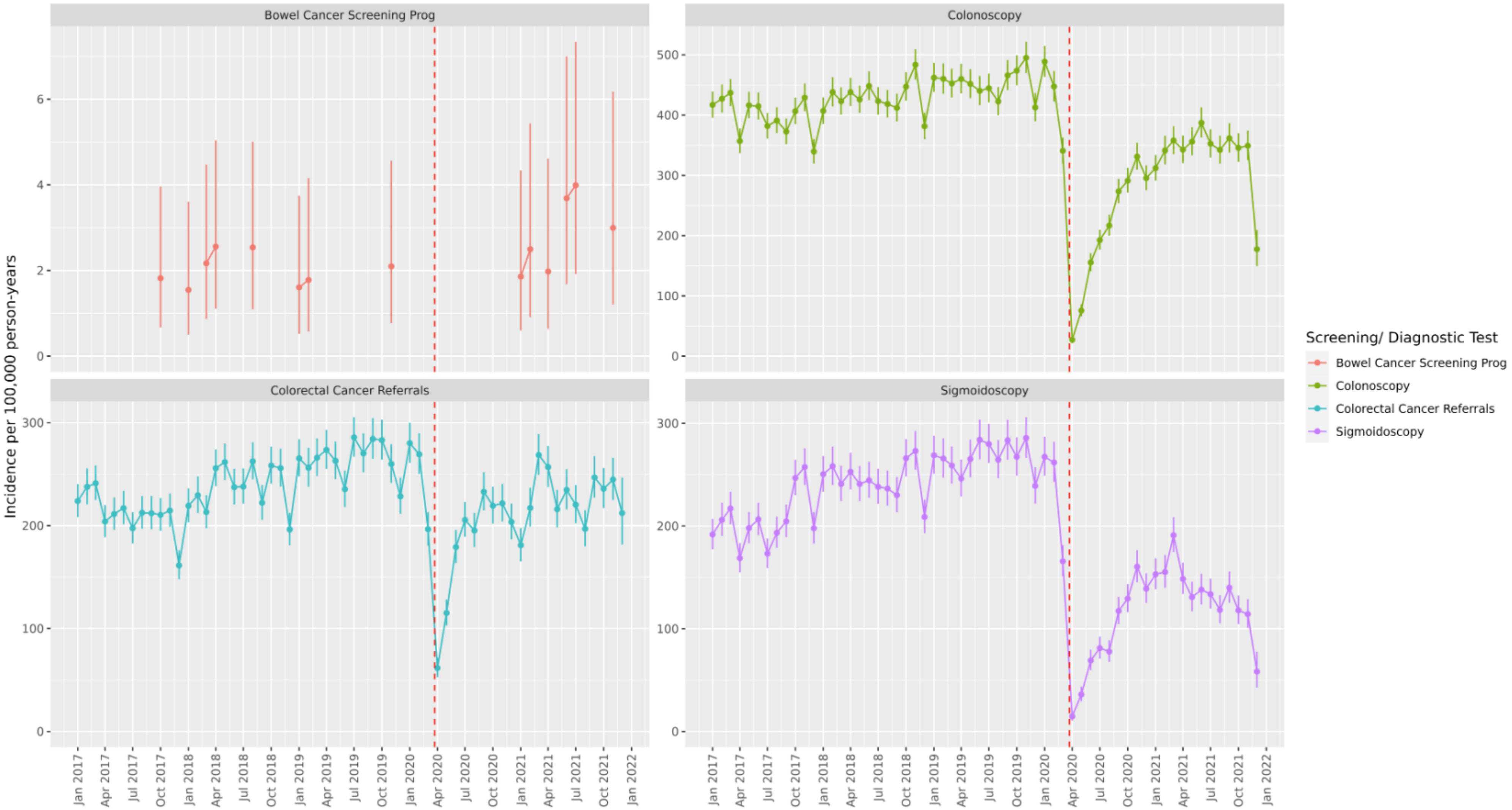


**Figure S5.** Incidence Rates (with 95% confidence intervals) per 100,000 Person-Years of Colorectal Cancer Screening/ Diagnostic Tests/ Referrals in Months Before and After COVID-19 Lockdown (red dashed line)

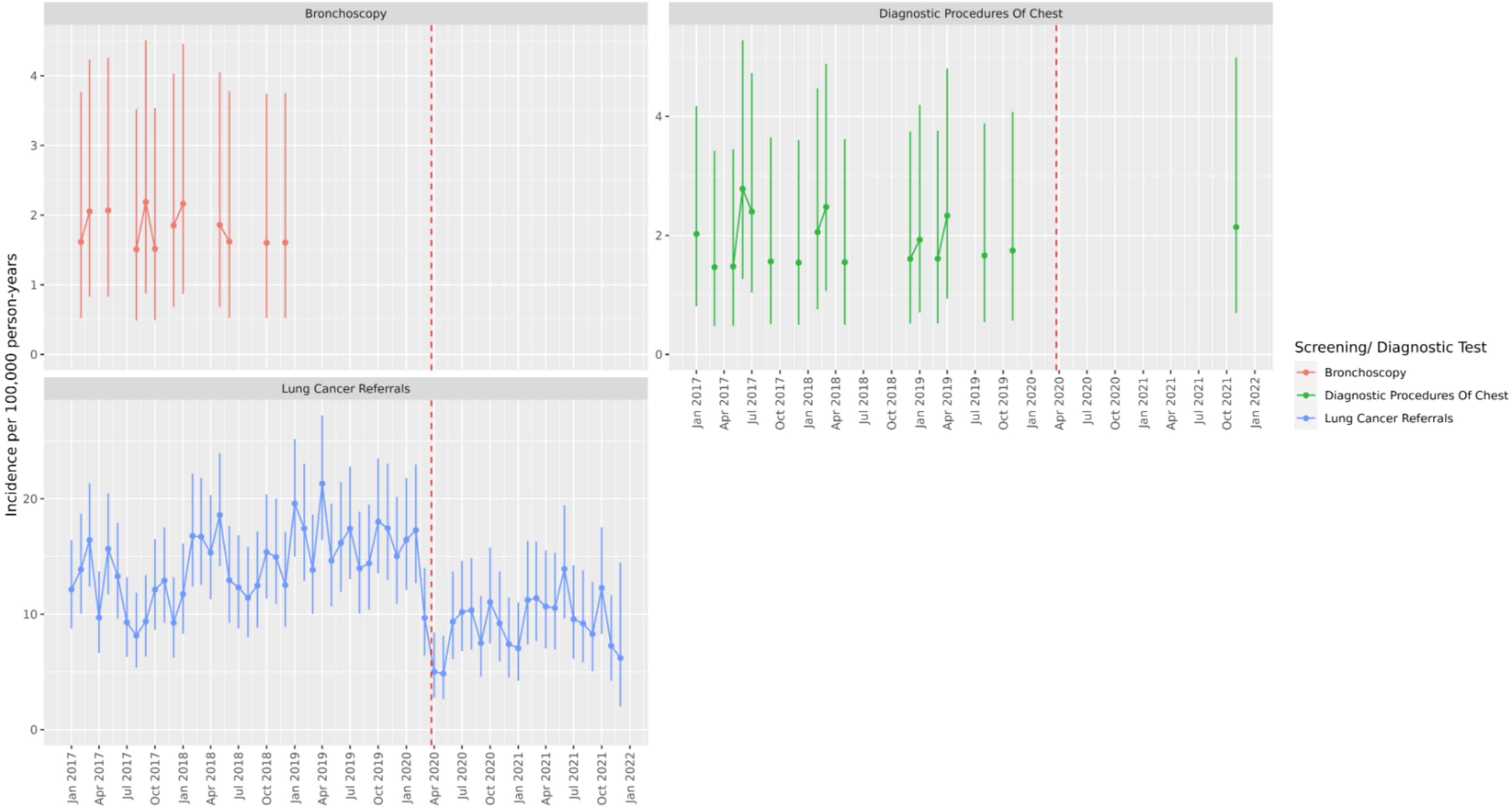


**Figure S6.** Incidence Rates (with 95% confidence intervals) per 100,000 Person-Years of Lung Cancer Screening/ Diagnostic Tests/ Referrals in Months Before and After COVID-19 Lockdown (red dashed line)

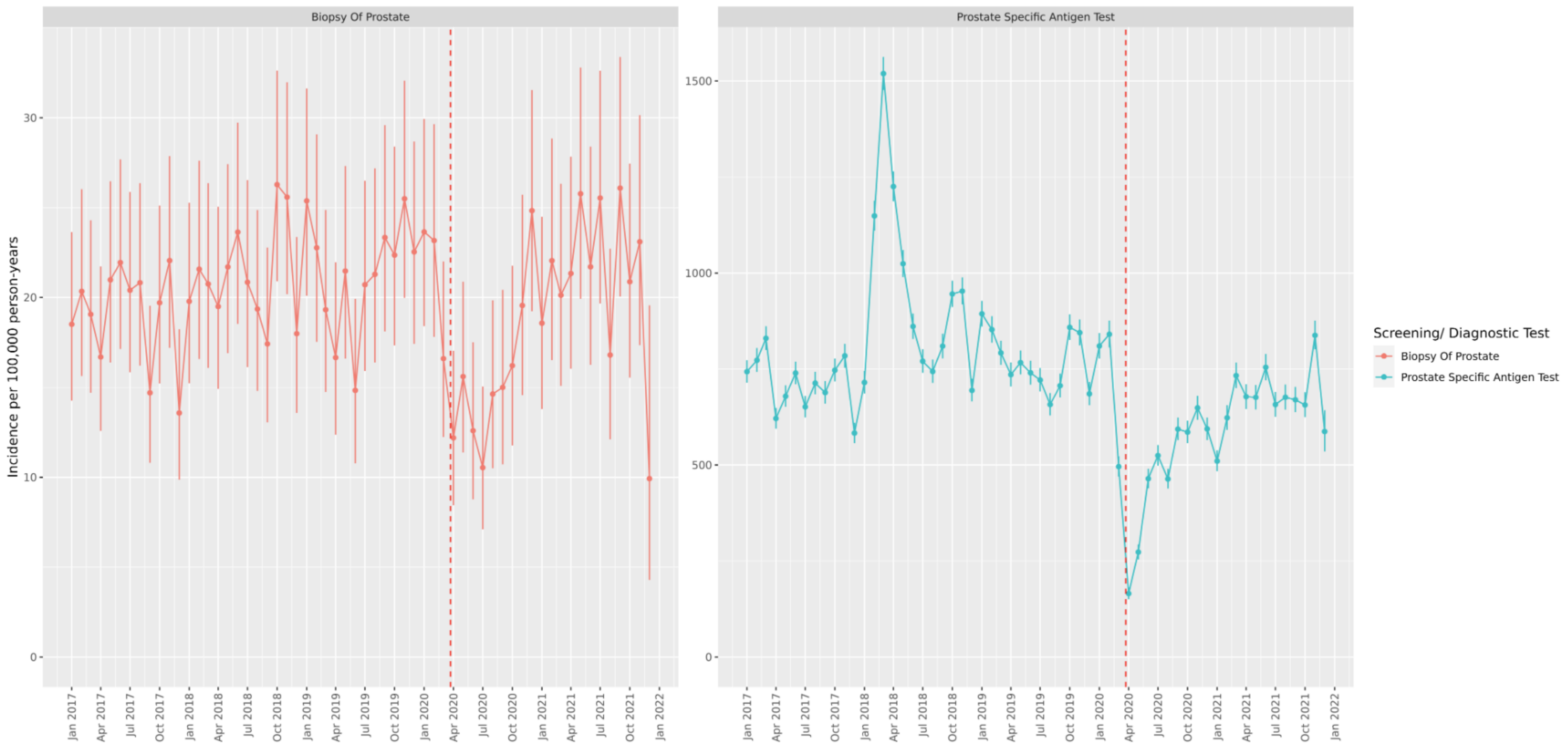


**Figure S7.** Incidence Rates (with 95% confidence intervals) per 100,000 Person-Years of Prostate Cancer Screening/ Diagnostic Tests/ Referrals in Months Before and After COVID-19 Lockdown (red dashed line)

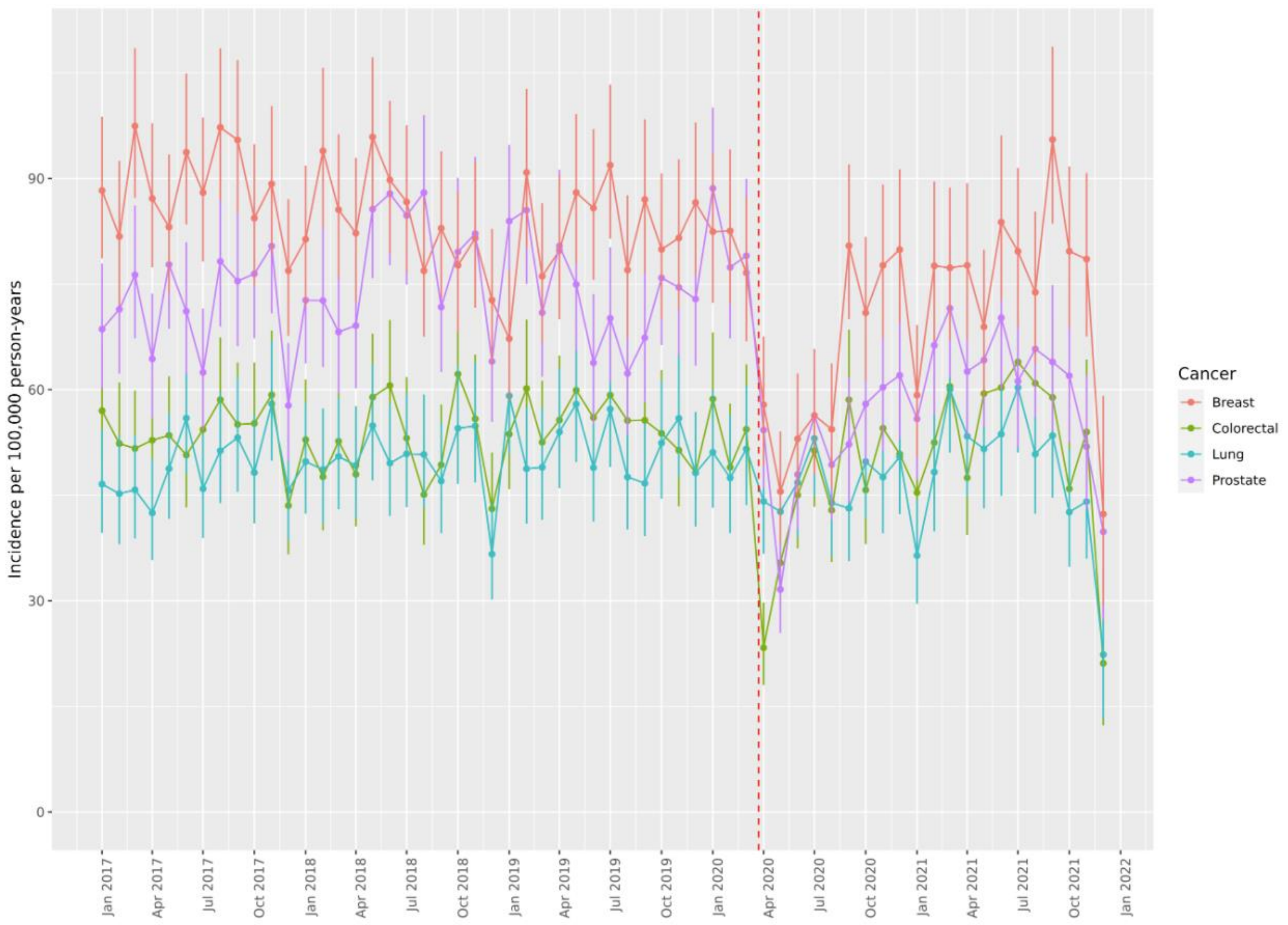


**Figure S8:** Incidence Rates per 100,000 Person-Years (with 95% confidence intervals) of Cancer Diagnoses in Months Before and After COVID-19 Lockdown (red dashed line)

**Table S15.** Incidence Rate Ratios (with 95% confidence intervals) for the cancer diagnoses, in each lockdown period, with the pre-pandemic era (Jan 2017 to Feb 2020) as a reference

| Cancer | First Lockdown (March 2020-June 2020) | Post-first lockdown 1 (July 2020-Oct 2020) | Second lockdown (Nov 2020-Dec 2020) | Third lockdown (Jan 2021-Feb 2021) | Easing of restrictions (March 2021-June 2021) | Legal restrictions removed (July 2021-Dec 2021) |
|---|---|---|---|---|---|---|
| Breast | 0.69 (0.63 to 0.74) | 0.77 (0.71 to 0.83) | 0.93 (0.84 to 1.02) | 0.8 (0.72 to 0.89) | 0.9 (0.84 to 0.97) | 0.93 (0.87 to 0.99) |
| Colorectal | 0.74 (0.67 to 0.81) | 0.92 (0.84 to 1.01) | 0.98 (0.87 to 1.1) | 0.91 (0.8 to 1.03) | 1.06 (0.97 to 1.15) | 1.02 (0.94 to 1.1) |
| Lung | 0.92 (0.84 to 1.01) | 0.95 (0.87 to 1.03) | 0.98 (0.86 to 1.1) | 0.84 (0.73 to 0.96) | 1.09 (1 to 1.19) | 0.97 (0.89 to 1.05) |
| Prostate | 0.71 (0.66 to 0.78) | 0.72 (0.67 to 0.79) | 0.82 (0.73 to 0.92) | 0.82 (0.73 to 0.91) | 0.9 (0.83 to 0.97) | 0.8 (0.74 to 0.86) |

**Table S16.** Observed incidence rates (with 95% confidence intervals) of breast cancer diagnoses in each of the time periods, overall and stratified by age and sex

| Sex | Age Group (years) | Pre-pandemic (Jan 2017-Feb 2020) | First Lockdown (March 2020-June 2020) | Post-lockdown (July 2020-Dec 2021) | Post-first lockdown 1 (July 2020-Oct 2020) | Second lockdown (Nov 2020-Dec 2020) | Third lockdown (Jan 2021-Feb 2021) | Easing of restrictions (March 2021-June 2021 | Legal restrictions removed (July 2021-Dec 2021) |
|---|---|---|---|---|---|---|---|---|---|
| Overall | | 7.1 (6.9 to 7.2) | 4.9 (4.5 to 5.2) | 6.2 (5.9 to 6.4) | 5.4 (5 to 5.9) | 6.6 (6 to 7.2) | 5.7 (5.1 to 6.3) | 6.4 (6 to 6.9) | 6.6 (6.2 to 7) |
| Female | 0;150 | 14 (13.7 to 14.3) | 9.6 (8.9 to 10.4) | 12.2 (11.8 to 12.6) | 10.7 (10 to 11.6) | 13.1 (11.8 to 14.4) | 11.3 (10.1 to 12.5) | 12.7 (11.8 to 13.6) | 13 (12.3 to 13.9) |
| Female | 20;39 | 2.1 (1.9 to 2.4) | 2.1 (1.5 to 2.9) | 2.2 (1.8 to 2.6) | 1.7 (1.1 to 2.6) | 2.8 (1.8 to 4.3) | 1.9 (0.8 to 4) | 2.2 (1.5 to 3.1) | 2.4 (1.6 to 3.3) |
| Female | 40;59 | 19.2 (18.6 to 19.8) | 12.9 (11.3 to 14.6) | 16.4 (15.5 to 17.3) | 15.8 (14 to 17.8) | 18 (15.3 to 21) | 15.1 (12.6 to 18) | 16.2 (14.3 to 18.2) | 16.8 (15.1 to 18.6) |
| Female | 60;79 | 30.7 (29.8 to 31.6) | 19.9 (17.6 to 22.3) | 25.3 (24 to 26.7) | 20.9 (18.6 to 23.5) | 27.6 (23.8 to 31.9) | 24.3 (20.6 to 28.3) | 26.2 (23.5 to 29.1) | 27.8 (25.3 to 30.4) |
| Female | 80;150 | 30.9 (29.2 to 32.6) | 21.6 (17.4 to 26.5) | 28.7 (26.1 to 31.4) | 26.5 (21.7 to 32) | 27.4 (20.5 to 36) | 23.7 (17.4 to 31.5) | 30.9 (25.6 to 36.9) | 31.5 (26.5 to 37.1) |
| Male | 0;150 | 0.3 (0.2 to 0.4) | NR | 0.3 (0.1 to 0.7) | 0.3 (0.1 to 0.7) | NR | NR | NR | NR |

Note. NR = counts <5 censored and so not reported.

**Table S17.** Observed incidence rates (with 95% confidence intervals) of colorectal cancer diagnoses in each of the time periods, overall and stratified by age and sex

| Sex | Age Group (years) | Pre-pandemic (Jan 2017-Feb 2020) | Lockdown (March 2020-June 2020) | Post-lockdown (July 2020-Dec 2021) | Post-first lockdown 1 (July 2020-Oct 2020) | Second lockdown (Nov 2020-Dec 2020) | Third lockdown (Jan 2021-Feb 2021) | Easing of restrictions (March 2021-June 2021 | Legal restrictions removed (July 2021-Dec 2021) |
|---|---|---|---|---|---|---|---|---|---|
| Overall | | 4.5 (4.4 to 4.6) | 3.3 (3 to 3.6) | 4.4 (4.2 to 4.6) | 4.1 (3.8 to 4.5) | 4.4 (3.9 to 4.9) | 4.1 (3.6 to 4.6) | 4.7 (4.4 to 5.1) | 4.5 (4.2 to 4.9) |
| Female | 0;150 | 3.9 (3.7 to 4) | 3 (2.6 to 3.5) | 3.8 (3.5 to 4) | 3.7 (3.3 to 4.2) | 3.4 (2.8 to 4.1) | 2.9 (2.3 to 3.6) | 4.2 (3.7 to 4.7) | 4 (3.6 to 4.5) |
| Female | 20;39 | NR | NR | 1.3 (0.4 to 3.1) | NR | NR | NR | 1.3 (0.4 to 3.1) | NR |
| Female | 40;59 | 2.4 (2.2 to 2.7) | 2.4 (1.7 to 3.2) | 2.2 (1.9 to 2.6) | 2.2 (1.6 to 3) | 1.7 (1 to 2.8) | 2.1 (1.3 to 3.4) | 2.7 (2 to 3.6) | 2.1 (1.5 to 2.9) |
| Female | 60;79 | 10.2 (9.7 to 10.8) | 6.7 (5.5 to 8.2) | 9.4 (8.6 to 10.2) | 9 (7.5 to 10.7) | 8.8 (6.7 to 11.3) | 6.2 (4.4 to 8.4) | 9.7 (8.1 to 11.5) | 11 (9.4 to 12.7) |
| Female | 80;150 | 17.9 (16.7 to 19.2) | 14.1 (10.8 to 18.1) | 17.8 (15.9 to 20) | 18.7 (14.7 to 23.4) | 17.3 (11.9 to 24.3) | 14.5 (9.7 to 20.9) | 18.9 (14.9 to 23.8) | 17.8 (14.2 to 22.1) |
| Male | 0;150 | 5.1 (4.9 to 5.2) | 3.6 (3.2 to 4.1) | 5.1 (4.8 to 5.3) | 4.5 (4 to 5.1) | 5.3 (4.6 to 6.2) | 5.2 (4.5 to 6.1) | 5.3 (4.8 to 5.9) | 5.1 (4.6 to 5.6) |
| Male | 40;59 | 2.8 (2.6 to 3.1) | 2.6 (1.7 to 3.8) | 3 (2.6 to 3.4) | 2.2 (1.6 to 3) | 2.9 (1.9 to 4.2) | 3 (2 to 4.4) | 3.4 (2.6 to 4.3) | 3.6 (2.8 to 4.5) |
| Male | 60;79 | 15.3 (14.6 to 16) | 10.7 (9 to 12.6) | 14.4 (13.4 to 15.4) | 13.1 (11.2 to 15.2) | 16.9 (13.8 to 20.4) | 15.5 (12.6 to 18.9) | 14.7 (12.7 to 17) | 13.7 (11.9 to 15.6) |
| Male | 80;150 | 28.7 (26.8 to 30.7) | 21 (16.1 to 26.9) | 26.7 (23.7 to 29.9) | 29.3 (23.3 to 36.4) | 24.8 (17 to 35) | 22.5 (15.3 to 32) | 28.3 (22.3 to 35.5) | 25.5 (20.2 to 31.7) |

Note. NR = counts <5 censored and so not reported.

**Table S18.** Observed incidence rates (with 95% confidence intervals) of lung cancer diagnoses in each of the time periods, overall and stratified by age and sex

| Sex | Age Group (years) | Pre-pandemic (Jan 2017-Feb 2020) | Lockdown (March 2020-June 2020) | Post-lockdown (July 2020-Dec 2021) | Post-first lockdown 1 (July 2020-Oct 2020) | Second lockdown (Nov 2020-Dec 2020) | Third lockdown (Jan 2021-Feb 2021) | Easing of restrictions (March 2021-June 2021 | Legal restrictions removed (July 2021-Dec 2021) |
|---|---|---|---|---|---|---|---|---|---|
| Overall | | 4.2 (4.1 to 4.3) | 3.9 (3.5 to 4.2) | 4.1 (3.9 to 4.3) | 4 (3.6 to 4.3) | 4.1 (3.6 to 4.6) | 3.5 (3 to 4) | 4.6 (4.2 to 5) | 4.1 (3.7 to 4.4) |
| Female | 0;150 | 4.2 (4.1 to 4.4) | 3.8 (3.4 to 4.3) | 4.1 (3.9 to 4.4) | 4.1 (3.6 to 4.6) | 4.1 (3.4 to 4.8) | 3.6 (3 to 4.4) | 4.5 (4 to 5.1) | 4 (3.6 to 4.5) |
| Female | 40;59 | 1.7 (1.6 to 1.9) | 1.5 (1 to 2.2) | 2 (1.6 to 2.4) | 1.8 (1.2 to 2.7) | 2 (1.2 to 3.2) | 2 (0.9 to 4) | 1.9 (1.2 to 2.8) | 2.2 (1.5 to 3) |
| Female | 60;79 | 13.1 (12.5 to 13.7) | 11.3 (9.6 to 13.2) | 12.3 (11.4 to 13.3) | 12.3 (10.5 to 14.3) | 11.8 (9.3 to 14.7) | 11.3 (8.9 to 14.1) | 13.3 (11.4 to 15.4) | 12.2 (10.6 to 14) |
| Female | 80;150 | 16.8 (15.6 to 18.1) | 15.7 (12.2 to 19.9) | 15.9 (14.1 to 18) | 17.4 (13.6 to 22) | 16.8 (11.5 to 23.7) | 12 (7.7 to 17.9) | 18.9 (14.8 to 23.7) | 13.4 (10.3 to 17.2) |
| Male | 0;150 | 4.2 (4 to 4.3) | 3.9 (3.5 to 4.4) | 4.1 (3.8 to 4.3) | 3.8 (3.3 to 4.3) | 4.1 (3.4 to 4.9) | 3.4 (2.8 to 4.1) | 4.6 (4.1 to 5.2) | 4.1 (3.6 to 4.6) |
| Male | 40;59 | 1.5 (1.4 to 1.7) | 1.6 (1.1 to 2.3) | 1.8 (1.4 to 2.3) | 1.8 (1.2 to 2.6) | 2.4 (1.2 to 4.3) | NR | 1.4 (0.7 to 2.4) | 1.9 (1.2 to 2.8) |
| Male | 60;79 | 13.7 (13 to 14.3) | 11.5 (9.7 to 13.4) | 13 (12.1 to 14) | 12 (10.2 to 14) | 13.5 (10.8 to 16.7) | 12.5 (9.9 to 15.5) | 14 (12 to 16.2) | 13 (11.3 to 15) |
| Male | 80;150 | 25.3 (23.5 to 27.2) | 25.3 (19.9 to 31.7) | 24.3 (21.5 to 27.3) | 22.4 (17.2 to 28.7) | 23.2 (15.6 to 33.1) | 16.7 (10.6 to 25) | 29.7 (23.6 to 37) | 25.1 (19.9 to 31.3) |

Note. NR = counts <5 censored and so not reported.

| **Table S19.** Observed incidence rates (with 95% confidence intervals) of prostate cancer diagnoses in each of the time periods, overall and stratified by age and sex | | | | | | | | | |
|---|---|---|---|---|---|---|---|---|---|
| **Sex** | **Age Group (years)** | **Pre-pandemic (Jan 2017-Feb 2020)** | **Lockdown (March 2020-June 2020)** | **Post-lockdown (July 2020-Dec 2021)** | **Post-first lockdown 1 (July 2020-Oct 2020)** | **Second lockdown (Nov 2020-Dec 2020)** | **Third lockdown (Jan 2021-Feb 2021)** | **Easing of restrictions (March 2021-June 2021)** | **Legal restrictions removed (July 2021-Dec 2021)** |
| Overall | | 6.2 (6.1 to 6.3) | 4.4 (4.1 to 4.8) | 5 (4.8 to 5.2) | 4.5 (4.1 to 4.9) | 5.1 (4.6 to 5.7) | 5.1 (4.5 to 5.7) | 5.6 (5.2 to 6) | 5 (4.6 to 5.3) |
| Male | 0;150 | 12.5 (12.3 to 12.8) | 8.9 (8.2 to 9.7) | 10.1 (9.7 to 10.5) | 9.1 (8.3 to 9.8) | 10.3 (9.2 to 11.5) | 10.2 (9.1 to 11.4) | 11.3 (10.4 to 12.1) | 10.1 (9.4 to 10.8) |
| Male | 40;59 | 4.8 (4.5 to 5.1) | 2.8 (2.1 to 3.6) | 3.1 (2.7 to 3.5) | 2.3 (1.7 to 3.1) | 3.2 (2.2 to 4.6) | 3.7 (2.5 to 5.2) | 3.9 (3.1 to 5) | 2.8 (2.1 to 3.6) |
| Male | 60;79 | 45.5 (44.4 to 46.7) | 33.1 (30.1 to 36.3) | 35.9 (34.3 to 37.5) | 32.7 (29.7 to 35.9) | 37.3 (32.7 to 42.4) | 34.7 (30.2 to 39.6) | 39.3 (35.9 to 43) | 35.8 (32.9 to 38.8) |
| Male | 80;150 | 55.5 (52.9 to 58.3) | 34.1 (27.8 to 41.5) | 44.7 (40.9 to 48.8) | 44.6 (37 to 53.2) | 47.7 (36.5 to 61.3) | 44 (33.6 to 56.7) | 44.4 (36.7 to 53.2) | 44.1 (37 to 52.2) |

**Figure S9.** Incidence Rates per 100,000 Person-Years (with 95% confidence intervals) of Cancer Diagnoses in Months Before and After COVID-19 Lockdown (red dashed line) Stratified by Age and Sex

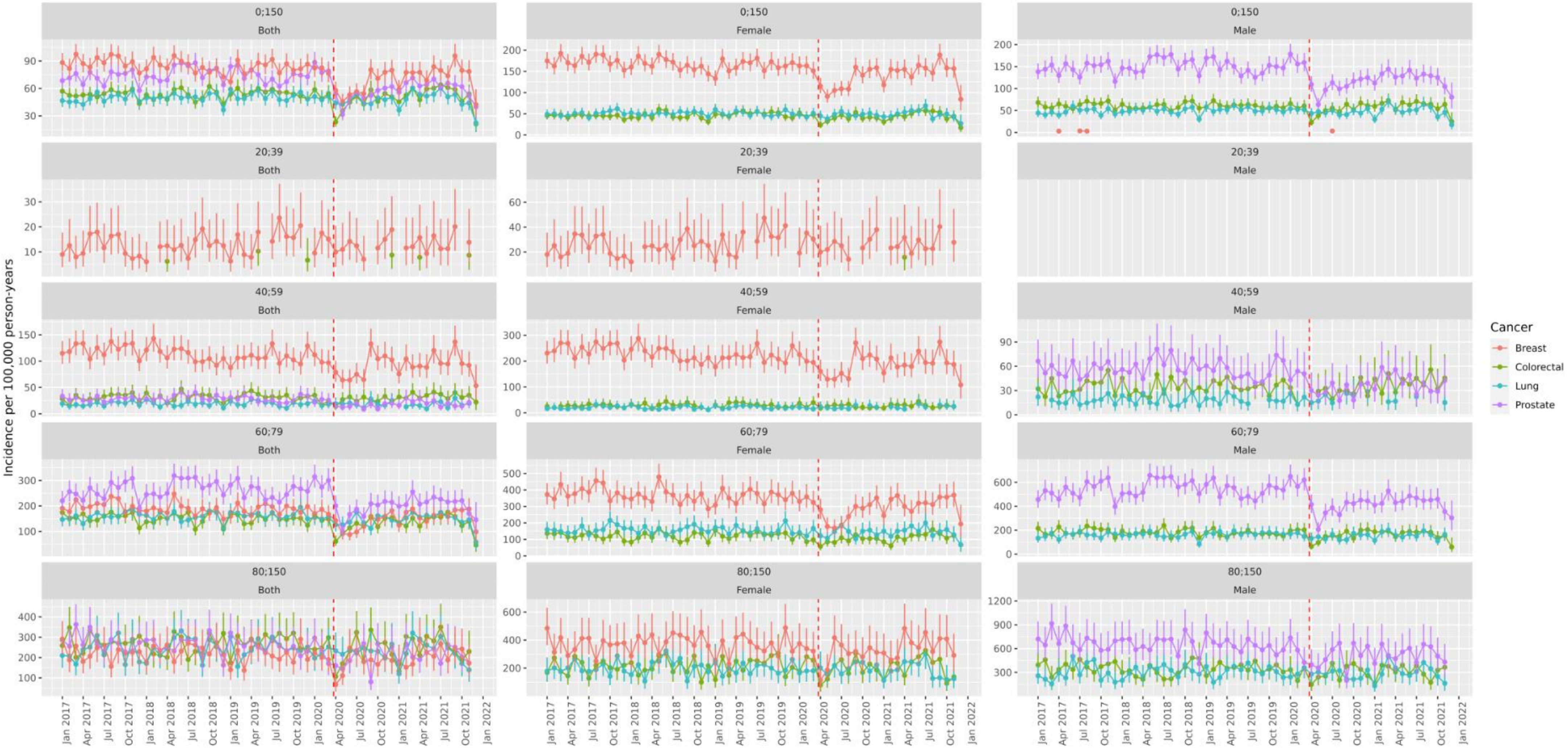

**Table S20.** Observed incidence rate ratios (with 95% confidence intervals) of breast cancer diagnoses in each of the time periods stratified by age and sex

| Sex | Age Group (years) | First Lockdown (March 2020-June 2020) | Post-first lockdown 1 (July 2020-Oct 2020) | Second lockdown (Nov 2020-Dec 2020) | Third lockdown (Jan 2021-Feb 2021) | Easing of restrictions (March 2021-June 2021 | Legal restrictions removed (July 2021-Dec 2021) |
|---|---|---|---|---|---|---|---|
| Female | 20;39 | 0.98 (0.69 to 1.37) | 0.81 (0.51 to 1.22) | 1.33 (0.85 to 1.99) | 0.92 (0.39 to 1.8) | 1.02 (0.7 to 1.43) | 1.1 (0.76 to 1.54) |
| Female | 40;59 | 0.67 (0.59 to 0.76) | 0.82 (0.73 to 0.93) | 0.94 (0.8 to 1.1) | 0.79 (0.66 to 0.94) | 0.84 (0.74 to 0.95) | 0.88 (0.79 to 0.97) |
| Female | 60;79 | 0.65 (0.57 to 0.73) | 0.68 (0.6 to 0.77) | 0.9 (0.77 to 1.04) | 0.79 (0.67 to 0.92) | 0.85 (0.76 to 0.95) | 0.9 (0.82 to 0.99) |
| Female | 80;150 | 0.7 (0.56 to 0.86) | 0.86 (0.7 to 1.04) | 0.89 (0.67 to 1.16) | 0.77 (0.57 to 1.02) | 1 (0.83 to 1.2) | 1.02 (0.86 to 1.21) |

**Table S21.** Observed incidence rate ratios (with 95% confidence intervals) of colorectal cancer diagnoses in each of the time periods, stratified by age and sex

| Sex | Age Group (years) | Lockdown (March 2020-June 2020) | Post-first lockdown 1 (July 2020-Oct 2020) | Second lockdown (Nov 2020-Dec 2020) | Third lockdown (Jan 2021-Feb 2021) | Easing of restrictions (March 2021-June 2021 | Legal restrictions removed (July 2021-Dec 2021) |
|---|---|---|---|---|---|---|---|
| Female | 40;59 | 0.98 (0.71 to 1.31) | 0.9 (0.64 to 1.22) | 0.71 (0.4 to 1.14) | 0.88 (0.53 to 1.37) | 1.12 (0.82 to 1.5) | 0.87 (0.63 to 1.18) |
| Female | 60;79 | 0.66 (0.53 to 0.8) | 0.88 (0.73 to 1.05) | 0.86 (0.66 to 1.11) | 0.6 (0.44 to 0.81) | 0.95 (0.79 to 1.13) | 1.07 (0.92 to 1.25) |
| Female | 80;150 | 0.79 (0.6 to 1.02) | 1.05 (0.82 to 1.32) | 0.97 (0.67 to 1.35) | 0.82 (0.55 to 1.16) | 1.06 (0.83 to 1.34) | 1 (0.79 to 1.25) |
| Male | 40;59 | 0.91 (0.6 to 1.33) | 0.77 (0.55 to 1.04) | 1.02 (0.67 to 1.48) | 1.06 (0.7 to 1.54) | 1.19 (0.9 to 1.54) | 1.26 (0.98 to 1.6) |
| Male | 60;79 | 0.7 (0.59 to 0.82) | 0.86 (0.73 to 1) | 1.11 (0.9 to 1.33) | 1.01 (0.82 to 1.24) | 0.96 (0.83 to 1.12) | 0.9 (0.78 to 1.03) |
| Male | 80;150 | 0.73 (0.56 to 0.94) | 1.02 (0.81 to 1.28) | 0.87 (0.6 to 1.21) | 0.79 (0.54 to 1.11) | 0.99 (0.78 to 1.24) | 0.89 (0.7 to 1.11) |

**Table S22.** Observed incidence rate ratios (with 95% confidence intervals) of lung cancer diagnoses in each of the time periods, stratified by age and sex

| Sex | Age Group (years) | Lockdown (March 2020-June 2020) | Post-first lockdown 1 (July 2020-Oct 2020) | Second lockdown (Nov 2020-Dec 2020) | Third lockdown (Jan 2021-Feb 2021) | Easing of restrictions (March 2021-June 2021) | Legal restrictions removed (July 2021-Dec 2021) |
|---|---|---|---|---|---|---|---|
| Female | 40;59 | 0.88 (0.59 to 1.27) | 1.04 (0.68 to 1.53) | 1.18 (0.71 to 1.84) | 1.17 (0.53 to 2.21) | 1.1 (0.71 to 1.62) | 1.25 (0.87 to 1.74) |
| Female | 60;79 | 0.86 (0.73 to 1.01) | 0.94 (0.8 to 1.1) | 0.9 (0.71 to 1.12) | 0.86 (0.68 to 1.08) | 1.01 (0.87 to 1.18) | 0.93 (0.81 to 1.07) |
| Female | 80;150 | 0.94 (0.72 to 1.19) | 1.04 (0.81 to 1.31) | 1 (0.69 to 1.4) | 0.72 (0.47 to 1.05) | 1.13 (0.88 to 1.42) | 0.8 (0.61 to 1.03) |
| Male | 40;59 | 1.07 (0.73 to 1.52) | 1.16 (0.75 to 1.71) | 1.58 (0.81 to 2.74) | NR | 0.9 (0.48 to 1.53) | 1.22 (0.78 to 1.83) |
| Male | 60;79 | 0.84 (0.71 to 0.99) | 0.88 (0.75 to 1.03) | 0.99 (0.79 to 1.22) | 0.91 (0.72 to 1.14) | 1.02 (0.88 to 1.19) | 0.96 (0.82 to 1.1) |
| Male | 80;150 | 1 (0.79 to 1.26) | 0.89 (0.68 to 1.14) | 0.92 (0.63 to 1.3) | 0.66 (0.43 to 0.98) | 1.18 (0.93 to 1.47) | 0.99 (0.78 to 1.25) |

Note. NR = counts <5 censored and so not reported.

**Table S23.** Observed incidence rate ratios (with 95% confidence intervals) of prostate cancer diagnoses in each of the time periods, stratified by age and sex

| Sex | Age Group (years) | Lockdown (March 2020-June 2020) | Post-first lockdown 1 (July 2020-Oct 2020) | Second lockdown (Nov 2020-Dec 2020) | Third lockdown (Jan 2021-Feb 2021) | Easing of restrictions (March 2021-June 2021) | Legal restrictions removed (July 2021-Dec 2021) |
|---|---|---|---|---|---|---|---|
| Male | 40;59 | 0.58 (0.44 to 0.76) | 0.49 (0.36 to 0.66) | 0.68 (0.46 to 0.96) | 0.78 (0.54 to 1.09) | 0.83 (0.64 to 1.05) | 0.58 (0.44 to 0.75) |
| Male | 60;79 | 0.73 (0.66 to 0.8) | 0.72 (0.65 to 0.79) | 0.82 (0.72 to 0.93) | 0.76 (0.66 to 0.87) | 0.86 (0.79 to 0.95) | 0.79 (0.72 to 0.86) |
| Male | 80;150 | 0.62 (0.5 to 0.75) | 0.8 (0.66 to 0.96) | 0.86 (0.66 to 1.1) | 0.79 (0.61 to 1.02) | 0.8 (0.66 to 0.96) | 0.8 (0.66 to 0.94) |

**Table S24.** Estimated percentage reduction, and estimated underdiagnoses (missed counts) (with 95% confidence intervals) for each cancer across the extended lockdown periods from the negative binomial modelling

| Cancer | Parameter | First Lockdown (March 2020-June 2020) | Post-lockdown (July 2020-Dec 2021) | Post-first lockdown 1 (July 2020-Oct 2020) | Second lockdown (Nov 2020-Dec 2020) | Third lockdown (Jan 2021-Feb 2021) | Easing of restrictions (March 2021-June 2021) | Legal restrictions removed (July 2021-Dec 2021) | Total (March 2020-Dec 2021) |
|---|---|---|---|---|---|---|---|---|---|
| Breast | Percent reduction | 32% (19.5 to 41.9) | 14.3% (-3.7 to 27.7) | 22.2% (7 to 33.8) | 4.9% (-14.2 to 19.2) | 20.6% (4.4 to 32.7) | 15% (-3.6 to 28.5) | 8.1% (-12.2 to 23.4) | 17.9% (1.1 to 30.6) |
| Breast | N underdiagnoses | 311 (160 to 475) | 541 (-117 to 1246) | 204 (54 to 366) | 22 (-52 to 99) | 90 (16 to 168) | 138 (-27 to 311) | 88 (-108 to 302) | 852 (43 to 1721) |
| Colorectal | Percent reduction | 33.9% (18.1 to 45.4) | 14.1% (-7.5 to 30.3) | 16.4% (-3.6 to 31.1) | 4.1% (-19.2 to 22.1) | 20.2% (1.2 to 34.7) | 14.1% (-8.2 to 30.7) | 13.6% (-9.3 to 30.7) | 18.1% (-2.4 to 33.3) |
| Colorectal | N underdiagnoses | 230 (99 to 373) | 385 (-164 to 1019) | 107 (-19 to 246) | 12 (-45 to 79) | 63 (3 to 132) | 95 (-44 to 256) | 109 (-59 to 306) | 615 (-65 to 1392) |
| Lung | Percent reduction | 15.4% (-5.2 to 30.4) | 13.9% (-9.4 to 30.4) | 12.9% (-9.4 to 28.5) | 7.3% (-16.1 to 24.4) | 25.5% (6.1 to 39.2) | 8.4% (-17.1 to 26.5) | 17.2% (-6.8 to 33.9) | 14.2% (-8.6 to 30.4) |
| Lung | N underdiagnoses | 96 (-26 to 229) | 349 (-187 to 946) | 77 (-45 to 209) | 20 (-36 to 84) | 73 (14 to 138) | 51 (-81 to 200) | 128 (-39 to 315) | 445 (-213 to 1175) |
| Prostate | Percent reduction | 28.6% (8.9 to 42.7) | 22.2% (-0.7 to 38.3) | 28.5% (8.9 to 42.4) | 18.1% (-4.9 to 34.4) | 25.5% (6.1 to 40.1) | 13.9% (-13.3 to 32.5) | 23.7% (-0.1 to 40.4) | 23.4% (1.2 to 39.2) |
| Prostate | N underdiagnoses | 241 (59 to 449) | 755 (-18 to 1647) | 237 (58 to 438) | 71 (-15 to 170) | 106 (20 to 207) | 110 (-80 to 328) | 231 (-1 to 504) | 997 (41 to 2096) |

**Table S25.** Estimated number of underdiagnosed cancer cases, extrapolated to the UK general population over each lockdown period

| Cancer | Lockdown (March 2020-June 2020) | Post-lockdown (July 2020-Dec 2021) | Post-first lockdown 1 (July 2020-Oct 2020) | Second lockdown (Nov 2020-Dec 2020) | Third lockdown (Jan 2021-Feb 2021) | Easing of restrictions (March 2021-June 2021) | Legal restrictions removed (July 2021-Dec 2021) | Total (March 2020-Dec 2021) |
|---|---|---|---|---|---|---|---|---|
| Breast | 6,714 | 11,680 | 4,404 | 475 | 1,943 | 2,979 | 1,900 | 18,395 |
| Colorectal | 4,966 | 8,312 | 2,310 | 259 | 1,360 | 2,051 | 2,353 | 13,278 |
| Lung | 2,073 | 7,535 | 1,662 | 432 | 1,576 | 1,101 | 2,764 | 9,608 |
| Prostate | 5,203 | 16,300 | 5,117 | 1,533 | 2,289 | 2,375 | 4,987 | 21,525 |

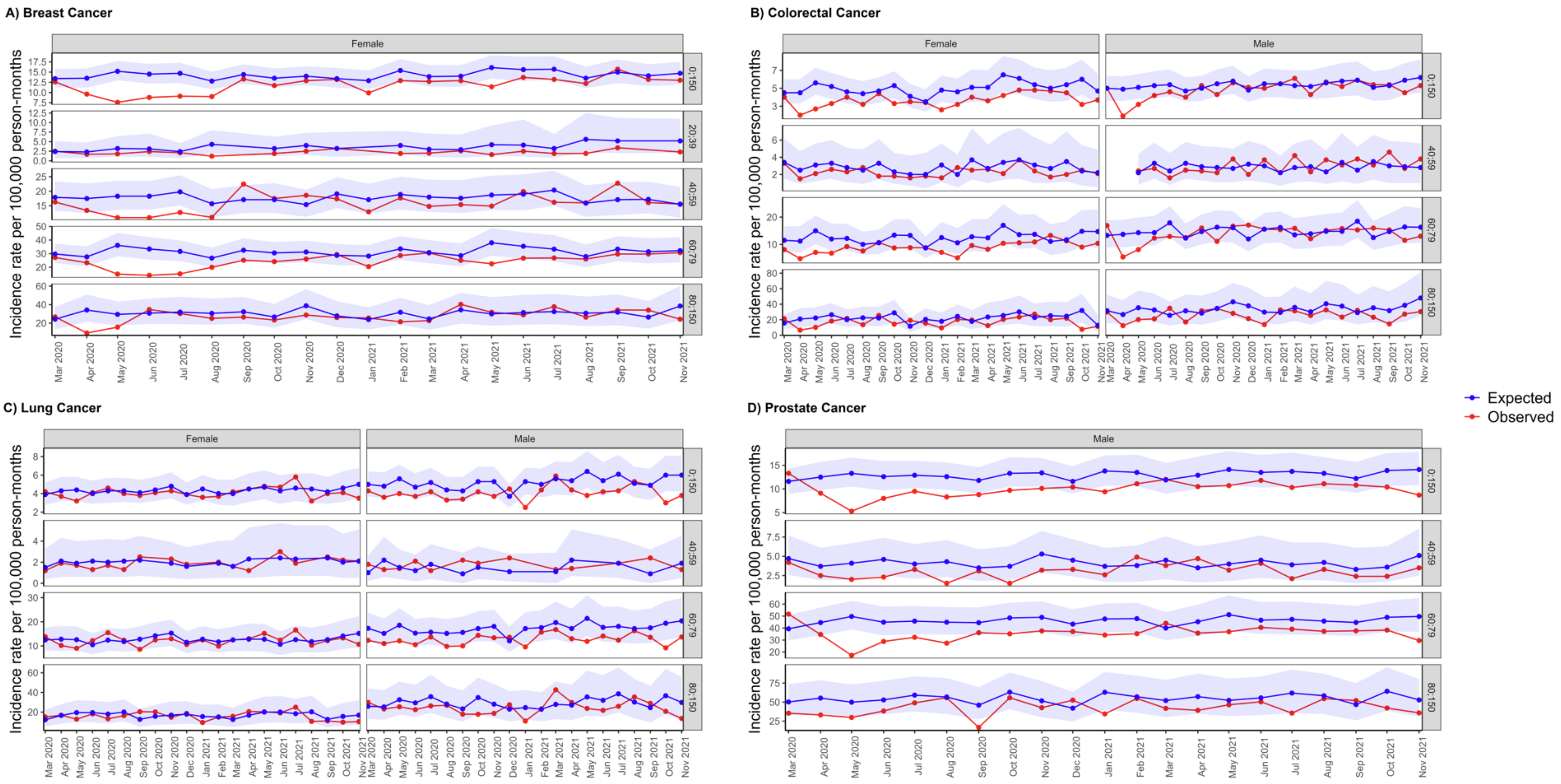


**Figure S10.** Expected and observed incidence rates per 100,000 person months of A) breast cancer, B) colorectal cancer, C) lung cancer, and D) prostate cancer in primary care records from CPRD GOLD UK, after lockdown, stratified by age and sex. Points represent monthly incidence rates. Expected rates (with 95% prediction intervals represented by shaded areas) were calculated using negative binomial regression using observed data from January 2017 to February 2020 to estimate expected counts from March 2020 to November 2021.

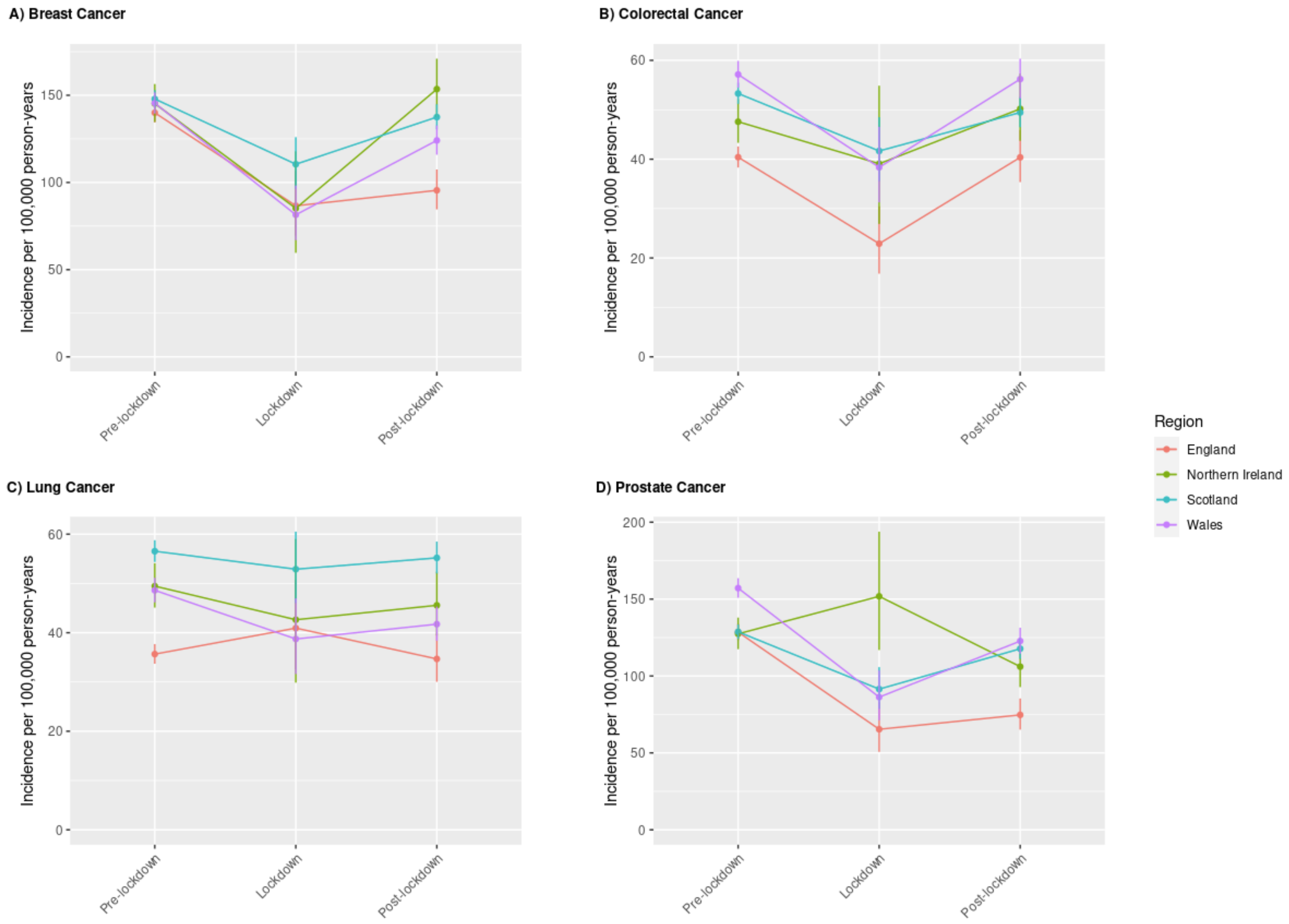


**Figure S11.** Incidence Rates per 100,000 Person-Years (with 95% confidence intervals) of Cancer Diagnoses in Months Before and After COVID-19 Lockdown (red dashed line) Stratified by UK Region

**Table S26**. Estimated percentage reduction, and estimated underdiagnoses (missed counts) (with 95% confidence intervals) for breast cancer across the extended lockdown periods from the negative binomial modelling, stratified by age and sex

| Sex/Age Group | Parameter | Lockdown (March 2020-June 2020) | Post-lockdown (July 2020-Dec 2021) | Post-lockdown 1 (July 2020-Oct 2020) | Second lockdown (Nov 2020-Dec 2020) | Third lockdown (Jan 2021-Feb 2021) | Easing of restrictions (March 2021-June 2021) | Legal restrictions removed (July 2021-Dec 2021) | Total (March 2020-Dec 2021) |
|---|---|---|---|---|---|---|---|---|---|
| Female: 20;39 | Percent reduction | 23.2% (-140 to 64) | 42.6% (-63 to 73.3) | 47.4% (-37.5 to 72.5) | 19.9% (-130 to 61.7) | 51.9% (-16.7 to 75) | 38.2% (-106.2 to 74.2) | 50.7% (-36 to 77.3) | 39.1% (-76.1 to 71.6) |
| Female: 20;39 | N underdiagnoses | 11 (-21 to 64) | 88 (-46 to 327) | 20 (-6 to 58) | 6 (-13 to 37) | 8 (-1 to 21) | 20 (-17 to 95) | 35 (-9 to 116) | 99 (-67 to 391) |
| Female: 40;59 | Percent reduction | 28.5% (3.2 to 44.7) | 6.4% (-29.7 to 29.8) | 9.4% (-23.6 to 30.6) | -4% (-41.4 to 20.7) | 15.8% (-16.5 to 36.2) | 11.5% (-23.7 to 34.5) | -0.4% (-41.3 to 26.1) | 11% (-22.7 to 32.9) |
| Female: 40;59 | N underdiagnoses | 96 (8 to 196) | 81 (-273 to 506) | 30 (-55 to 127) | -6 (-46 to 41) | 24 (-18 to 72) | 35 (-52 to 143) | -1 (-102 to 123) | 178 (-265 to 702) |
| Female: 60;79 | Percent reduction | 37.4% (18.4 to 50.3) | 18.6% (-9.1 to 36.8) | 30.9% (9.7 to 45.5) | 7.6% (-22.7 to 27.6) | 21% (-4.6 to 38.2) | 21.1% (-6.5 to 39) | 9.6% (-24.3 to 30.8) | 22.5% (-3.4 to 39.5) |
| Female: 60;79 | N underdiagnoses | 170 (64 to 287) | 329 (-120 to 835) | 130 (31 to 242) | 15 (-34 to 70) | 43 (-7 to 99) | 92 (-21 to 221) | 49 (-89 to 203) | 498 (-56 to 1122) |
| Female: 80;150 | Percent reduction | 27.1% (-20.8 to 51.3) | 6.8% (-61.5 to 39.9) | 13.1% (-46.6 to 42.5) | 17.6% (-36.8 to 45.3) | 14.1% (-51.6 to 43.4) | -2.7% (-81.8 to 35.1) | 1.5% (-73.5 to 38.2) | 10.9% (-53 to 42.1) |
| Female: 80;150 | N underdiagnoses | 35 (-16 to 98) | 34 (-179 to 312) | 16 (-34 to 79) | 11 (-14 to 43) | 8 (-16 to 36) | -3 (-54 to 65) | 2 (-61 to 89) | 69 (-195 to 410) |

**Table S27.** Estimated percentage reduction, and estimated underdiagnoses (missed counts) (with 95% confidence intervals) for colorectal cancer across the extended lockdown periods from the negative binomial modelling, stratified by age and sex

| Sex/Age Group | Parameter | Lockdown (March 2020-June 2020) | Post-lockdown (July 2020-Dec 2021) | Post-lockdown 1 (July 2020-Oct 2020) | Second lockdown (Nov 2020-Dec 2020) | Third lockdown (Jan 2021-Feb 2021) | Easing of restrictions (March 2021-June 2021) | Legal restrictions removed (July 2021-Dec 2021) | Total (March 2020-Dec 2021) |
|---|---|---|---|---|---|---|---|---|---|
| Male: 40;59 | Percent reduction | 5.8% (-127.3 to 49) | -5.8% (-174.4 to 45) | 23.4% (-86.4 to 58.2) | 2% (-136.4 to 46.9) | -15% (-225 to 38.1) | -16.6% (-205.3 to 42) | -22.7% (-236.4 to 38.3) | -4.5% (-168.8 to 45.4) |
| Male: 40;59 | N underdiagnoses | 2 (-14 to 24) | -12 (-143 to 184) | 13 (-19 to 57) | 1 (-15 to 23) | -3 (-18 to 16) | -8 (-39 to 42) | -14 (-52 to 46) | -11 (-157 to 208) |
| Male: 60;79 | Percent reduction | 23.1% (-15.9 to 45.3) | 3.6% (-47.1 to 32.1) | 14.5% (-24.5 to 38.4) | -20.6% (-87.7 to 14.4) | 2.4% (-46.3 to 29) | -3.5% (-64.6 to 28.7) | 9.1% (-40 to 37.1) | 7.3% (-41.1 to 34.6) |
| Male: 60;79 | N underdiagnoses | 44 (-20 to 121) | 29 (-250 to 369) | 29 (-34 to 108) | -18 (-50 to 18) | 2 (-31 to 40) | -6 (-73 to 75) | 22 (-62 to 128) | 73 (-270 to 490) |
| Male: 80;150 | Percent reduction | 33.5% (-26.5 to 59.7) | 22.4% (-57.1 to 54.5) | 2.1% (-97.6 to 42.1) | 38.7% (-18.5 to 61.4) | 23.9% (-55 to 54.4) | 21.4% (-58.3 to 55) | 29.9% (-45.5 to 60) | 24.5% (-50.8 to 55.5) |
| Male: 80;150 | N underdiagnoses | 31 (-13 to 92) | 86 (-109 to 360) | 2 (-40 to 59) | 20 (-5 to 51) | 10 (-11 to 37) | 21 (-28 to 93) | 34 (-25 to 120) | 118 (-122 to 452) |
| Female: 40;59 | Percent reduction | 22.9% (-87.5 to 57.9) | 19.8% (-128.2 to 60.1) | 18.7% (-110.5 to 57.4) | 14.2% (-200 to 60.5) | 16.7% (-157.1 to 57.1) | 19.3% (-109.1 to 59.6) | 24% (-138.9 to 63.6) | 20.5% (-117.9 to 59.6) |
| Female: 40;59 | N underdiagnoses | 13 (-21 to 62) | 40 (-91 to 244) | 9 (-21 to 54) | 2 (-10 to 23) | 4 (-11 to 24) | 11 (-24 to 68) | 14 (-25 to 75) | 53 (-112 to 306) |
| Female: 60;79 | Percent reduction | 45.8% (16.4 to 62.3) | 25.1% (-22 to 50) | 21.8% (-24.8 to 45.9) | 19.7% (-31.1 to 44.9) | 46.8% (14.6 to 64) | 30.6% (-13.2 to 53.9) | 16.8% (-38.3 to 46.2) | 29.3% (-13.9 to 52.3) |
| Female: 60;79 | N underdiagnoses | 82 (19 to 160) | 178 (-96 to 531) | 35 (-25 to 107) | 14 (-14 to 48) | 36 (7 to 73) | 57 (-15 to 151) | 36 (-49 to 152) | 260 (-77 to 691) |
| Female: 80;150 | Percent reduction | 33.6% (-24.5 to 59.1) | 20.5% (-60.7 to 53.6) | 19.9% (-49 to 50.6) | -7.8% (-135.7 to 38.9) | 30.4% (-38.1 to 57.4) | 21.3% (-60.9 to 55.4) | 24.4% (-60.8 to 57.1) | 23.1% (-53 to 54.6) |
| Female: 80;150 | N underdiagnoses | 31 (-12 to 88) | 76 (-111 to 339) | 19 (-25 to 78) | -2 (-19 to 21) | 13 (-8 to 39) | 20 (-28 to 92) | 26 (-31 to 109) | 106 (-123 to 427) |

**Table S28.** Estimated percentage reduction, and estimated underdiagnoses (missed counts) (with 95% confidence intervals) for lung cancer across the extended lockdown periods from the negative binomial modelling, stratified by age and sex

| Sex/Age Group | Parameter | Lockdown (March 2020-June 2020) | Post-lockdown (July 2020-Dec 2021) | Post-lockdown 1 (July 2020-Oct 2020) | Second lockdown (Nov 2020-Dec 2020) | Third lockdown (Jan 2021-Feb 2021) | Easing of restrictions (March 2021-June 2021) | Legal restrictions removed (July 2021-Dec 2021) | Total (March 2020-Dec 2021) |
|---|---|---|---|---|---|---|---|---|---|
| Male: 40;59 | Percent reduction | -12.1% (-357.1 to 49.2) | -22.2% (-491.7 to 50.7) | -26.9% (-525 to 46.8) | -111.9% (-1000 to 15.4) | NR | 15.2% (-300 to 65.7) | -20.5% (-475 to 53.1) | -18.8% (-442.1 to 50.2) |
| Male: 40;59 | N underdiagnoses | -3 (-25 to 31) | -13 (-59 to 73) | -5 (-21 to 22) | -6 (-10 to 2) | NR | 2 (-9 to 23) | -4 (-19 to 26) | -16 (-84 to 104) |
| Male: 60;79 | Percent reduction | 31% (-1.9 to 50.2) | 25.5% (-13.3 to 48.1) | 24.5% (-12 to 46.3) | 10% (-38.7 to 36.8) | 28.2 (-6.8 to 48.7) | 26.4% (-12 to 49.7) | 29.5% (-9.3 to 51.6) | 26.5% (-11 to 48.5) |
| Male: 60;79 | N underdiagnoses | 71 (-3 to 158) | 239 (-82 to 649) | 52 (-17 to 137) | 10 (-24 to 50) | 31 (-5 to 75) | 63 (-19 to 175) | 83 (-17 to 212) | 310 (-85 to 807) |
| Male: 80;150 | Percent reduction | 10.5% (-74.4 to 45.3) | 18% (-72.3 to 52.9) | 26.8% (-44.2 to 55.7) | 9.4% (-87.5 to 48.3) | 29.9 (-43.8 to 59.6) | 2.8% (-110.5 to 45.6) | 21.8% (-71.7 to 56.1) | 16.5% (-72.8 to 51.5) |
| Male: 80;150 | N underdiagnoses | 9 (-32 to 62) | 60 (-115 to 308) | 23 (-19 to 78) | 3 (-14 to 28) | 10 (-7 to 34) | 2 (-42 to 67) | 22 (-33 to 101) | 69 (-147 to 370) |
| Female: 40;59 | Percent reduction | 17.9% (-163.6 to 60.8) | 2.9% (-243.8 to 56.7) | 13.1% (-177.8 to 57.6) | -18.9% (-350 to 43.8) | -7.8 (-300 to 50) | 9.1% (-242.9 to 61.9) | 1.5% (-250 to 58.3) | 6.5% (-223.3 to 57.6) |
| Female: 40;59 | N underdiagnoses | 6 (-18 to 45) | 3 (-78 to 144) | 4 (-16 to 34) | -3 (-14 to 14) | -1 (-6 to 8) | 2 (-17 to 39) | 1 (-25 to 49) | 10 (-96 to 189) |
| Female: 60;79 | Percent reduction | 6.5% (-46.8 to 34.8) | 2.6% (-55.7 to 32.7) | 3.6% (-48.3 to 31.7) | 11.9% (-38.6 to 36.3) | 8.2 (-44.2 to 35.9) | -8.8% (-78.8 to 26.9) | 4.4% (-56.9 to 35.2) | 3.3% (-54 to 33.1) |
| Female: 60;79 | N underdiagnoses | 11 (-52 to 87) | 19 (-253 to 343) | 6 (-56 to 80) | 11 (-22 to 45) | 7 (-23 to 42) | -14 (-78 to 65) | 9 (-74 to 111) | 30 (-305 to 430) |
| Female: 80;150 | Percent reduction | 7.1% (-88.9 to 44.7) | 5% (-112.1 to 47.2) | -4.6% (-121.9 to 38.8) | 3.8% (-113.3 to 45.8) | 21.7 (-71.4 to 56.4) | -9.5% (-146.7 to 40.3) | 19.9% (-87.9 to 56.9) | 5.4% (-106.9 to 46.7) |
| Female: 80;150 | N underdiagnoses | 5 (-32 to 55) | 14 (-139 to 235) | -3 (-39 to 45) | 1 (-17 to 27) | 7 (-10 to 31) | -6 (-44 to 50) | 15 (-29 to 82) | 19 (-171 to 290) |

Note. NR = counts <5 censored and so not reported.

**Table 29.** Estimated percentage reduction, and estimated underdiagnoses (missed counts) (with 95% confidence intervals) for prostate cancer across the extended lockdown periods from the negative binomial modelling, stratified by age and sex

| Sex/Age Group | Parameter | Lockdown (March 2020-June 2020) | Post-lockdown (July 2020-Dec 2021) | Post-lockdown 1 (July 2020-Oct 2020) | Second lockdown (Nov 2020-Dec 2020) | Third lockdown (Jan 2021-Feb 2021) | Easing of restrictions (March 2021-June 2021) | Legal restrictions removed (July 2021-Dec 2021) | Total (March 2020-Dec 2021) |
|---|---|---|---|---|---|---|---|---|---|
| Male: 40;59 | Percent reduction | 35.3% (-22.7 to 60) | 24.5% (-54.4 to 55.1) | 40.3% (-15.8 to 63.9) | 34.5% (-20.8 to 58.6) | 1.9% (-113.3 to 40.7) | 4.4% (-106.1 to 44.7) | 31.2% (-46.2 to 60.1) | 26.8% (-47.2 to 56.1) |
| Male: 40;59 | N underdiagnoses | 29 (-10 to 81) | 75 (-81 to 282) | 30 (-6 to 78) | 15 (-5 to 41) | 1 (-17 to 22) | 3 (-35 to 55) | 26 (-18 to 86) | 104 (-91 to 363) |
| Male: 60;79 | Percent reduction | 25.8% (2.8 to 42) | 22.4% (-3.2 to 40) | 28.9% (7.2 to 44.2) | 19.1% (-7.8 to 36.6) | 27.3% (5.2 to 42.9) | 14% (-15.8 to 35) | 22.9% (-3.8 to 40.9) | 23% (-2 to 40.4) |
| Male: 60;79 | N underdiagnoses | 156 (13 to 324) | 552 (-59 to 1280) | 173 (33 to 338) | 55 (-17 to 135) | 82 (12 to 163) | 80 (-67 to 264) | 163 (-20 to 380) | 708 (-46 to 1604) |
| Male: 80;150 | Percent reduction | 34.4% (-3.1 to 55) | 19.3% (-33.9 to 45.8) | 20.9% (-27.1 to 45) | -1.9% (-74.3 to 32.2) | 26.9% (-15.4 to 49.6) | 18.2% (-37.2 to 46.4) | 22.5% (-33 to 48.9) | 22.3% (-27.5 to 47.6) |
| Male: 80;150 | N underdiagnoses | 52 (-3 to 122) | 119 (-126 to 421) | 32 (-26 to 100) | -1 (-26 to 29) | 22 (-8 to 59) | 26 (-32 to 102) | 40 (-34 to 131) | 172 (-129 to 543) |

**Table S30.** Estimated number of underdiagnosed cancer cases, extrapolated to the UK general population over each lockdown period, stratified by age and sex

| **Cancer** | **Sex/Age Group** | **Lockdown (March 2020-June 2020)** | **Post-lockdown (July 2020-Dec 2021)** | **Post-lockdown 1 (July 2020-Oct 2020)** | **Second lockdown (Nov 2020-Dec 2020)** | **Third lockdown (Jan 2021-Feb 2021)** | **Easing of restrictions (March 2021-June 2021)** | **Legal restrictions removed (July 2021-Dec 2021)** | **Total (March 2020-Dec 2021)** |
|---|---|---|---|---|---|---|---|---|---|
| Breast | | | | | | | | | |
| | Female: 20;39 | 237 | 1,900 | 432 | 130 | 173 | 432 | 756 | 2,137 |
| | Female: 40;59 | 2,073 | 1,749 | 648 | -130 | 518 | 756 | -22 | 3,843 |
| | Female: 60;79 | 3,670 | 7,103 | 2,807 | 324 | 928 | 1,986 | 1,058 | 10,752 |
| | Female: 80;150 | 756 | 734 | 345 | 237 | 173 | -65 | 43 | 1,490 |
| Colorectal | | | | | | | | | |
| | Male: 40;59 | 43 | -259 | 281 | 22 | -65 | -173 | -302 | -237 |
| | Male: 60;79 | 950 | 626 | 626 | -389 | 43 | -130 | 475 | 1,576 |
| | Male: 80;150 | 669 | 1,857 | 43 | 432 | 216 | 453 | 734 | 2,548 |
| | Female: 40;59 | 281 | 864 | 194 | 43 | 86 | 237 | 302 | 1,144 |
| | Female: 60;79 | 1,770 | 3,843 | 756 | 302 | 777 | 1,231 | 777 | 5,613 |
| | Female: 80;150 | 669 | 1,641 | 410 | -43 | 281 | 432 | 561 | 2,289 |
| Lung | | | | | | | | | |
| | Male: 40;59 | -65 | -281 | -108 | -130 | NR | 43 | -86 | -345 |
| | Male: 60;79 | 1,533 | 5,160 | 1,123 | 216 | 669 | 1,360 | 1,792 | 6,693 |
| | Male: 80;150 | 194 | 1,295 | 497 | 65 | 216 | 43 | 475 | 1,490 |

**Table S30.** Estimated number of underdiagnosed cancer cases, extrapolated to the UK general population over each lockdown period, stratified by age and sex

| Cancer | Sex/Age Group | Lockdown (March 2020-June 2020) | Post-lockdown (July 2020-Dec 2021) | Post-lockdown 1 (July 2020-Oct 2020) | Second lockdown (Nov 2020-Dec 2020) | Third lockdown (Jan 2021-Feb 2021) | Easing of restrictions (March 2021-June 2021) | Legal restrictions removed (July 2021-Dec 2021) | Total (March 2020-Dec 2021) |
|---|---|---|---|---|---|---|---|---|---|
| | Female: 40;59 | 130 | 65 | 86 | -65 | -22 | 43 | 22 | 216 |
| | Female: 60;79 | 237 | 410 | 130 | 237 | 151 | -302 | 194 | 648 |
| | Female: 80;150 | 108 | 302 | -65 | 22 | 151 | -130 | 324 | 410 |
| Prostate | | | | | | | | | |
| | Male: 40;59 | 626 | 1,619 | 648 | 324 | 22 | 65 | 561 | 2,245 |
| | Male: 60;79 | 3,368 | 11,918 | 3,735 | 1,187 | 1,770 | 1,727 | 3,519 | 15,286 |
| | Male: 80;150 | 1,123 | 2,569 | 691 | -22 | 475 | 561 | 864 | 3,713 |

Note. NR = counts <5 censored and so not reported.
